# Clinical and Virological Response to a Neutralizing Monoclonal Antibody for Hospitalized Patients with COVID-19

**DOI:** 10.1101/2021.07.19.21260559

**Authors:** ACTIV-3/TICO Bamlanivimab Study Group, Jens D. Lundgren, Birgit Grund, Christina E. Barkauskas, Thomas L. Holland, Robert L. Gottlieb, Uriel Sandkovsky, Samuel M. Brown, Kirk U. Knowlton, Wesley H. Self, D. Clark Files, Mamta K. Jain, Thomas Benfield, Michael E. Bowdish, Bradley G. Leshnower, Jason V. Baker, Jens-Ulrik Jensen, Edward M. Gardner, Adit A. Ginde, Estelle S. Harris, Isik S. Johansen, Norman Markowitz, Michael A. Matthay, Lars Østergaard, Christina C. Chang, Anna Goodman, Weizhong Chang, Robin L. Dewar, Norman P. Gerry, Elizabeth S. Higgs, Helene Highbarger, Daniel D. Murray, Thomas A. Murray, Ven Natarajan, Roger Paredes, Mahesh K.B. Parmar, Andrew N. Phillips, Cavan Reilly, Adam W. Rupert, Shweta Sharma, Kathryn Shaw-Saliba, Brad T. Sherman, Marc Teitelbaum, Deborah Wentworth, Huyen Cao, Paul Klekotka, Abdel G. Babiker, Victoria J. Davey, Annetine C. Gelijns, Virginia L. Kan, Mark N. Polizzotto, B. Taylor Thompson, H. Clifford Lane, James D. Neaton

## Abstract

**BACKGROUND:** Bamlanivimab, a neutralizing monoclonal antibody given in combination with remdesivir, did not improve outcomes among hospitalized COVID-19 patients based on an early futility assessment. In this final study report, we evaluate an *a priori* hypothesis that greater benefit of bamlanivimab would be identified in those without detectable endogenous neutralizing antibody levels at study entry, especially if viral levels were high.

**METHODS:** Hospitalized COVID-19 patients were randomized to receive bamlanivimab (7000mg) or placebo and followed for 90 days for sustained recovery (home for 14 consecutive days); recovery rate ratios (RRRs) are cited.

**RESULTS:** Among 314 participants (163 on bamlanivimab and 151 on placebo), the median time to sustained recovery was 19 days and RRR=0.99 (95% CI: 0.79-1.22; p=0.89). At entry, 50% evidenced production of anti-spike neutralizing antibodies; 50% had SARS-CoV-2 nucleocapsid plasma antigen levels ≥ 1,000 ng/L. Among those without and with antibodies at study entry, the RRRs were 1.24 (95% CI: 0.90-1.70) and 0.74 (95% CI: 0.54-1.00) (p=0.02 for interaction). The RRRs were elevated for those with plasma antigen or nasal viral RNA levels above versus below median at entry, and was greatest for those without antibodies and with elevated antigen or viral RNA levels: 1.48 (95% CI: 0.99-2.23), 1.94 (1.25-3.00), respectively (p<0.05 for all interactions). Hazard ratios for safety outcomes also differed by serostatus at entry.

**CONCLUSIONS:** Sustained recovery after administration of bamlanivimab versus placebo differed by presence of neutralizing antibodies at study entry, especially if participants had markers of elevated viral replication.

ClinicalTrials.gov number, NCT04501978.

## Introduction

SARS-CoV-2 neutralizing monoclonal antibodies may reduce hospitalization risk among outpatients with early COVID-19 and appear to accelerate viral load decline in the nasopharynx^1-6^. The US FDA has issued emergency use authorization for several such products^7-10^.

Preliminary results of the ACTIV-3/TICO trial of bamlanivimab in hospitalized patients with COVID-19 were reported after enrolment was terminated because futility guidelines were met^11,12^. Similarly, other neutralizing monoclonal products, convalescent plasma, and hyperimmune immunoglobulin products have not appeared to provide overall clinical benefit for such patients^13-16^.

We report here the final results of the bamlanivimab trial after completing the planned 90-day follow-up. In addition to clinical characteristics, plasma binding antibody and antigen levels, nasal viral RNA levels, and inflammatory markers were determined. These data were used to test an *a priori* hypothesis that patients without neutralizing antibodies at entry would benefit more from bamlanivimab and that benefit would be greatest in those patients with high plasma antigen levels or with high nasal viral RNA levels.

## Methods

### Design and Treatments

As previously described, this randomized placebo controlled study compared bamlanivimab (7000mg) with placebo, administered as a single intravenous infusion over a 1-hour period^12,17^. The infusion was prepared by an unblinded pharmacist. All other site personnel, and study participants, were blinded to treatment assignment.

### Patients

We enrolled adult hospitalized patients without invasive mechanical ventilation and with documented SARS-CoV-2 infection who had symptoms attributable to COVID-19 for ≤ 12 days. Detailed eligibility criteria, human subjects protection, and written consent have been reported previously^17^ and are given in the supplementary appendix. All patients received remdesivir.

### Outcomes

Two ordinal outcomes termed pulmonary and pulmonary-plus assessed at day 5 following infusion were used to assess futility after at least 300 patients. These outcomes are defined in the supplementary appendix (pulmonary outcome is shown in **Figure S1**)^17^.

The primary endpoint for agents studied in TICO is time to sustained recovery, defined as being discharged to home and remaining at home for at least 14 days, through 90 days of follow-up. By definition, the shortest possible time to sustained recovery is 14 days following randomization.

Other outcomes included composite safety outcomes assessed through days 5, 28 and 90, and mortality. These outcomes are defined in the supplementary appendix. Results as of day 5 and partly as of day 28 have been reported previously^17^.

### Serologic and Virologic Assays on Stored Samples

Details concerning the *a priori* hypotheses that were developed for antibody and viral levels and laboratory methods are provided in the supplementary appendix. Briefly, SARS-CoV-2 viral RNA levels were measured from a mid-turbinate nasal swab. Sequences were aligned to SARS-CoV-2 reference (Genbank) and assigned Nextstrain clades and PANGO lineages. Plasma samples collected at study entry and at days 1, 3 and 5 were used to measure anti-spike receptor binding domain (RBD) neutralizing antibodies (nAbs) in a surrogate viral neutralization test, and anti-nucleocapsid (anti-N) binding antibody levels. Qualitative plasma SARS-CoV-2 N antigen was measured using a microbead-based immunoassay. Stored samples were used to measure plasma levels of interleukin-6 (IL-6) and D-dimer and serum levels of C-reactive protein (CRP).

### Statistical Analyses

The analyses in this report are based on the same population described in our preliminary report^17^. This modified intention-to-treat analysis population was restricted to randomized patients who received any volume of bamlanivimab or placebo (**Figure S2**).

Antibody levels assessed during follow-up are reported as the percentage of patients positive or negative for nAbs and for anti-N antibodies. Data for each antibody assay are summarized for the overall cohort and are reported for two groups, seronegative and seropositive at the time of study entry. Antigen levels are described as the percentage negative, < 3 ng/L (the lower level of quantification), at each follow-up timepoint. Treatment differences in the percentage negative at days 1, 3, and 5 are summarized. Additional analyses describing antibody and antigen levels are described in the supplemental appendix.

Time to sustained recovery was analyzed with a Fine-Gray model to account for the competing risk of death, stratified by trial pharmacy. Recovery rate ratios (RRRs) and 95% CIs are reported; RRRs >1.0 favor bamlanivimab. To address the stated *a priori* hypotheses, the heterogeneity of RRRs for sustained recovery across subgroups defined by antibody level (negative versus positive for nAb at entry) and dichotomized by median levels of viral replication markers (plasma antigen (< 1000, the approximate median, versus ≥ 1000 ng/L) or viral RNA (<10,000, the approximate median, versus ≥ 10,000 copies/mL)), and combinations of these subgroups were assessed by including interaction terms between treatment and subgroup indicators in expanded Fine-Gray regression models. These subgroup analyses for sustained recovery are described further in the supplemental appendix.

Subgroups defined by nAb were also examined for the percentage plasma antigen negative at days 1, 3 and 5 and for safety outcomes. Methods are given in the supplemental appendix.

Statistical analyses were performed using SAS version 9.4 (SAS Institute, Cary, NC) and R version 4.0 (R Foundation for Statistical Computing)^18^. P-values are two-sided, unless noted. The statistical analysis plan, including the supplemental plan developed for the measurements made on specimens, are included as appendices.

## Results

### Study Participants

Of the 326 patients randomized between August 5 and October 13, 2021, 314 were infused and are included in the analysis cohort. (**Figure S2**); 163 received bamlanivimab and 151 received placebo.

Characteristics at study entry, which were previously reported^17^, and central laboratory measurements on stored samples are shown in **Table 1**. Median (IQR) age was 61 (49, 71) years; median days from symptom onset were 7 (5, 9); and 73% were receiving oxygen, most by a standard nasal cannula. Most patients had elevated IL-6, CRP, and D-dimer levels: medians (IQRs) were 6.7 (2.7, 15.0) ng/L, 137 (39, 544) mg/L, and 0.90 (0.63, 1.38) mg/L, respectively. At enrolment, 50% had positive endogenous neutralizing antibodies (nAbs), and 59% had antibodies against nucleocapsid (anti-N Abs); 66% were positive for both. At enrolment, 95% had plasma antigen levels ≥ 3ng/L, and 50% had antigen levels ≥ 1,000 ng/L.

**Table 1.** Participant characteristics at time of randomization.

| Baseline Characteristics |  | Bamlanivimab<br>(n=163 ) | Placebo<br>(n=151 ) | Total<br>(n=314 ) |
| --- | --- | --- | --- | --- |
| Age | Median (IQR) - yr | 63 (50, 72) | 59 (48, 71) | 61 (49, 71) |
| Female sex | No. (%) | 66 (40%) | 71 (47%) | 137 (44%) |
| Pre-COVID-19 residence: | No. (%) |  |  |  |
| independent/community dwelling |  | 157 (96%) | 142 (94%) | 299 (95%) |
| Race/ethnicity | No. (%) |  |  |  |
| White |  | 76 (47%) | 71 (47%) | 147 (47%) |
| Hispanic |  | 41 (25%) | 33 (22%) | 74 (24%) |
| Black |  | 33 (20%) | 34 (23%) | 67 (21%) |
| Other |  | 13 (8%) | 13 (9%) | 26 (8%) |
| Body mass index; | No. (%) |  |  |  |
| ≥ 30 |  | 81 (50%) | 83 (55%) | 164 (52%) |
| ≥ 40 |  | 20 (12%) | 22 (15%) | 42 (13%) |
| History of: | No. (%) |  |  |  |
| Hypertension requiring medication |  | 84 (52%) | 74 (49%) | 158 (50%) |
| Diabetes requiring medication |  | 54 (33%) | 36 (24%) | 90 (29%) |
| Renal impairment |  | 24 (15%) | 9 (6%) | 33 (11%) |
| Asthma and/or COPD |  | 23 (14%) | 22 (15%) | 45 (14%) |
| At least one co-morbidity |  | 117 (72%) | 100 (66%) | 217 (69%) |
| Days since symptom onset | Median (IQR) | 7 (5, 9) | 8 (5, 9) | 7 (5, 9) |
| Current use of: | No. (%) |  |  |  |
| Remdesivir |  | 60 (37%) | 66 (44%) | 126 (40%) |
| Antibacterial |  | 57 (35%) | 36 (24%) | 93 (30%) |
| Glucocorticoid |  | 85 (52%) | 74 (49%) | 159 (51%) |
| Antiplatelets/anticoagulation |  | 110 (67%) | 97 (64%) | 207 (66%) |
| ACE inhibitor or ARB |  | 41 (25%) | 31 (21%) | 72 (23%) |
| NSAID |  | 17 (10%) | 16 (11%) | 33 (11%) |
| Oxygen Requirement | No. (%) |  |  |  |
| Not receiving supplemental oxygen |  | 44 (27%) | 42 (28%) | 86 (27%) |
| Supplemental oxygen < 4 L/min |  | 60 (37%) | 56 (37%) | 116 (37%) |
| Supplemental oxygen ≥ 4 L/min |  | 29 (18%) | 35 (23%) | 64 (20%) |
| Non-invasive ventilation or HFNC |  | 30 (18%) | 18 (12%) | 48 (15%) |
| Invasive ventilation or ECMO |  | 0 (0%) | 0 (0%) | 0 (0%) |
| Laboratory assessments |  |  |  |  |
| Plasma nucleocapsid antigen | Median (IQR) - ng/L | 1240 (141, 3578) | 856 (138, 3400) | 1037 (141, 3578) |
| ≥ 3 ng/L (positive) | No. (%) | 148 (94%) | 142 (96%) | 290 (95%) |
| Nasal swab fluid viral RNA | No. (%) positive | 120 (78%) | 118 (82%) | 238 (80%) |
| Viral load if RNA positive | Median (IQR) copies/mL | 30742<br>(2706, 313397) | 31411<br>(1177, 376109) | 30742<br>(1466, 340338) |
| Neutralizing antibodies (nAb) | No. (%) positive | 83 (53%) | 69 (47%) | 152 (50%) |
| Anti-nucleocapsid (N) antibodies | No. (%) positive | 93 (59%) | 87 (59%) | 180 (59%) |
| Interleukin-6<sup>+</sup> | Median (IQR) - ng/L | 7.6 (3.2, 16.2) | 5.7 (2.4, 14.8) | 6.7 (2.7, 15.0) |
| D-dimer<sup>+</sup> | Median (IQR) - mg/L | 0.95 (0.64, 1.38) | 0.86 (0.62, 1.38) | 0.90 (0.63, 1.38) |
| C-reactive protein<sup>+</sup> | Median (IQR) - mg/L | 153 (37, 602) | 130 (40, 492) | 137 (39, 544) |
| B-Lymphocytes<sup>++</sup> | Median (IQR) - 10<sup>9</sup> | 0.80 (0.56, 1.07) | 0.81 (0.55, 1.31) | 0.80 (0.55, 1.12) |

+ Upper limit of normal for Interleukin-6 (1.8 pg/ml), D-dimer (0.5 µg/mL) and CRP (10 µg/mL).
++ Lower limit of normal for B-lymphocytes is 1 10<sup>9</sup>/L.

Among 298 participants with available nasal swab material at entry, viral RNA was detected in 238 (80%); median (IQR) RNA viral level among those RNA positives was 30,742 (1466, 340338) copies/mL.

Viral RNA sequences from 255 participants (average depth: 4393; 204 with 75% or more genome coverage; Figure S3) identified no concerning mutations in codons 417, 452 or 484 in the spike protein; virus from six participants had deletions in codon 69-70. All genomes contained the D614G mutation (i.e. the B strain), and one person had strain B.1.1.7 (i.e. alpha variant).

Characteristics according to nAb and anti-N Ab status at entry are given in **Tables S2-S4**.

### Antibody Changes through 5 Days of Follow-up

The percentage with positive nAbs following infusion at day 1, 3, and 5 are summarized for all patients (Figure 1A) and for patients seronegative (Figure 1B) and seropositive (Figure 1C) for nAbs at entry. Treatment differences are more pronounced for patients who were negative at entry (**Figure 1B**). As anticipated, nearly all (97%) seronegative patients given the neutralizing monoclonal antibody bamlanivimab had nABs on day 1 post infusion, compared with 32% on placebo (p<0.001). Subsequently, the placebo group progressively seroconverted. In the group with positive nAbs at entry, little difference between treatment groups (**Figure 1C**). A summary of nAb titres, instead of percentages, demonstrated similar patterns (**Figure S4**).

**Figure 1.**
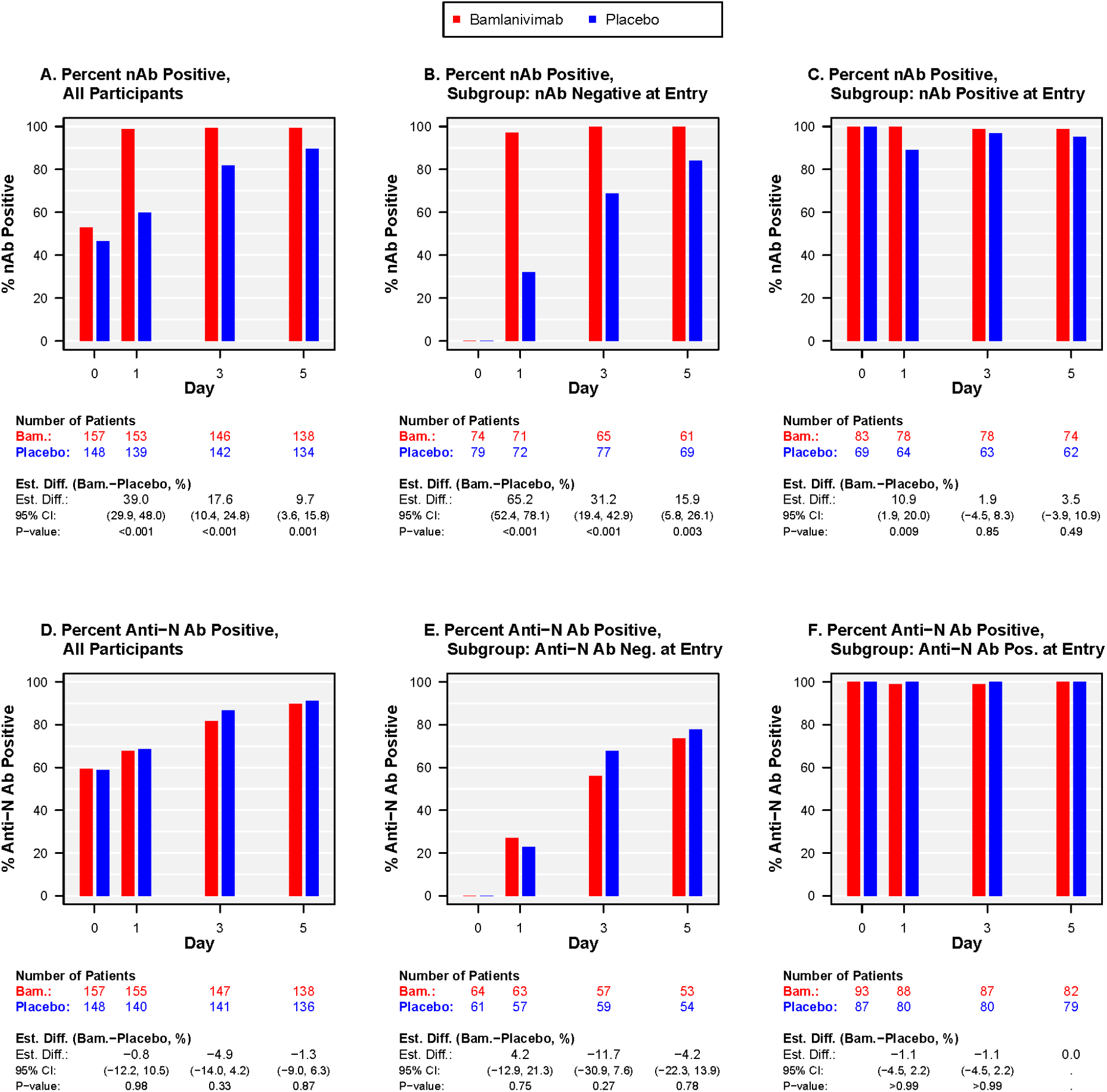
Percent positive of IgG antibodies against the receptor-binding domain of SARS-CoV-2 spike protein (nAb)(top row, A-C) and total antibodies (IgG/A/M) against the nucleocapsid antigen (anti-N Ab)(bottom row, D-F), overall (left column, A, D), and according to antibody status at entry: (middle column, B, E) negative and (right column C, F) positive.

Overall, and for the subgroups with anti-N Abs at entry, the percentage positive for anti-N Abs on days 1, 3 and 5 did not vary between the two treatment groups (**Figures 1D-F**). Findings were similar when levels were considered (**Figure S5**).

### Plasma Antigen Changes, Overall and by nAb Status, through 5 Days of Follow-up

Percentage with plasma antigen levels < 3 ng/L at day 5 comprised 44% and 43% of the bamlanivimab and placebo groups, respectively (p=0.95); the percentage with levels < 1000 ng/L at day 5 was 98% for both the bamlanivimab and placebo groups (**Figure 2** and **Table S5**). Fewer patients in both treatment groups achieved antigen levels < 3 ng/L at day 5, if they were nAbs negative at entry (**Figure 2B**; 26% and 27%, respectively) compared to nAb positive (**Figure 2C**; 57% and 58%, respectively).

**Figure 2.**
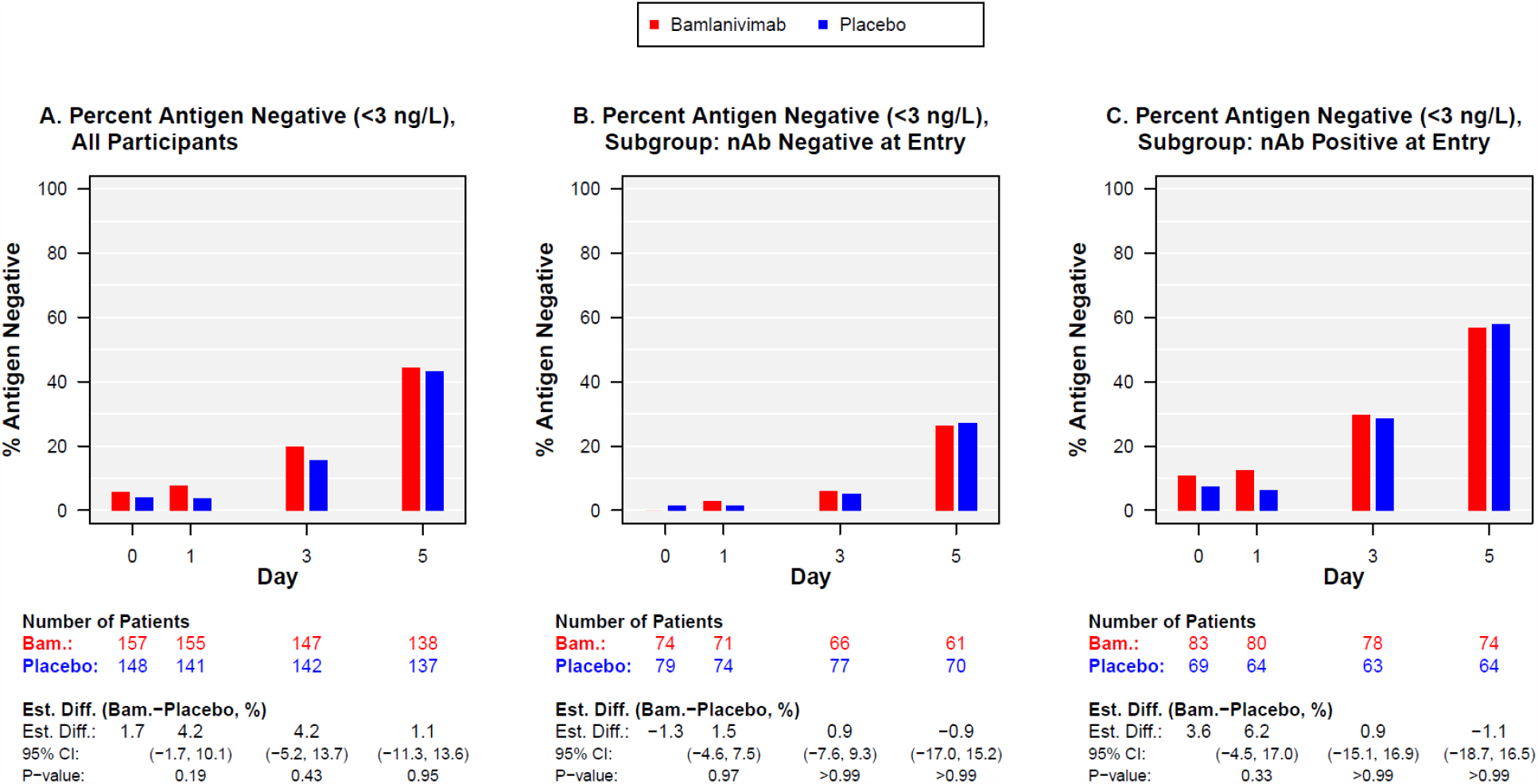
Percentage with plasma antigen below the level of quantification (3 ng/L) at days 0 to 5 for those randomised to bamlanivimab and placebo, overall (A), and according to neutralizing antibody (nAb) status at entry: (B) negative and (C) positive. ^+^ ^+^ Table S6 and Figure S5 display plasma antigen levels.

Log10_10_-transformed antigen levels are summarized overall, and by nAb status at entry in the supplemental appendix (**Figure S6**).

### Sustained Recovery

Sustained recovery by day 90 was achieved by 144 patients (88%) in the bamlanivimab group and 136 patients (90%) in the placebo group; RRR=0.99 (95% CI:0.79-1.22; p=0.89) (**Figure 3A**); median days to sustained recovery was 19 days and sustained recovery at day 28 was 80% for each treatment group.

**Figure 3:**
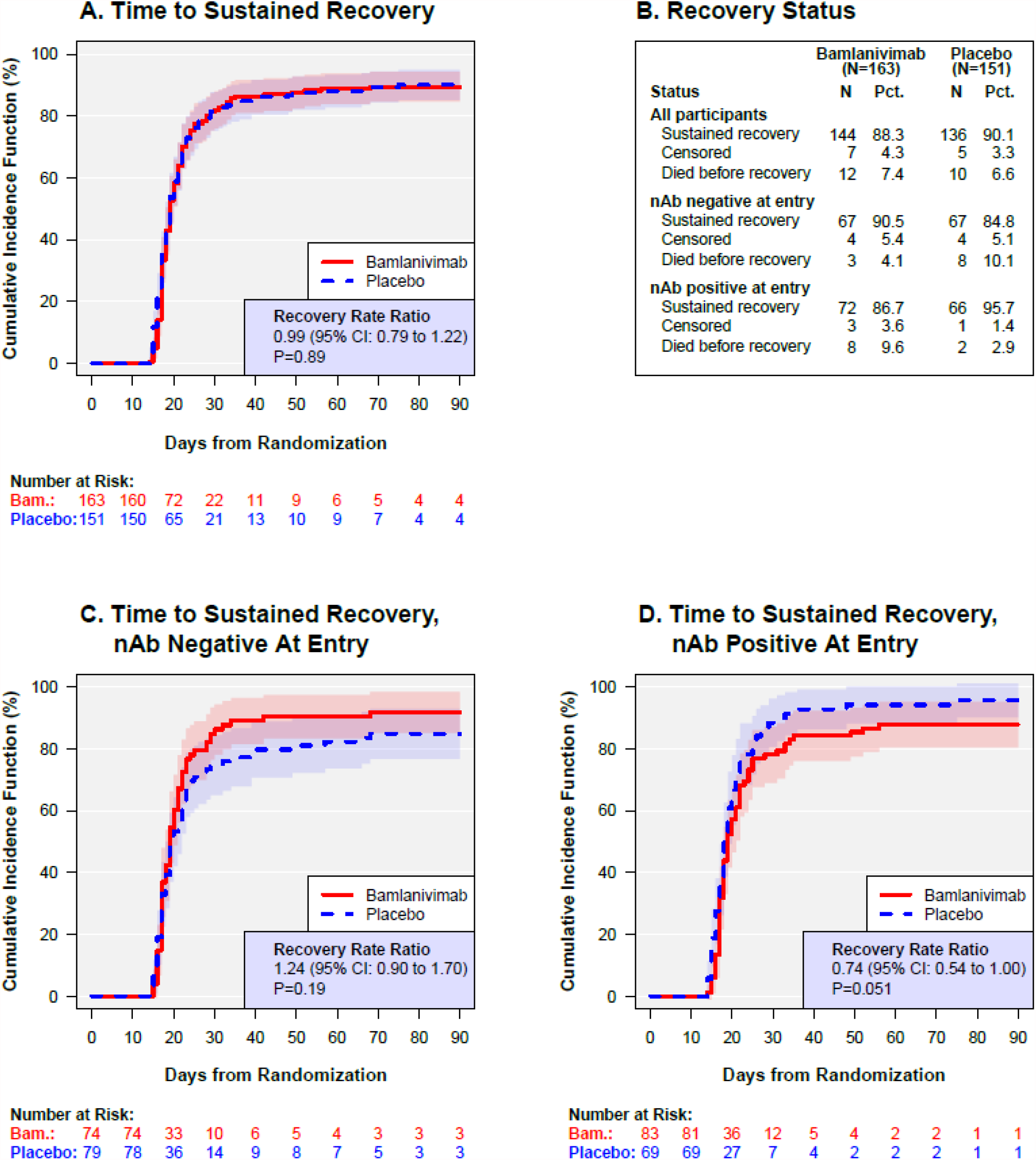
Sustained recovery for Bamlanivimab versus placebo: overall (A), and according to neutralizing antibody (nAb) status at entry: (C) negative and (D) positive. ^+^ (B) summarizes the recovery status at day 90 for cohorts displayed in (A), (C) and (D). ^+^The hazard ratios in (C) were different from those in (D) (p=0.02 for difference). See Figures 4, and S7-S9 for further subgroupings.

Among patients nAbs negative and positive at entry, the RRRs were 1.24 (95% CI: 0.90-1.70) and 0.74 (95% CI: 0.54-1.00), respectively (p=0.02 for difference) (**Figures 3C, 3D, and 4**). For nAb negative patients, the rate of sustained recovery was 91% for the bamlanivimab group and 85% for the placebo group; these percentages were 87% and 96%, respectively for those who were nAb positive (**Figure 3B**).

**Figure 4.**
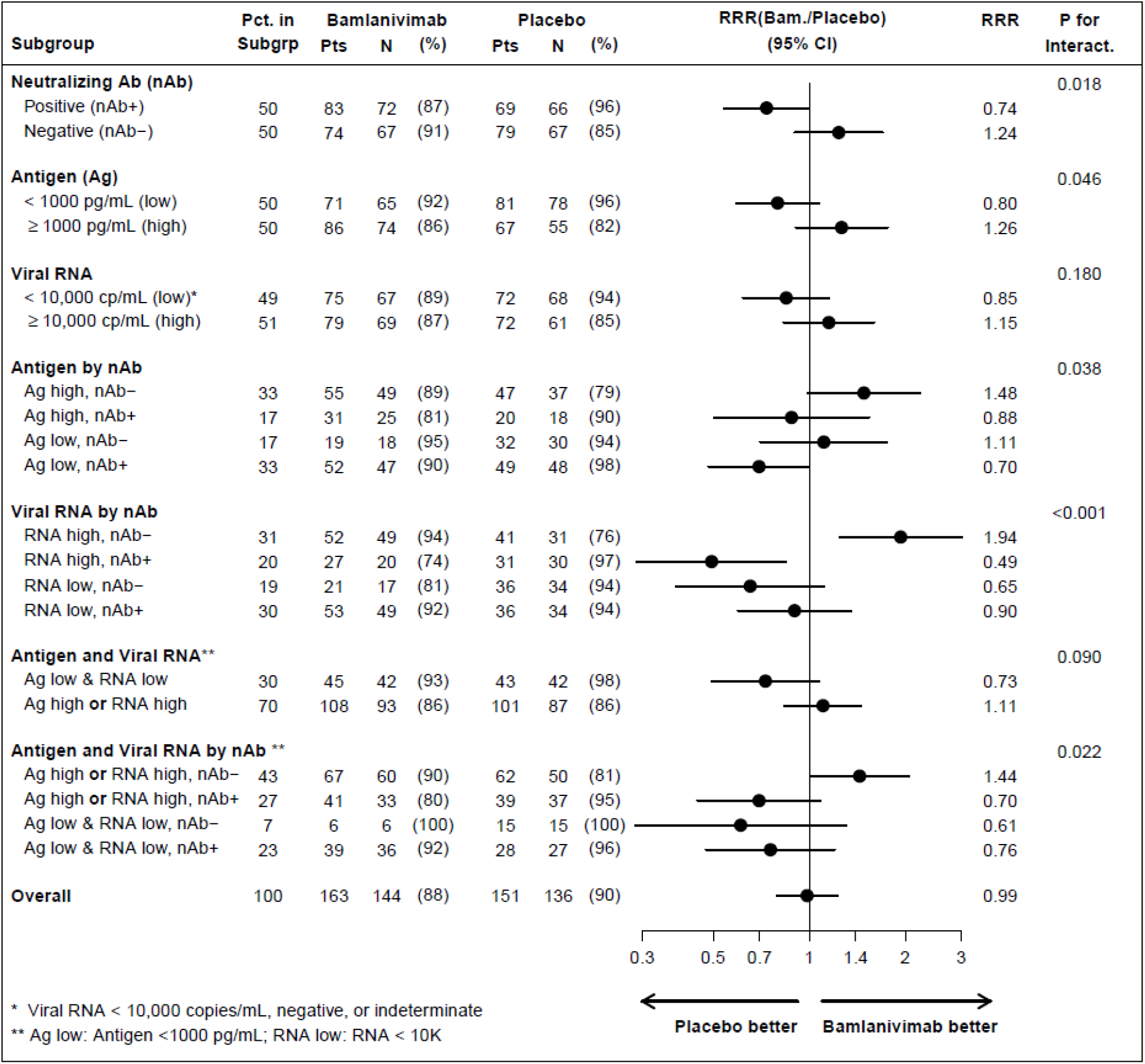
Sustained recovery according to subgroups at study entry: nAb status and levels of viral measures (i.e. plasma antigen and nasal viral RNA)^+^ ^+^See Figures S7-S9 for other subgroupings.

We next compared sustained recovery in subgroups according to median entry levels of plasma antigen and viral RNA, and then, further subdividing these subgroups, according to whether they had nAbs at entry. As hypothesized, among those who were nAb negative, the difference between bamlanivimab and placebo were more evident if plasma antigen or nasal-swab viral RNA were above the median entry levels (RRRs=1.48; 95% CI: 0.99-2.23 and 1.94; 95% CI: 1.25-3.00, respectively). The interaction p-values were 0.04 and < 0.001, respectively, reflecting heterogeneity among the four subgroups defined by nAb and viral level (**Figure 4**). For the subgroup considering presence of either elevated nasal RNA levels or elevated plasma antigen who were nAb negatives, the RRR was 1.44 (95% CI: 1.01-2.06) (**Figure 4**).

Other subgroups for sustained recovery, including those that considered demographic, clinical factors, anti-N antibody levels at baseline, are summarized in **Figures S7, S8, and S9**.

### Safety Outcomes

Safety outcomes are described below and summarized in **Figure 5, Tables S6-S8**.

**Figure 5.**
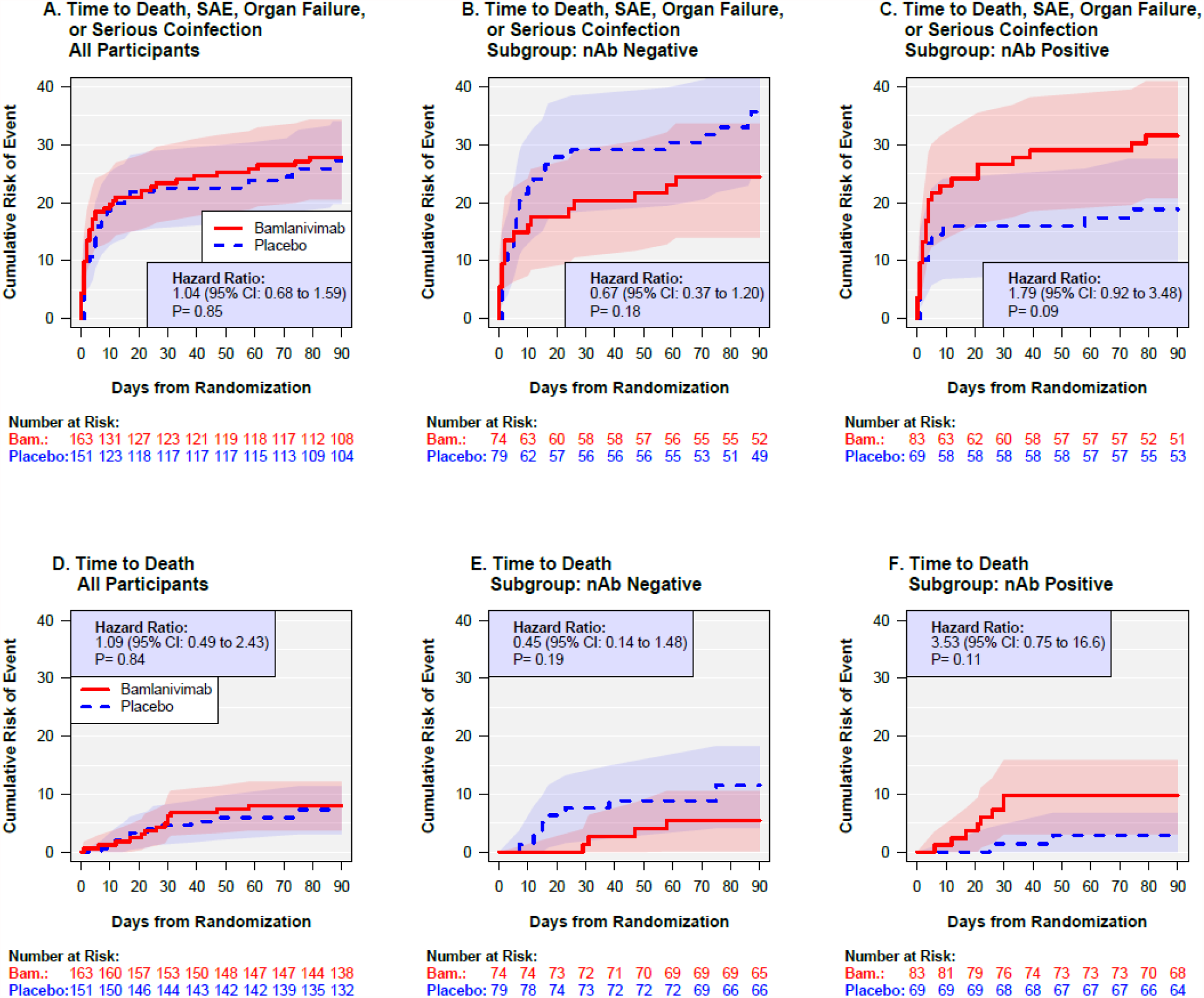
A composite safety outcome (death, serious adverse event (SAE), organ failure or serious infection) (top row, A-C) and death (bottom row, D-F) for bamlanivimab versus placebo: overall (left column, A, D), and according to neutralizing antibody (nAb) status at entry: (middle column, B, E) negative and (right column C, F) positive. ^+^The hazard ratios in (B) were different from those in (C) (p=0.03 for difference), as were the those in (E) versus (F) (p=0.04). See table S7 for overview of all predefined safety outcomes.

By day 5, the composite outcome of mortality, organ failure, serious infections, serious adverse events (SAEs), or grade 3 or 4 adverse events occurred in 45 patients (28%) in the bamlanivimab group and 28 (19%) patients in the placebo group (OR=1.83 (95% CI: 1.01-3.29); p=0.05). By day 28, 52 patients (32%) in the bamlanivimab group and 42 patients (28%) in the placebo group had developed this composite outcome (p=0.35) (**Table S6**). Through day 90, a composite of death, organ failure, serious infection or SAEs occurred in 45 (28%) patients in the bamlanivimab group and in 41 patients (27%) in the placebo group (p=0.85) (**Figure 5A** and **Table S6**). Most of the events at day 90 were due to organ failure (68 events) (Table S8); most were respiratory failure (41 of 68 events). Through day 90, 13 deaths (8.0%) occurred in the bamlanivimab group and 11 deaths (7%) occurred in the placebo group (hazard ratio = 1.09; 95% CI: 0.49-2.43) (Table S6 and **Figure 5D)**. Twenty-two of the 24 patients who died did so before sustained recovery (see Figure 3B).

For the day 90 composite safety outcome, the HRs (bamlanivimab versus placebo) for those nAb negative and positive at entry, were 0.67 (**Figure 5B**) and 1.8 (**Figure 5C**), respectively, (p=0.03 for difference) (**Tables S7**). For death, these HRs were 0.45 (**Figure 5D**) and 3.5 (**Figure 5E**) (p=0.04 for difference). Treatment differences for those who were nAb positive and nAb negative at entry were evident for each of the components of the day 90 composite safety outcome (**Table S8**).

### Inflammatory and Coagulation Markers through 5 Days of Follow-up

Average log_10_ transformed levels of IL-6 (**Figure S10A-C**) and CRP (**Figure S10D-F**) declined in both treatment groups from entry through Day 5, overall and for subgroups defined by nAb at entry. The decline in D-dimer for both treatment groups was more modest than for IL-6 and CRP (**Figure S11**). Overall and for each subgroup the differences by day of follow-up did not differ between the treatment groups.

## Discussion

In support of a pre-specified hypothesis, we found that treatment benefit from bamlanivimab compared to placebo differed according to the presence of neutralizing antibodies at study entry, with possible benefit in seronegative patients with high plasma antigen or high nasal viral RNA levels. Additionally, in patients already having mounted an endogenous antibody response, administration of bamlanivimab may cause harm. These data need urgent confirmation, as they may have potential impact on routine clinical administration of passive immunotherapy to patients with COVID-19 and our future research agenda.

Overall, we found that the chance of sustained recovery was comparable for those randomized to bamlanivimab versus placebo. This contrasts with the benefit observed from using various neutralizing monoclonal antibody preparations in early COVID-19^7-10,19,20^. Before reviewing unblinded laboratory results, we considered whether this null-effect was due to enrolling patients who had already started to mount an effective neutralizing antibody response, compared to patients earlier in the course of their infection. Therefore, infusion of an additional exogenous neutralizing antibody would be redundant in hospitalized patients.

In support of such an explanation, we found a positive trend for the bamlanivimab arm among the subgroup of patients (50% of total) without nAbs at study entry and a trend in the opposite direction for those with nAbs (p for subgroup by treatment interaction =0.02). A similar pattern was observed across subgroups according to total nucleocapsid antigen antibodies. The limited sample size of the study does not allow firm conclusions based on these findings, which should be reproduced in independent trials assessing other types of passive immune therapies in the same patient setting.

Consistent with these observations, a recent preprint reported preliminary data comparing open label use of two neutralizing monoclonal antibodies (casirivimab and imdevimab) versus usual care from the RECOVERY study. A reduced risk of death among hospitalized patients entering the study seronegative for anti-SARS-CoV-2 antibodies was found, but not overall or for those seropositive at entry^16^. Those results and ours support using serological screening at hospital admission to identify the subgroup with a possible benefit.

In RECOVERY, remdesivir was not standard-of-care; it was in our trial. Here, most participants had COVID-19 pneumonia, but none had pulmonary failure requiring mechanical ventilation. The population not requiring invasive ventilation was the one where remdesivir, in an earlier randomized-placebo-controlled trial^21^, showed clinical benefit. Of note, 28-day mortality in the usual care arm of RECOVERY was 21%; in our trial placebo group mortality after 90 days was 7%. Regardless, given the two trials’ consistent findings by antibody status, possible benefit from neutralizing monoclonal antibodies may be expected whether or not combined with remdesivir.

Neutralizing monoclonal antibodies function as antiviral agents. Identification of patients in whom SARS-CoV-2 is replicating and advancing the disease course should be the goal—this is the population where benefit seems likely. We assessed two viral replication markers, plasma antigen and nasal-swab viral RNA levels. The correlation between plasma antigen and nasal viral RNA levels at entry was minimal. IL-6 levels, known to be increased in a variety of viral infections^22,23^, paralleled with plasma antigen levels at baseline, but not with levels of nasal viral RNA. This suggests that viral replication in the nasal cavity and elsewhere in the body is partly uncoupled.

Higher levels of either or both viral replication markers at entry were associated with reduced chance of sustained recovery from the disease; this confirms findings from several smaller studies assessing viral markers and outcomes^24-27^. In support of our *a priori* hypothesis, higher levels of either viral marker, among patients not yet having mounted endogenous neutralizing antibodies at entry may identify a subgroup for which there is clinical utility of bamlanivimab.

Plasma antigen levels decreased rapidly over time and in 40% of the population were cleared after 5 days of follow-up; the clearance rate was comparable between the two arms of the study, providing some reassurance that bamlanivimab does not lead to enhanced viral replication.

The neutralising effect of bamlanivimab is markedly reduced by certain mutations (including those encoding residues K417N/T, L452R and E484K) in the spike protein^28,29^. However, our data show that these viral variants are not present in this patient cohort. The cohort was recruited between August and October 2020, before escape variants with these signature mutations were widely observed. Deep sequencing of 255 viral isolates retrieved at study entry failed to identify these mutations.

Infusion of high concentrations of neutralizing antibodies, as was done in this trial, may have led to an undesired and harmful inflammatory reaction (e.g. type III hypersensitivity -forming of antibody-antigen complexes) in some patients, which may have nullified our ability to find a beneficial effect of the intervention here. We cannot verify this possibility, but among participants with endogenous neutralizing antibody production, and hence a low chance of benefit from infused neutralizing antibodies, the effect estimates of sustained recovery and overall risk of death favored placebo. A mortality trend among seropositive persons favoring standard-of-care was also seen in the RECOVERY trial^16^. Hence, infused neutralizing antibodies may cause harm in hospitalized patients already having mounted an endogenous neutralizing antibody response.

Further insight into the potential causes of such a potential harmful effect from passive immunity is urgently needed. Trajectories of three host biomarkers, IL-6, CRP and D-dimer, though the first 5 days of follow-up, did not differ significantly according to randomized treatment. However, the limited sample size does not allow us to exclude an adverse inflammatory effect from bamlanivimab overall and, more specifically, to those patients with endogenous antibody production. Additional studies of these and other biomarkers in larger cohorts assessing passive immunotherapy, is underway.

The TICO platform will proceed with clinical evaluation of additional COVID-19 treatments. Two other neutralizing antibody agents, VIR-7831 and Brii Bio 196/198, failed TICO’s futility assessment^30^. A fourth neutralizing antibody product (AZD7442)^31^, genetically modified to remove binding to FcγR and complement proteins, is currently under late stage evaluation. Those cohorts are now being characterized for antibody and antigen status at entry in order to produce similar analyses to those reported here.

Disclosure forms provided by the authors are available with the full text of this article at NEJM.org.

## Data Availability

The data set used for this manuscript is available upon request to the INSIGHT scientific steering committee at

## Acknowledgements

We would like to acknowledge the independent DSMB of TICO for their constructive review of our protocol and for their thoughtful guidance based on interim reviews of data. Members of the DSMB are Merlin L. Robb, MD (chair), David Glidden, PhD, Graeme A. Meintjes, MBChB, PhD, Barbara E. Murray, MD, Stuart Campbell Ray, MD, Valeria Cavalcanti Rolla, MD, PhD, Haroon Saloojee, MB.BCh, FCPaed, MSc, Anastasios A. Tsiatis, PhD, Paul A. Volberding, MD, and Jonathan Kimmelman, PhD, and the Executive Secretary is Sally Hunsberger, PhD.

