## Supplemental Appendix for "Clinical and Virological Response to a Neutralizing Monoclonal Antibody for Hospitalized Patients with COVID-19"

#### Section 1: TICO Study Group

**Writing group affiliations**

The affiliations of the members of the writing group are as follows: CHIP Centre of Excellence for Health, Immunity, and Infections, Department of Infectious Diseases, Rigshospitalet, Copenhagen, Denmark (J.D.L. and D.D.M.), Division of Biostatistics, School of Public Health, University of Minnesota, Minneapolis (B.G.,T.A.M, C.R., S.S., D.W., J.D.N.), Division of Pulmonary, Allergy, and Critical Care Medicine (C.E.B.) and Division of Infectious Diseases (T.L.H.), Department of Medicine, Duke University, Durham, Baylor University Medical Center, Dallas (R.L.G., U.S.), Intermountain Medical Center, Murray (S.M.B.), University of Utah, Salt Lake City (S.M.B., E.S.H., K.U.K.), Department of Emergency Medicine, Vanderbilt University Medical Center, Nashville (W.H.S), Department of Internal Medicine, Section on Pulmonary, Critical Care, Allergy, and Immunology, Wake Forest School of Medicine, Winston-Salem (D.C.F.), UT Southwestern Medical Center, Dallas (M.K.J), Department of Infectious Diseases, Copenhagen University Hospital, Amager and Hvidovre, Denmark (T.B), Department of Surgery, Keck School of Medicine, University of Southern California, Los Angeles (M.E.B.), Division of Cardiothoracic Surgery Emory University School of Medicine, Atlanta (B.G.L.), Hennepin Healthcare Research Institute, Minneapolis and University of Minnesota (J.V.B.), Department of Internal Medicine, Respiratory Medicine Section, Herlev and Gentofte Hospital, University of Copenhagen, Hellerup, Denmark and Department of Infectious Diseases, Rigshospitalet, Copenhagen, Denmark (J.-U.J.), Denver Public Health, Denver Health and Hospital Authority, Denver (E.M.G.), Department of Emergency Medicine, University of Colorado School of Medicine, Aurora (A.A.G.) Department of Infectious Diseases, Odense University Hospital, Odense (I.S.J.), Department of Infectious Diseases, Henry Ford Hospital, Detroit (N.M.), Department of Medicine, Department of Anesthesia, The University of California, San Francisco (M.A.M.), Aarhus University Hospital, Skejby (L.Ø.), The Kirby Institute, University of New South Wales, Sydney (C.C.C., M.N.P.), U.S. Department of Veterans Affairs, Washington, D.C. (V.J.D.), Medical Research Council Clinical Trials Unit at UCL, University College London (A.G., A.G.B., M.K.B.P.), Guy's & St. Thomas' NHS Foundation Trust, London (A.G.), National Institute of Allergy and Infectious Diseases, Bethesda (E.S.H., K.S.S., H.C.L.), Infectious Diseases Department & irsiCaixa AIDS Research Institute, Hospital Universitari Germans Trias i Pujol, Catalonia (R.P.), Institute for Global Health, University College London (A.N.P.), Leidos Biomedical Research, Inc., Frederick (R.L.D., M.T., H.H.), Advanced Biomedical Laboratories, LLC., Cinnaminson (N.P.G.), Laboratory of Human Retrovirology and Immunoinformatics (W.C. and B.T.S.), Laboratory of Molecular Cell Biology (V.N.) and AIDS Monitoring Laboratory (A.W.R, H.H.), all Frederick National Laboratory for Cancer Research, Frederick, Gilead Sciences, Foster City (H.C.), Eli Lilly and Company, Indianapolis (P.K.), Department of Population Health Science and Policy, Icahn School of Medicine at Mount Sinai, New York (A.C.G.), Veteran Affairs Medical Center, Washington, D.C. (V.L.K.), George Washington University School of Medicine and Health Sciences, Washington (V.L.K.), St Vincent's Hospital, Sydney (M.N.P.), Division of Pulmonary and Critical Care, Department of Medicine, Massachusetts General Hospital, Boston (B.T.T.), Harvard Medical School Boston (B.T.T.)

U.S. National Institute of Allergy and Infectious Diseases incl. Department of Clinical Research. H. Clifford Lane, *M.D.,* John Tierney, *B.Sc.N.*, Elizabeth Higgs, *M.D., M.P.M.,* Kevin Barrett, *R.N., B.Sc.N.,* Betsey R. Herpin, *M.Sc.N., C.C.R.C., R.N.,* Mary C. Smolskis, *B.Sc.N., M.A.,* Susan E. Vogel, *R.N., B.Sc.N.,* Laura A. McNay, *M.Sc.,* Kelly Cahill, *R.N., M.Sc., C.C.R.C., R.A.C.,* Page Crew, Pharm.D., *M.P.H., B.C.P.S.,* Matthew Kirchoff, *Pharm.D., M.Sc., M.B.A.,* Ratna Sardana, *B.A.,* Sharon Segal Raim, *M.P.H.,* Kathryn Shaw-Saliba, *Ph.D.*

Frederick National Laboratory for Cancer Research/Leidos Biomedical Research, Inc., R. Baseler, *M.Sc.*, Marc J. Teitelbaum, *M.D., M.Sc.*, Shelly M. Simpson, *M.Sc.*, Molly J. Buehn, *M.Sc.*, David Vallée, *Pharm.D., M.P.H.*, Vanessa Eccard-Koons, *M.Sc.*, Stacey Kopka, *M.Sc.*, Theresa M. Engel, *M.F.S.*, Cynthia Osborne, *B.Sc.*, Leah Giambarresi MacDonald, *M.Sc., R.A.C.*, Liam M. Harmon, *C.C.R.A.,* Denise M. Shelley, *M.Sc.,* Mi Ha Kim, Ph.D., *C.C.R.P.*, Joy Beeler-Knights, *M.P.H., C.C.R.A., A.C.R.P.-C.P.M.*, Lindsey Yeon, Robin L. Dewar, *Ph.D.,* Ven Natarajan, *Ph.D.*, Weizhong Chang, *Ph.D.*, Brad T. Sherman, *M.S.*, Adam W. Rupert, *BS*, *MT(ASCP),* Helene Highbarger, *M.S.,* Michael Baseler, *Ph.D.,* Perrine Lallemand, *Ph.D..,* Tauseef Rehman, *Ph.D.,* Danielle Lynam, *Ph.D.,* Gregg Mendez, *Ph.D..,* Tom Imamichi, *Ph.D.,* Sylvain Laverdure, *Ph.D..,* Sharada Paudel, *Ph.D.,* Kyndal Cook, *M.S.,* Shawn Brown, *M.S.*, Ayub Khan, *Ph.D.,* Allison Hazen, *M.S.,* Yunden Badralmaa, *M.S.*.

INSIGHT SDMC, Division of Biostatistics, School of Public Health and School of Statistics, University of Minnesota, Minneapolis, MN, USA. James D. Neaton, Ph.D., Laura Amos, Anita Carter, Gary Collins, *M.S.,* Bionca Davis, *M.P.H.,* Eileen Denning, *M.P.H.,* Alain DuChene, Nicole Engen, *M.S.,* Greg Grandits, *M.S.,* Birgit Grund, *Ph.D.,* Merrie Harrison, Nancy Hurlbut, Joseph Koopmeiners, *Ph.D.,* Gregg Larson, *M.A.,* Sue Meger, Shweta Sharma Mistry, *M.S.,* Thomas Murray, *Ph.D.,* James D. Neaton, *Ph.D.,* Ray Nelson, *R.N.,* Kien Quan, *M.S.,* Siu Fun Quan, Cavan Reilly, *Ph.D.,* Greg Thompson, David Vock, *Ph.D.,* Deborah Wentworth, *M.P.H.*

CTSN International Coordinating Center. Annetine C. Gelijns, *Ph.D.*, Alan J. Moskowitz, *M.D.*, Emilia Bagiella, *P.hD.*, Ellen Moquete, *R.N., B.Sc.N.*, Karen O'Sullivan, *M.P.H.*, Evan Accardi, *B.A.*, Emily Kinzel, *M.P.H.,* Gabriela Bedoya, Lopa Gupta*, M.P.H.,* Jessica R. Overbey, *Dr.P.H.*, Maria L. Padillia, *M.D.*, Milerva Santos, *M.P.A.*
U.S. National Heart Lung and Blood Institute. Marissa A. Miller, *D.V.M., M.P.H.*, Wendy C. Taddei-Peters, *Ph.D.*
Duke University Hospital. Peter K. Smith, *M.D.*, Christina E. Barkauskas, *M.D.*, Andrew M. Vekstein, *M.D.*, Emily R. Ko, *M.D.*, *Ph.D.*, Tatyana Der, *M.D.*, John Franzone, *M.D.*, Noel Ivey, *M.D.*, Thomas L. Holland, *M.D.*, Kathleen Lane, *B.Sc.N.*, *R.N.*, Andrew Bouffler, *B.Sc.*, Lauren M. McGowan, *B.Sc.*, *E.M.T.B.*, Ben Stallings, Jennifer Stout, *B.Sc.*, Whitney Franz, *B.Sc.*, R.N., Beth McLendon-Arvik, *Pharm.D.*, Beth A. Hollister, *B.Sc.N.*, *R.N.,* Dana M. Giangiacomo
Baylor, Scott and White Health. Uriel Sandkovsky, *M.D.*, M.Sc., Robert L. Gottlieb, *M.D.*, *Ph.D.*, Michael Mack, *M.D.*, Mezgebe Berhe, *M.D.*, *M.P.H.*, Clinton Haley, *M.D.*, *M.P.H.*, Emma Dishner, *M.D.*, *M.P.H.*, Christopher Bettacchi, *M.D.*, Kevin Golden, *M.D.*, Erin Duhaime, *P.A.-C.*, Cedric Spak, *M.D.*, *M.P.H.*, Sarah Burris, Felecia Jones, Samantha Villa, Samantha Wang, Raven Robert, Tanquinisha Coleman, Laura Clariday, Rebecca Baker, Mariana Hurutado, Nazia Iram, Michelle Fresnedo, Allyson Davis, Kiara Leonard, Noelia Ramierez, Jon Thammavong, Krizia Duque, Emma Turner, Aaron Killian, *Pharm.D.*, Adriana Palacious, *PharmD*, Edilia Solis, Janet Jerrow, Matthew Watts, Heather Whitacre, Elizabeth Cothran
University of Southern California. Michael E. Bowdish, *M.D.*, *M.Sc.*, Zea Borok, *M.B., Ch.B.*, Noah Wald-Dickler, *M.D.*, Douglass Hutcheon, *M.D.*, Amytis Towfighi, *M.D.*, May Lee, *M.D.*, Meghan R. Lewis, *M.D.*, Brad Spellberg, *M.D.*, Linda Sher, *M.D.*, Aniket Sharma, *M.D.*, Anna P. Olds, *M.D.*, Chris Justino, *P.A.-C.,* Edward Lozano, *M.D.*, Chris Romero, *C.R.C.*, Janet Leong, *C.R.C.*, Valentina Rodina, *M.D.*, Christine Quesada, *C.R.C.*, Luke Hamilton, Jose Escobar
Emory University. Brad Leshnower, *M.D.*, *F.A.C.S.*, William Bender, *M.D.*, *M.P.H.*, Milad Sharifpour, *M.D.*, *M.Sc.*, Jeffrey Miller, *M.D.*, Kim T. Baio, *R.N.*, *M.Sc.N.*, Mary K. McBride, *R.N.*, *B.Sc.N.*, *M.A.S.*, Michele Fielding, *R.N., B.Sc.N.*, *C.C.R.C.*, Sonya Mathewson, *R.N.*, *B.Sc.N.*, *C.C.R.C.*, Kristina Porte, *B.A.*, Missy Maton, *R.N.*, *B.Sc.N.*, Chari Ponder, *R.N.*, *B.Sc.N.*, Elizabeth Haley, *R.N.*, *B.Sc.N.*, *C.C.R.N.*, Christine Spainhour, *R.N.*, *C.C.R.C.*, Susan Rogers, *R.Ph.*, Derrick Tyler, *C.C.R.P.*
University of Virginia Health Systems. Jeffrey M. Sturek, *M.D.*, *Ph.D.*, Andrew Barros, *M.D.*, *M.Sc.,* Kyle B. Enfield, *M.D.*, *F.C.C.M.*, *F.S.H.E.A.*, China J. Green, *B.Sc.*, *C.C.R.C.*, Rachel M. Simon, *R.N.*, *B.Sc.N.*, *C.C.R.C.*, Kara Thornton, *Pharm.D.*, *M.Ed.*, *C.C.R.P.*
Ochsner Clinic. Patrick E. Parrino, *M.D.*, *F.A.C.S.*, Stephen Spindel, *M.D.*, Aditya Bansal, *M.D.*, Katherine Baumgarten, *M.D.*, *F.A.C.P.*, *F.I.D.S.A.*, Jonathan Hand, *M.D.*, Derek Vonderhaar, *M.D.*, Bobby Nossaman, *M.D.*, Sylvia Laudun, *D.N.P.*, *M.B.A.*, *R.N.*, *CPHQ*, DeAnna Ames, *M.Sc.*, Geralyn Isaac, *Pharm.D.*, Huan Dinh, *Pharm.D.*, Yiling Zheng, *Pharm.D.*, Hunter McDaniel, Nicolle Crovetto, *M.Sc.*, Abby Richardson
West Virginia University. Vinay Badhwar, *M.D.*, J. Awori Hayanga, *M.D.*, *M.P.H.*, Sunil Sharma, *M.D.*, Brian Peppers, *D.O.*, Paul McCarthy, *M.D.*, Troy Krupica, *M.D.*, Arif Sarwari, *M.D.*, *M.Sc.*, *M.B.A.*, Rebecca Reece, *M.D.*, Lisa Fornaresio, *Ph.D.*, Chad Glaze, *M.Sc.*, Raquel Evans, *B.Sc.N.*, *R.N.*, Fang Di, *R.N.*, *M.Sc.N.*, Shawn Carlson, *M.D.*, *M.Sc.*, Tanja Aucremanne, *B.Sc.N.*, *R.N.*, Connie Tennant, *B.Sc.N.*, *R.N.*, Lisa Giblin Sutton, *Pharm.D.*, Sabrina Buterbaugh, *Pharm.D.*, Roger Williams, *C.Ph.T.*, Robin Bunner, *B.Sc.*
University of Maryland. Ronson J. Madathil, *M.D.*, Joseph Rabin, *M.D.*, Andrea Levine, *M.D.*, Kapil Saharia, *M.D.*, Ali Tabatabai, *M.D.*, Christine Lau, *M.D.*, *M.B.A.*, James S. Gammie, *M.D.*, Maya-Loren Peguero, Kimberly McKernan, Matthew Audette, Emily Fleischmann, Kreshta Akbari, *M.Sc.*, Myounghee Lee, *Ph.D.*, *Pharm.D.*, Andrew Chi, *Pharm.D.*, Hanna Salehi, *Pharm.D.*, Alan Pariser, *Pharm.D.*, Phuong Tran Nyguyen, *Pharm.D.*, Jessica Moore, Adrienne Gee, Shelika Vincent
Dartmouth-Hitchcock Medical Center. Richard A. Zuckerman, *M.D.*, *M.P.H.*, Alexander Iribarne, *M.D.*, *M.Sc.*, Sara Metzler, *B.Sc.N.*, *R.N.*, Samantha Shipman, *B.Sc.N.*, *R.N.*, Taylor Caccia, *B.A.*, Haley Johnson, Crystallee Newton, *B.A.*, *C.C.R.C.*, Doug Parr, *Pharm.D.*
Lutheran Medical Group. Vicente Rodriguez, *M.D.*, Gordon Bokhart, *Pharm.D.*, Sharon M. Eichman

PETAL ICC.
Massachusetts General Hospital, Boston, USA. B. Taylor Thompson *M.D.,* Crystal North *M.D.,* Cathryn Oldmixon *R.N.,* Nancy Ringwood *B.Sc.N.*, Laura Fitzgerald *B.A./B.Sc.,* Ariela Muzikansky *R. N., B.A./B.Sc.,* Richard Morse *B.A./B.Sc.*
Johns Hopkins University. Roy G. Brower, *M.D.*
NIH/NHLBI. Lora A., Reineck *M.D., M.S.*, Neil R. Aggarwal, *M.D.*

ALIGNE SCC**.** Lead Investigators, Baystate Medical Center: Jay Steingrub, *M.D.*, Brigham and Women's Hospital: Peter Hou *M.D.*
Baystate Medical Center: Jay S. Steingrub, *M.D.,* Mark A. Tidswell, *M.D.*, Lori-Ann Kozikowski, *R.N., B.Sc.N.*, *C.C.R.N.,* Cynthia Kardos, *R.N., B.Sc.N.*, C.C.R.N., Leslie De Souza

Boston SCC, Lead Investigators. Beth Israel Deaconess Medical Center: Daniel Talmor, *M.D.* and Nathan Shapiro, *M.D.*
University of Mississippi: Alan E. Jones, *M.D.,* James Galbraith, *M.D.,* Utsav Nandi, *M.D.,* Rebekah Peacock, *R.N.*
Massachusetts General Hospital: Michael R. Filbin, *M.D., M.Sc.,* Kathryn Hibbert, *M.D.,* Blair Alden Parry, *C.C.R.A., B.A.,* Justin D, Margolin, *B.Sc.,* Kelsey Brait, *B.B.A., B.Sc.*

California SCC, Lead Investigators. University of California San Francisco: Michael A. Matthay, *M.D.,* David Geffen School of Medicine at UCLA: Gregory Hendey, *M.D.*
University of California San Francisco, University of San Francisco Mount Zion: Michael A, Matthay, *M.D.,* Kirsten N. Kangelaris, *M.D.,* M.A.S, Kimberly J. Yee, *B.Sc.,* Alejandra E. Jauregui, *B.A.,* Hanjing Zhuo, *M.P.H.*
University of California Fresno: Eyad Almasri, *M.D.*, Alyssa R. Hughes, *B.Sc.,* Rebekah L. Garcia, *C.C.R.P.*, Kinsley A. Hubel, *M.D.*
Stanford University: Angela J. Rogers, *M.D.,* Jennifer G. Wilson, *M.D., M.Sc.,* Rosemary Vojnik, *B.Sc.*, Jonasel Roque *B.Sc.*
University of Texas Health Science Center: Henry Wang, *M.D., M.Sc.,* Ryan M. Huebinger, *M.D.,* Bela Patel, *M.D.,* Elizabeth Vidales, *M.P.H., B.M.Sc.*

Colorado SCC, Lead Investigators. University of Colorado Hospital: Adit A. Ginde, *M.D., M.P.H,* Marc Moss, *M.D.*
University of Colorado Hospital: Adit A. Ginde, *M.D., M.P.H.,* Amiran Baduashvili, *M.D.,* Lakshmi Chauhan, *M.D.,* David J. Douin, *M.D.,* Lani L. Finck, *B.A.,* Carrie Higgins, *R.N.,* Michelle Howell, *R.N.,* Jeffrey McKeehan, *M.Sc.N.,* Marc Moss, *M.D.*
National Jewish Health/St. Joseph Hospital: James H. Finigan, *M.D.,* William Janssen, *M.D.,* Peter Stubenrauch, *M.D.,* Christine Griesmer, *R.N., B.Sc.N., M.P.H.*

Michigan SCC, Lead Investigators**:** University of Michigan: Robert C. Hyzy, *M.D.,* Pauline K. Park, *M.D.*
University of Michigan: Robert C. Hyzy, *M.D.,* Pauline K. Park, *M.D.,* Kristine Nelson, *R.N.,* Kelli McDonough, Jake I. McSparron, *M.D.,* Ivan N. Co, *M.D.,* Bonnie R. Wang, *M.D.,* Shijing Jia, *M.D.,* Barbara Sullins, Sinan Hanna, Norman Olbrich

Montefiore-Sinai SCC, Lead Investigators**:** Montefiore Medical Center**:** Michelle N. Gong, *M.D.,* Mount Sinai Hospital: Lynne D. Richardson, *M.D.*
Montefiore Medical Center Moses, Montefiore Medical Center Weiler: Michelle N. Gong, *M.D.,* M.S, Rahul Nair, *M.D.*
Banner University Medical Center Tucson: Jarrod M. Mosier, *M.D.,* Cameron Hypes, *M.D.,* Elizabeth Salvagio Campbell, *Ph.D.,* Billie Bixby, *M.D.,* Christian Bime, *M.D.,* Sairam Parthasarathy, *M.D.*

Ohio SCC, Lead Investigators: University of Cincinnati**:** R. Duncan Hite, *M.D.,* Ohio State University: Thomas E. Terndrup, *M.D.*, Cleveland Clinic Foundation: Herbert P. Wiedemann, *M.D., M.B.A.*
Cleveland Clinic Foundation, Cleveland Clinic Fairview Hospital, Cleveland Clinic Marymount Hospital: Abhijit Duggal, *M.D.,* Siddharth Dugar, *M.D.,* Debasis Sahoo, *M.D.*, Kiran Ashok, *B.Sc.,* Alexander H. King, *M.Sc.,* Omar Mehkri, M.D.

Pacific Northwest SCC, Lead Investigators**:** Oregon Health and Science University: Catherine L. Hough, *M.D.*, University of Washington Medical Center: Bryce H. Robinson, *M.D.*

Harborview Medical Center, University of Washington Medical Center: Nicholas J. Johnson, *M.D.,* Bryce H. Robinson, *M.D.,* Stephanie J. Gundel, *R.D.,* Sarah C, Katsandres, *B.Sc.*
Oregon Health and Science University: Catherine L. Hough, *M.D.,* Akram Khan, *M.D.*, Olivia F. Krol, Mistry Kinjal, Milad K. Jouzestani
Cedars-Sinai Medical Center: Peter Chen, *M.D.*, Sam S. Torbati, *M.D.,* Yuri Matusov, *M.D.*, June Choe, *M.D.,* Niree A. Hindoyan, *B.Sc.,* Susan E. Jackman, *B.Sc.N., M.Sc*., Emad Bayoumi, *M.B.A.,* Timothy Wynter, *B.Sc.,* Antonina Caudill, *M.P.H., C.P.H.,* Ethan Pascual, *M.A.,* Gregg J. Clapham, *M.A*., Lisa Herrera

Southeast SCC, Lead Investigators**:** Wake Forest Baptist Health: D. Clark Files *M.D.,* Chadwick Miller *M.D.*
Wake Forest Baptist Health: D. Clark Files, *M.D.*, Keven W. Gibbs, *M.D.,* Lori S. Flores, *D.N.P.,* Mary E. LaRose, *R.N.*, *B.Sc.N,* Leigha D. Landreth, *R.N*., *B.S.N*
University of Kentucky: Peter E. Morris, *M.D.,* Jamie L. Sturgill, *Ph.D.,* Evan P. Cassity, *M.Sc.,* Sanjay Dhar, *M.D.,* Ashley A. Montgomery-Yates, *M.D.,* Sara N. Pasha, *M.D.,* Kirby P. Mayer, *Ph.D.*
Virginia Commonwealth University: Marjolein de Wit, M.D., *M.Sc.,* Jessica Mason, *M.P.H*

Utah SCC, Lead Investigators**:** Intermountain Medical Center: Samuel M. Brown, *M.D.,* Joseph Bledsoe, *M.D.*
Intermountain Medical Center: Kirk U. Knowlton, *M.D.,* Samuel Brown, *M.D.,* Michael Lanspa, *M.D.,* Lindsey Leither, *M.D.,* Ithan Pelton, *M.D.,* Brent P. Armbruster, *B.Sc.*, Quinn Montgomery, *B.Sc., AEMT*, Darrin Applegate, *B.Sc.,* Naresh Kumar, *M.P.H., C.C.R.P.,* Melissa Fergus, *B.Sc.,* Katie Brown, *B.Sc., R.N.,* Mardee Merrill, *B.Sc., C.C.R.P.,* Erna Serezlic, *B.Sc.,* Ghazal Palmer, *Pharm.D.,* Brandon Webb, *M.D.,* Valerie T. Aston, *M.B.A., R.R.T., C.C.R.P.*
University of Utah: Estelle S. Harris, *M.D.,* Elizabeth A. Middleton, *M.D.,* Macy A.G. Barrios, *B.Sc.,* Jorden Greer, *B.Sc.,* Amber D. Schmidt, *B.Sc.,* Melissa K. Webb, *Pharm.D*., Robert Paine*, M.D.,* Sean J. Callahan, *M.D.*

Vanderbilt SCC, Lead Investigators**:** Vanderbilt University Medical Center: Wesley H. Self M.D., M.P.H., Todd W. Rice M.D., M.S.C.I.
Vanderbilt University Medical Center: Wesley H. Self M.D., M.P.H., Todd W. Rice M.D., M.S.C.I., Jonathan D. Casey M.D., M.S.C.I., Jakea Johnson M.P.H, Christopher Gray R.N., Margaret Hays R.N., Megan Roth R.N.

INSIGHT Washington ICC, Veterans Affairs (VA) Medical Center, Washington DC: Virginia L. Kan, *M.D.*; Adriana Sánchez, *M.Sc*.; Laura Popielski, *M.P.H.*; Heather Rivasplata, *D.N..P*, *M.P.H.*; Melissa Turner, *M.S.W.*; Michael J. Vjecha, *M.D.*; Amy Weintrob, *M.D.*.
University of Texas Southwestern Medical Center, Dallas, TX: Mamta K. Jain, *M.D.*, *M.P.H.*; Tianna Petersen, *M.Sc*.; Claudia Sanchez Lucas, *M.P.H.*.; Alexis Avery, *PS.M.*; Natalie DellaValle, *Pharm.D*., *B.C.P.S.*; Khanh-Hau Moss, *M.Sc*., *PharmD*.
Hennepin Healthcare Research Institute, Minneapolis, MN: Jason V. Baker, *M.D.*; Jonathan Klaphake, *B.Sc.*; Shari Mackedanz, *R.N.*; Rachael Goldsmith, *B.Sc.*; Hodan Jibrell, *B.Sc.*
Henry Ford Health System, Detroit, MI: Norman Markowitz, *M.D.*, Erika Pastor, *R.N.*;
Mayur Ramesh *M.D.*; Indira Brar, *M.D.*; Emanuel Rivers *M.D.*
Denver Public Health, Denver CO: Edward Gardner, *M.D.*; James Scott, *R.N.*; David Wyles, *M.D.*; Ivor Douglas, *M.D.*; Jason Haukoos, *M.D.*; Mitch Cohen, *M.D.*; Kevin Kamis, *M.P.H.*; Caitlin Robinson, *M.P.H.*
Georgetown University Medical Center, Washington, DC: Princy Kumar, *M.D.*, Maximilian Menna

VA Cooperative Studies Group (CSP) Site Coordinating Center (SCC): Kousick Biswas, *Ph.D.*; Cristin Harrington, *B.A*.
Michael E. DeBakey VA Medical Center, Houston, TX: Barbara W. Trautner, *M.D.*, *Ph.D.;* Lavannya Pandit, *M.D.*, *M.Sc*.; Yigun Wang, *M.A*.
Miami Bruce Carter VA Health Care System, Miami, FL: Paola Lichtenberger, *M.D.*; Gio Baracco, *M.D.*; Carol Ramos, *M.D.*; Lauren Bjork, *Pharm.D.*; Melyssa Sueiro, *M.Sc*.
San Francisco VA Health Care System, San Francisco, CA: Phyllis Tien, *M.D.*; Heather Freasier, *M.Sc*., *R.D.*
Bay Pines VA Healthcare System, Bay Pines, FL: Theresa Buck, *M.D.*; Hafida Nekach, *M.D.*

INSIGHT Copenhagen ICC, CHIP (Centre of Excellence for Health, Immunity and Infections), Department of Infectious Diseases, Rigshospitalet, University of Copenhagen, Copenhagen, Denmark. Jens D. Lundgren, *M.D., D.M.Sc.,* Dorthe Raben, *M.Sc.,* Daniel D. Murray, *Ph.D.,* Lars Peters, *M.D., D.M.Sc.,* Bitten Aagaard, *B.Sc.N.,*

Charlotte B. Nielsen, Katharina Krapp, *Ph.D.,* Bente Rosdahl Nykjær, Katja Lisa Kanne, *M.Sc. B.Sc.N.*, Anne Louise Grevsen, *M.Sc. Dent.,* Zillah Maria Joensen, *B.Sc.N.,* Tina Bruun, *B.Sc.N.*

Denmark.
Copenhagen University Hospital, Amager Hvidovre, Center of Research & Disruption of Infectious Diseases, Department of Infectious Diseases. Thomas Benfield, *M.D.,* Clara Lundetoft Clausen, *M.D.,* Nichlas Hovmand, *M.D.,* Simone Bastrup Israelsen, *M.D.,* Louise Krohn-Dehli, *R.N.,* Cæcilie Leding, *M.D.,* Dorthe Pedersen, *R.N.,* Karen Brorup Pedersen, *M.D.,* Louise Thorlacius-Ussing, *M.D.,* Michaela Tinggaard, *M.D.,* Sandra Tingsgård, *M.D.,* Signe Villadsen, *R.N.*
Herlev-Gentofte Hospital, Respiratory Medicine Section, Department of Internal Medicine. Jens-Ulrik Stæhr Jensen, *M.D., Ph.D.,* Dorthe S. Høgsberg, *R.N.,* Christian P. Rønn, *M.D.,* Ema Rastoder, *M.D.,* Christian Søborg, *M.D., Ph.D.,* Christina Bergsøe, *B.Sc.,* Nuria M.S. Hissabu, *B.Sc.,* Bodil C. Arp, *B.Sc.*
Aarhus Universitetshospital, Skejby, Aarhus. Lars Østergaard, *M.D., Ph.D. D.M.Sc.,* Nina Breinholt Stærke, *M.D.*
Odense University Hospital, Department of Infectious Diseases. Andreas Arnholdt Pedersen, *M.D.,* Inge K Holden, *M.D., Ph.D.*
Dept. of Infectious Diseases, Rigshospitalet, Copenhagen University Hospital. Marie Helleberg, *M.D. Ph.D., D.M.Sc.,* Jan Gerstoft, *MD, D.M.Sc.*
North Zealand University Hospital, Department of Pulmonary and Infectious Diseases. Tomas O. Jensen, *M.D.,* Birgitte Lindegaard, *M.D.*
Kolding Hospital, Department of Medicine. Birgit Thorup Røge *MD. Ph.D.*
Aalborg University Hospital, Department of Infectious Diseases. Henrik Nielsen, *M.D., D.M.Sc.*

Spain.
INSIGHT SCC Spain, Hospital Universitari Germans Trias i Pujol, Badalona. Roger Paredes, *M.D., Ph.D.*, Anna Chamorro, *B.Sc.,* Ariadna Figuerola, *B.Sc.* Maria Exposito, *B.Sc.*
Hospital Universitari Germans Trias i Pujol, Badalona. Roger Paredes, *M.D., Ph.D.,* Lourdes Mateu, *M.D., Ph.D.,* Ana Martínez *B.Sc.,* Adrian Siles *B.Sc.*
Hospital General Universitario Gregorio Marañón, Madrid. Eduardo Fernández-Cruz, *M.D., Ph.D.,* Javier Carbone, *M.D., Ph.D.,* Paco López, *M.D.*
Barcelona Institute for Global Health (ISGlobal), Hospital Clínic - Universitat de Barcelona, Barcelona. José Muñoz, M.D., Ph.D., Daniel Camprubí, M.D. Almudena Legarda, B.VSc.
Hospital Universitario La Paz, IdiPAZ, Madrid. Jose R Arribas, *M.D.*, Alberto Borobia, *M.D. Ph.D.,* Marta Mora-Rillo, *M.D. Ph.D.*, Irene García García, *M.D.*
Hospital Clínico San Carlos, Madrid. Vicente Estrada, *M.D., Ph.D.,* Noemi Cabello, *M.D.,* Maria Jose Nuñez, *M.D.*
Hospital del Mar, Barcelona. Juan P. Horcajada, *M.D., Ph.D.,* Elena Sendra, *M.D.,* Joan Gómez-Junyent, *M.D.*

INSIGHT Sydney ICC, The Kirby Institute, University of New South Wales, Sydney, Australia. Mark Polizzotto, *M.D., Ph.D.,* Gesalit Cabrera, *B.MSc., M.I.P.H,* Catherine Carey, *B.A., M.Sc.,* Christina Chang, *M.D., Ph.D.,* Sally Hough, *B.Sc.,* Sophie Virachit, *B.Sc., Ph.D.,* Amy Zhong, *B.Sc., M.P.H.*

Tan Tock Seng Hospital, NCID, Singapore. *surname underlined, Barnaby E. Young, *M.D., Ph.D.,* Chia Po Ying, *M.D.,* Lee Tau Hong, *M.D.,* Ray J. Lin, *M.D.* , David Lye, *M.D.,* Sean Ong, *M.D.,* M.Med., Puah Ser Hon, *M.D.,* Yeo Tsin Wen, *M.D., Ph.*, Diong Shiau Hui B.Bio, *M.Sc., M.M.Sc.,* Juwinda Ongko, *B.Sc.*

INSIGHT London ICC, MRC Clinical Trials Unit at UC, London, UK. Abdel G. Babiker, *Ph.D.,* Sarah L. Pett, *M.D.,* Fleur Hudson, *B.Sc.*, Mahesh KB Parmar, *Ph.D*, Anna Goodman, *M.D.,* Jonathan Badrock, *B.Sc.,* Adam Gregory, *M.A.*
Greece SCC, National & Kapodistrian University of Athens Medical School. Giota Touloumi, *Ph.D.,* Vicky Gioukari, *B.Sc.*
Uganda SCC, JCRC/MRC/UVRI Uganda Research Unit. Joseph Lutaakome, *M.D.,* Cissy M. Kityo, *M.D.*, Francis Kiweewa, *M.D.*

Eli Lilly and Company, Indianapolis, IN, USA. Paul Klekotka, M.D., Ph.D., Karen Price, Ph.D., Ajay Nirula, M.D., Ph.D.

Gilead Sciences, Foster City, CA, USA. Anu Osinusi, M.D. M.P.H., Huyen Cao, M.D.

Advanced Biomedical Laboratories, LLC., Cinnaminson, NJ, USA. Amanda Kubernac, Bhakti Patel, Kenneth Smith, Robert Kubernac, Norman P. Gerry, *Ph.D.*, Marie L. Hoover, *Ph.D.*

PCI Pharma Services. Craig Brown, Nadine DuChateau, Tris Evans, Adam Flosi, Les Johnson, Amy Treagus, and Christine Wenner.

**FNIH, ACTIV and OWP representative on protocol team**

Stacey J. Adam, *Ph.D.,* Sarah Read, *M.D.,* Eric Hughes, *M.D., Ph.D.*, Rachel H. Harrigan, *M.D.*

#### Section 2: Methods

The complete inclusion and exclusion criteria from the protocol are given below.

**Inclusion Criteria**

- Age ≥ 18 years;
- Informed consent by the patient or the patient's legally-authorized representative
- SARS-CoV-2 infection, documented by PCR or other nucleic acid test (NAT) within 3 days prior to randomization OR documented by NAT more than 3 days prior to randomization AND progressive disease suggestive of ongoing SARS-CoV-2 infection per the responsible investigator;
- Duration of symptoms attributable to COVID-19 ≤ 12 days per the responsible investigator;
- Requiring admission for inpatient hospital acute medical care for clinical manifestations of COVID-19, per the responsible investigator, and NOT for purely public health or quarantine purposes.

**Exclusion Criteria**

- Prior receipt of
  - Any SARS-CoV-2 hIVIG, convalescent plasma from a person who recovered from COVID-19 or
  - SARS-CoV-2 nMAb at any time prior to hospitalization;
- Not willing to abstain from participation in other COVID-19 treatment trials until after Day 5;
- In the opinion of the responsible investigator, any condition for which, participation would not be in the best interest of the participant or that could limit protocol-specified assessments;
- Expected inability to participate in study procedures;
- Women of child-bearing potential who are not already pregnant at study entry and who are unwilling to abstain from sexual intercourse with men or practice appropriate contraception through Day 90 of the study.
- Men who are unwilling to abstain from sexual intercourse with women of child-bearing potential or who are unwilling to use barrier contraception through Day 90 of the study.
- **[stage 1, prior to futility assessment, only]** Presence at enrollment of any of the following:
  a. stroke
  b. meningitis
  c. encephalitis
  d. myelitis
  e. myocardial infarction
  f. myocarditis
  g. pericarditis
  h. symptomatic congestive heart failure (NYHA class III-IV)
  i. arterial or deep venous thrombosis or pulmonary embolism

- **[stage 1, prior to futility assessment, only]** Current or imminent requirement for any of the following:
    a. invasive mechanical ventilation
    b. ECMO
    c. mechanical circulatory support
    d. vasopressor therapy
    e. commencement of renal replacement therapy at this admission (i.e. not patients on chronic renal replacement therapy).

**Outcomes**

Day 5 Ordinal Outcomes use for Early Futility

Two ordinal outcomes used to determine early futility were assessed at day 5. The first ordinal outcome is a 7-category outcome largely based on oxygen requirements. The highest category that applies on day 5 was assigned. This outcome is referred to as the “pulmonary” ordinal outcome and is defined below:

1. Can independently undertake usual activities with minimal or no symptoms
2. Symptomatic and currently unable to independently undertake usual activities but no need of supplemental oxygen (or not above premorbid requirements)
3. Supplemental oxygen (<4 liters/min, or <4 liters/min above premorbid requirements)
4. Supplemental oxygen (≥4 liters/min, or ≥4 liters/min above premorbid requirements, but not high-flow oxygen)
5. Non-invasive ventilation or high-flow oxygen
6. Invasive ventilation, extracorporeal membrane oxygenation (ECMO), mechanical circulatory support, or new receipt of renal replacement therapy
7. Death

The second ordinal outcome, also assessed at Day 5, captures the range of organ dysfunction that may be associated with progression of Coronavirus-Induced Disease 2019 (COVID-19), such as respiratory dysfunction and coagulation-related complications. Again, the highest category that applies on day 5 was assigned. This outcome is referred to as the “pulmonary+” ordinal outcome. The 7 categories of the pulmonary+ ordinal outcome assessed at Day 5 are:

1. Can independently undertake usual activities with minimal or no symptoms
2. Symptomatic and currently unable to independently undertake usual activities but no need of supplemental oxygen (or not above premorbid requirements)
3. Supplemental oxygen (<4 liters/min, or <4 liters/min above premorbid requirements)
4. Supplemental oxygen (≥4 liters/min, or ≥4 liters/min above premorbid requirements, but not high-flow oxygen) or any of the following: stroke (NIH Stroke Scale [NIHSS] ≤14), meningitis, encephalitis, myelitis, myocardial infarction, myocarditis, pericarditis, new onset CHF NYHA class III or IV or worsening to class III or IV, arterial or deep venous thromboembolic events.
5. Non-invasive ventilation or high-flow oxygen, or signs and symptoms of an acute stroke (NIHSS >14)
6. Invasive ventilation, ECMO, mechanical circulatory support, vasopressor therapy, or new receipt of renal replacement therapy
7. Death

Primary Efficacy Endpoint

The primary endpoint is ***time from randomization to sustained recovery,*** where sustained recovery is defined as being discharged from the index hospitalization, followed by being alive and *home* for 14 consecutive days prior to Day 90.

*Home* is defined as the level of residence or facility where the participant was residing prior to hospital admission leading to enrollment in this trial (the index hospitalization).

Residence or facility groupings to define home are:
1) **Independent/community dwelling** with or without help, including house, apartment, undomiciled/homeless, shelter, or hotel;
2) **Residential care facility** (e.g., assisted living facility, group home, other non-medical institutional setting);
3) **Other healthcare facility** (e.g., skilled nursing facility, acute rehab facility); and
4) L**ong-term acute care hospital** (hospital aimed at providing intensive, longer term acute care services, often for more than 28 days).

Lower (less intensive) level of residence or facility will also be considered as home. By definition, "home" cannot be a "short-term acute care" facility. Participants previously affiliated with a "long-term acute care" hospital recover when they return to the same or lower level of care.

Readmission from "home" may occur and if this occurs within 14 days of the first discharge to "home", then the primary endpoint will not be reached until such time as the participant has been at home for 14 consecutive days. Participants residing in a facility solely for public health or quarantine purposes will be considered as residing in the lowest level of required residence had these public health measures not been instated.

Safety Outcome
The primary safety endpoint, which was assessed at day 5, was defined as a composite of deaths, serious adverse events (SAEs), or grade 3 or 4 AEs. This composite safety was also expanded to include serious clinical events that included end organ dysfunction and serious infections. These serious events are outlined in section 10.2.5 of the protocol and were defined as "protocol-specified exempt events". The events were systematically reported during follow-up but not reported as a SAE unless they were considered related to study agent.

This expanded composite is also reported through day 28.

Adverse events were graded for severity using a toxicity table of the Division of AIDS, NIAID.[1] For adverse events not in the table, a generic grading scheme was used. Adverse events were categorized according to codes in the Medical Dictionary for Regulatory Activities (MedDRA®), version 23.1.

Other safety outcomes included in the protocol and the data analysis plan were:

- Deaths through day 90; and
- A composite of SAEs, including the protocol-specified exempt events, or death through day 90.

In our earlier report, we speculated that one possibility why bamlanivimab failed the futility assessment is that harmful effects may have occurred such as antibody exaggerated inflammation. To address this, stored samples were used to measure plasma levels of interleukin-6 (IL-6), serum levels of C-reactive protein (CRP), and plasma levels of D-dimer.

**Laboratory Methods**
Laboratory specimens were collected for consenting participants and stored by clinical sites and periodically sent to a central biorepository, Advanced BioMedical Laboratories (ABML), for use in future research.

A nasal mid-turbinate swab was collected at baseline. Swabs were immediately placed into tubes containing 3 mL of sterile Universal Transport Medium (UTM). Samples were aliquoted into 3 cryovials, frozen, and shipped on a regular basis to ABML

Four 1.0 mL aliquots or serum and four 1 mL aliquots of plasma were collected at baseline, and on follow-up days 1, 3, 5, 28 and 90. Two 9mL tubes, one SST and one EDTA of blood was drawn to obtain the 8 aliquots.

SARS-CoV-2 RNA load
SARS-CoV-2 RNA load in the nasal swab material was determined using extraction, master mix preparation, and RT-PCR as described in the CDC 2019-Novel Coronavirus (2019-nCoV) Real-Time RT-PCR Diagnostic Panel. The lower limit of quantification (LLoQ) for this measurement is 399 copies/mL.

Viral RNA measurements were centrally determined by ABML.

SARS-CoV-2 Lineages and Variants

The nasal swab material was also used to determine viral lineages and variants. cDNA and amplicons were prepared as described in the ARTIC protocol[2]. Qualitative Assessment of the amplicon was also performed by Bioanalyzer DNA 1000 Chip (Agilent, Santa Clara, CA). Library preparation and sequencing was carried out at the National Human Genome Research Institute. The Cecret pipeline (https://github.com/UPHL-BioNGS/Cecret) was adopted as our sequencing analysis workflow backbone with some modification and addition of new components. Sequence reads in FASTQ format were quality assessed using FastQC (v0.11.9)[3]with a minimum frequency threshold of 0.6 and a minimum depth of 10 reads. They were then adapter and quality trimmed with SeqyClean (v1.10.09)[4]. Trimmed reads were aligned to the SARS-Cov2 reference (Genbank accession MN908947.3) using BWA (v0.7.17-r1188)[5] and primer sequences were masked and consensus sequences called using iVar (v1.3.1)[6]. SARS-CoV-2 Nextstrain clade assignments and variants were determined by Nextclade (v0.14.0)[7] based on the consensus sequences. PANGO lineages were assigned by pangolin (v2.3.3)[8]. Multiple sequence alignment of the consensus sequences was performed using MAFFT (v7.475) and a phylogenetic tree produced from the alignment with IQ-TREE (v1.6.7) and plotted with iTOL (v6; https://itol.embl.de/).

Antibody Levels

Stored plasma specimens were used to measure total anti-SARS-CoV-2 antibody levels. Antibody levels were determined using the BioRad Platelia SARS-CoV-2 Total Ab assay (BioRad, Hercules, California) (anti-N antibodies). Results of this assay are reported as "specimen ratios". Specimen ratios are defined as the specimen optical density (OD) divided by the OD of the control R4($OD_M$R4). Specimen ratios ≥ 1.0 are considered positive, those between 0.8 and 1.0 equivocal, and those < 0.8 negative. In this report, we refer to those with levels < 1.0 specimen ratios as having "negative" anti-N Abs and those with specimen ratios ≥ 1.0 as having positive anti-N Abs.

Levels of neutralizing antibodies (nAbs) directed against the SARS-CoV-2 receptor binding domain (RBD) were determined using the GenScript SARS-CoV-2 Surrogate Virus Neutralization Test (sVNT) assay (GenScript, Piscataway, NJ) (nAbs). nAbs are expressed as percent binding inhibition; levels ≥ 30% are considered positive for nAbs as recommended by the manufacturer, and those < 30% are considered negative for nAbs.

Both antibody determinations were made centrally at the Frederick National Laboratory, blinded to treatment group.

Antigen Levels

SARS-CoV-2 nucleocapsid antigen levels were determined in 90 µL plasma in duplicate using a Quanterix assay (Quanterix, Billerica, MA). The lower level of quantification was determined to be 3 ng/L. Results below that level are imputed as 2.9 ng/L. The antigen determinations were made centrally at the Frederick National Laboratory, blinded to treatment group.

Interleukin-6, C-Reactive Protein and D-dimer

Plasma levels of interleukin-6 (IL-6) and serum levels of C-reactive protein (CRP) were measured using electrochemilumiescence (Meso Scale Discovery, Gaithersburg, MD) at baseline and at days 1, 3, and 5. D-dimer was measured by an enzyme-linked fluorescent assay on a VIDAS instrument (BioMerieux, Durham, NC). Upper limits of normal for IL-6, CRP, and D-dimer are 2 ng/L, 10 mg/L, and 0.5 mg/L, respectively.

**Pre-Specified Hypotheses Based on Antibody and Viral Levels**

Prior to unblinding the 90 day follow-up results for Bamlanivimab, an analysis plan was developed using these determinations that was based on biological reasoning. This supplemental analysis plan is included as a separate appendix.

As part of that plan two subgroup hypotheses were stated for the primary endpoint:

1) Patients with negative or low positive neutralizing antibody levels at entry (GenScript) will benefit more from the investigational agent compared to placebo than patients with high antibody levels. Furthermore, those with low neutralizing antibody levels AND with high antigen levels, will benefit more from the

investigational agent compared to placebo than other subgroups categorized by both antibody and antigen levels.

2) Patients with lower neutralizing antibody levels at entry (Genscript) AND with high levels of RNA in nasal turbinates will benefit more from the investigational agent compared to placebo than other subgroups categorized by both antibody and RNA levels.

While the primary hypotheses consider the neutralizing antibody levels (nAbs; Genscript), similar pre-specified analyses were stated for the total antibody levels (BioRad).

In addition to the primary endpoint, subgroup analyses are also carried out for day 28 and day 90 composite safety outcomes and for mortality through day 90.

**Sample Size to Assess Primary Endpoint**

For the primary end point of sustained recovery we estimated that 843 primary events would be accrued if 1,000 patients were followed for 90 days; 843 primary events provides 90% power at the 0.025 (1-sided) level of significance to detect a sustained recovery rate ratio (RRR) (investigational agent/placebo) of 1.25

**Statistical Methods**

As previously described, following the DSMB recommendation on October 26, 2020 that no further participants be randomized based on a planned futility assessment that was pre-specified in the protocol, an analysis data set that administratively censored follow-up data on October 26, 2020 was locked in order to prepare a preliminary report.[9] The median follow-up at the time of the preliminary report was 31 days.

Data collected after October 26, 2020 remained blinded to investigators until all participants completed the planned 90 days of follow-up. The statistical analysis plan for primary and secondary endpoints and for early futility analyses using the day 5 ordinal outcomes are included in the appendix of our earlier report[9].

Following unblinding of the clinical data, stored specimen analyses were planned as described in Methods of this supplement. After preliminary baseline levels of antibody and antigen data were generated (samples were analyzed in batches), a supplementary analysis plan was developed before carrying out the planned subgroup analyses by antibody level, antigen level, and viral RNA level.

A proportional odds model was used to summarized the 7-category ordinal outcome assessed at Day 5; this intermediate outcome was for assessing futility (see Figure S1).[9]

In addition, to summarizing antibody levels (nAb and anti-N Ab) as percentage positive during follow-up (main paper), box plots of levels reported, including those negative or indeterminate, are given and treatment differences at each time-point are summarized using analysis of covariance with the baseline antibody level as a covariate. Antigen levels are summarized with similar methods.

Treatment comparisons of the percentage of participants with antigen levels < 3 ng/L at each follow-up visit through day 5 are summarized in the main paper with the difference in the percentages; in the supplement, logistic regression with $\log_{10}$ antigen levels as a covariate is used to summarize the treatment differences for antigen levels < 3 and <1000 ng/L, and odds ratios (ORs) and 95% CIs are cited.

Composite safety outcomes through day 28 and day 90 and mortality through day 90 are summarized using time-to-event methods, Kaplan-Meier curves and proportional hazards regression. Hazard ratios (HRs) and 95% confidence intervals (CIs) are cited. For completeness other safety outcomes, infusion reactions and day 5 composite safety outcomes are also cited. The percentage with these outcomes in each treatment group are compared using logistic regression models and ORs are cited.

The protocol defined a number of baseline-defined subgroup analyses for the primary endpoint of sustained recovery and for the Day 5 and Day 28 primary safety outcome. In this report, we summarize day 5, day 28 and day 90 safety outcomes by antibody levels and sustained recoverey for selected other baseline-defined subgroups. Heterogeneity of the treatment effect across subgroups was assessed by including interaction terms between treatment group and baseline subgroups in logistic or proportional hazards regression models for these outcomes.

#### Section 3: Results

This section briefly summarizes tables and figures included in this supplement. The tables and figures are summarized in the order they are cited in the main paper,

**Figure S1.** A few updates to the day 5 ordinal pulmonary outcome were made since the preliminary report was published.[6] These final data indicate that the odds of a more favorable outcome for bamlanivimab compared to placebo is 0.86 favoring placebo (95% CI: 0.57-1.30; p=0.48).

**Figure S2.** 314 participants (169 bamlanivimab, 157 placebo) were infused and followed through 90 days. The database for this trial was locked after all participants completed the day 90 visit.

**Table S1.** Viral RNA results at study entry are given by plasma antigen levels. Overall, the Spearman correlation coefficient for the two measures was .26 (p<.001). Among the 59 participants with viral RNA levels classified as negative or indeterminate, 90% had positive antigen levels.

We also considered correlations of each marker of viral replication with IL-6 at baseline. The rank correlation between antigen and IL-6 levels was 0.37 (p<0.001) and between viral RNA level and IL-6 was 0.11 (p=0.08).

**Figure S3.** Sequences from 255 participants identified no concerning mutations in codons 417,452 or 484 in the spike protein; virus from 6 participants had deletions in codon 69-70.

Coverage of 75% or more of genome achieved in 204 of 255 samples; the insert displays phylogeny (with indication of Pango Lineage and Nextstrain clade) with genome coverage color coded (red: < 75% and ≥ 50 (n=21, 30 samples with coverage <50% not included in the plot); green: >75% general and 100% of spike (n=152); blue and yellow: >75% general and less than 100% (yellow; n=21) or 100% (blue; n=31) for RBD within spike). Samples color-coded green and red are displayed to illustrate coverage below (bottom sample is reference).

Mutations compared with reference genomic sequence (NCBI Accession: MN908947.3) were called with a minimum frequency threshold of 0.6 and a minimum depth of 10 reads with iVar. The most frequently found Spike mutations included: D614G (222), K1045R (11), A222V (8), H69-V70 deletion (6), P681H (5), N439K (5), L5F (4), Q677H (4), A688V (4), G946V (4).

In RBD, the following additional mutations were found: N439K (5), S477N (4), R346K (2), P384L (1), T385I (1), T470N (1), N501Y (1), A522V (1).

**Table S2.** Characteristics at entry are given by nAb sero-status at entry. Among those nAb positive, median (IQR) plasma antigen levels are 297 (37, 1924). Among those nAb negative, the corresponding levels are 2130 (796, 5060). Among those positive for nAbs, 86% are positive for anti-N antibodies.

**Table S3.** Among those anti-N antibody positive, median (IQR) plasma antigen levels are 634 (64, 2790). Among those nAb negative, antigen levels are 1930 (776, 4640). Among those positive for anti-N antibodies, 73% are positive for nAbs.

**Table S4.** Characteristics at entry are given by baseline antibody status that considers both nAb and anti-N antibodies. Among those sero-positive to either nAbs or anti-N Abs, median (IQR) plasma antigen levels are 683 (69, 2802). Among those negative to both nAbs and anti-N Abs, the corresponding levels are 1941 (784, 4960) ng/L.

**Figure S4.** nAb levels measured with the GenScript assay were determined at baseline and days 1, 3, and 5. These results are given overall (A) and according to nAb status at entry: (B) negative and (C) positive. This figure complements Figure 1A, 1B and 1C in the main paper which gives the percentage positive (≥ 30% for binding inhibition). In Figure S4, higher levels for those in the bamlanivimab compared to the placebo group are evident at each follow-up visits, overall and for those nAb negative and positive at baseline.

**Figure S5.** Anti-N antibody levels with the BioRad assay were determined at baseline and days 1, 3, and 5. These results are given overall (A) and according to anti-N antibody status at entry: (B) negative and (C) positive. This figure complements Figure 1D, 1E and 1F in the main paper which gives the percentage positive (≥ 1.0 for the specimen ratio). Anti_N levels during follow-up, like the percentage positive shown in the main paper, did not differ significantly between treatment groups, overall or according to anti-N Ab sero-postive status at baseline.

**Table S5.** According to the supplementary analysis plan developed, antigen levels < 3 and < 1000 ng/L for the bamlanivimab and placebo groups were to be compared at day 5 using logistic regression with baseline log10 transformed antigen level as a covariate. These results are summarized in Table S5 and complement the analyses shown in Figure 2 of the main report. At day 5 the OR (bamlanivimab versus placebo) for antigen levels < 3 ng/mL is 1.40 (95% CI: 0.74-2.66; p=0.30). The OR for antigen levels < 1000 ng/L at day 5 is 1.24 (95% CI: 0.23-6.64; p=0.80). At no timepoint did these ORs differ significantly from 1.0.

**Figure S6.** Antigen levels at baseline and days 1, 3, and 5 after $\log_{10}$ transformation are given overall (A) and according to nAb antibody status at entry: (B) negative and (C) positive in Figure S6. Figure S6 complements Figure 2A, 2B, and 2C in the main paper. Antigen levels < 3 ng/L, the lower level of quantification, are imputed as 2.9 ng/L in these analyses. Antigen levels during follow-up, like the percentage < 3 ng/L, did not differ significantly between treatment groups, overall or according to nAb sero-positive status at baseline.

**Figure S7.** Baseline-defined subgroups that consider demographic and clinical factors which were pre-specified for the the primary endpoint of sustained recovered are summarized in Figure S7. There was no evidence of heterogeneity among the RRRs for any of the subgroups considered in Figure S7.

**Figure S8.** In the supplementary analysis plan, methods for evaluating subgroups using baseline anti-N Abs levels instead of nAb levels were also stated. Figure S8 summarizes these subgroups for the sustained recovery outcome in a similar manner to those presented in Figure 4. Like the finding for negative and positive nAbs in Figure 4, RRRs for sustained recovery were significantly greater, favoring bamlanivimab, for those with negative anti-N Abs (< 1.0 specimen ratios) (RRR=1.26) compared to those with positive anti-N Abs (≥ 1.0 specimen ratios) (RRR=0.80) (p=0.05 for the difference in RRRs.

In addition, as hypothesized, among those with negative anti-N abs and with antigen levels ≥ 1000 ng/L or those with viral RNA levels ≥ 10,000 copies/mL and antigen levels ≥ 1000 ng/L had higher RRRs than the other 3 groups defined by anti-N Abs and viral levels. The 3 df interactions for these groups gave p-values for the heterogeneity of RRRs of 0.06 and 0.04, for antigen and viral RNA, respectively.

An analysis was also carried out using classifying participants as having high viral levels by either antigen levels ≥ 1000 ng/L or viral RNA ≥ 10,000, and findings were similar and the interaction p-value was 0.06.

**Figure S9.** Given the similar subgroup findings for nAbs and anti-N Abs, a subgroup analysis for sustained recovery was also carried out classifying participants as negative on both nAbs and anti-N Abs versus positive on either. The percentage of participants negative to both antibodies was 34%. This subgroup yielded a 1df interaction p-value of 0.008 (RRR for those negative to both = 1.47 and RRR for those positive to either = 0.78).

RRRs for those with both antibodies negative and with high viral levels were 1.93, 2.01, and 1.60 for high viral levels defined by antigen, viral RNA and either, respectively. The 3 df interaction p-values for the 4 groups considered in each of these analyses were 0.01, 0.004, and 0.03.

For comparison with other studies, we also estimated the RRR of sustained recovery in the placebo group according to antigen and viral RNA level at entry. These RRRs were estimated with no other covariates in the regression model. The RRR for those with antigen ≥ 1000 vs < 1000 ng/L was 0.44 (95% CI: 0.32 – 0.62); for those with viral RNA ≥ 10000 vs <10000 copies/mL, the RRR was 0.63 (95% CI: 0.45 – 0.87); and for those with either antigen ≥ 1000 ng/L or viral RNA ≥ 10000 copies/mL versus both lower, the RRR was 0.50 (95% CI: 0.35 to 0.72).

**Table S6.** Overall and subgroup analyses by nAb status at entry for the day 90 composite safety outcome and mortality are given in the main paper (Figure 5 A-F). Table S6 summarizes the overall results for each of the pre-specified safety outcomes. We previously reported the percentage of participants with infusion reactions and with the primary composite safety outcome considered for the early futility analysis of death, SAEs, or grade 3 or 4 adverse events by day 5. Since that report 2 additional events were reported in the bamlanivimab group and the OR (bamlanivimab/placebo) is 1.66 (95% CI: 0.87-3.16). The previously reported OR was 1.56.

When this composite outcome at day 5 is expanded to include organ failure events and serious infections, serious events that according to protocol were exempt from SAE reporting unless they were considered related to treatment, the OR was 1.83 (95% CI: 1.01-3.29).

Point estimates for all of the pre-specified outcomes exceeded 1.0.

**Table S7.** Safety outcomes given in Table S6 are summarized by nAb status at baseline in Table S7. With the exception of the composite outcome at day 90 and mortality each of the ORs or HRs for bamlanivimab versus placebo was greater 1.0 for both subgroups and there was no evidence of an interaction. The day 90 composite outcome and mortality are discussed in the main paper.

**Table S8.** The components of the day 90 safety outcome are summarized in Table S8, overall and by nAb status at entry. For each component among those with negative nAbs, HRs were less than 1.0 favoring bamlanivimab; for those with positive nAbs at entry, each HR was > 1.0. Organ failure was the most frequently occuring component of the composite outcomes and most events were categorized as respiratory failure events. Respiratory failure was defined as the receipt of high-flow nasal oxygen, non-invasive ventilation, invasive mechanical ventilation, or ECMO.

**Figure S10 (A, B, C, D, E, and F).** Box plots for $\log_2$ transformed IL-6 (A-C) and CRP (D-F) are shown in Figure S10. Both inflammatory markers declined over follow-up. Overall, median levels of IL-6 on the untransformed scale (data not shown) declined from 7.6 at baseline to 5.0 ng/L at Day 5 for bamlanivimab and from 5.7 to 3.6 ng/L for placebo. Median CRP declined from 153 to 38 mg/L for bamlanivimab and from 130 to 24 mg/L for placebo. As indicated in Figure S10 levels of these biomarkers did not differ significantly from one another at any follow-up visit either overall or by nAb subgroup.

**Figure S11 (A, B, and C).** Box plots for $\log_2$ transformed D-dimer are given in Figure S11 (A-C). D-dimer did not decline from baseline to day 5. At no time-point did levels differ between treatments, overall (A) or in the subgroups defined by nAb status at entry (B and C).

#### Section 4: Tables and Figures

#### Figure S1: Pulmonary Outcome on Day 5

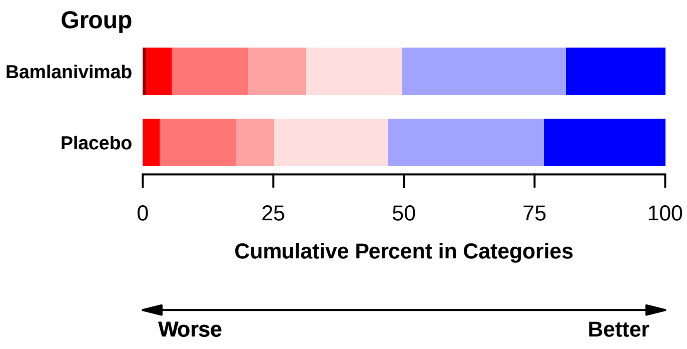

| Group | N | 7 | 6 | 5 | 4 | 3 | 2 | 1 |
|---|---|---|---|---|---|---|---|---|
| Bamlanivimab | 163 | 0.6 | 4.9 | 14.7 | 11.0 | 18.4 | 31.3 | 19.0 |
| Placebo | 151 | 0.0 | 3.3 | 14.6 | 7.3 | 21.9 | 29.8 | 23.2 |

Percent in Category (columns 7–1)

**Odds Ratio, Adjusted***
0.86 (95% CI: 0.57 to 1.30), P=0.48

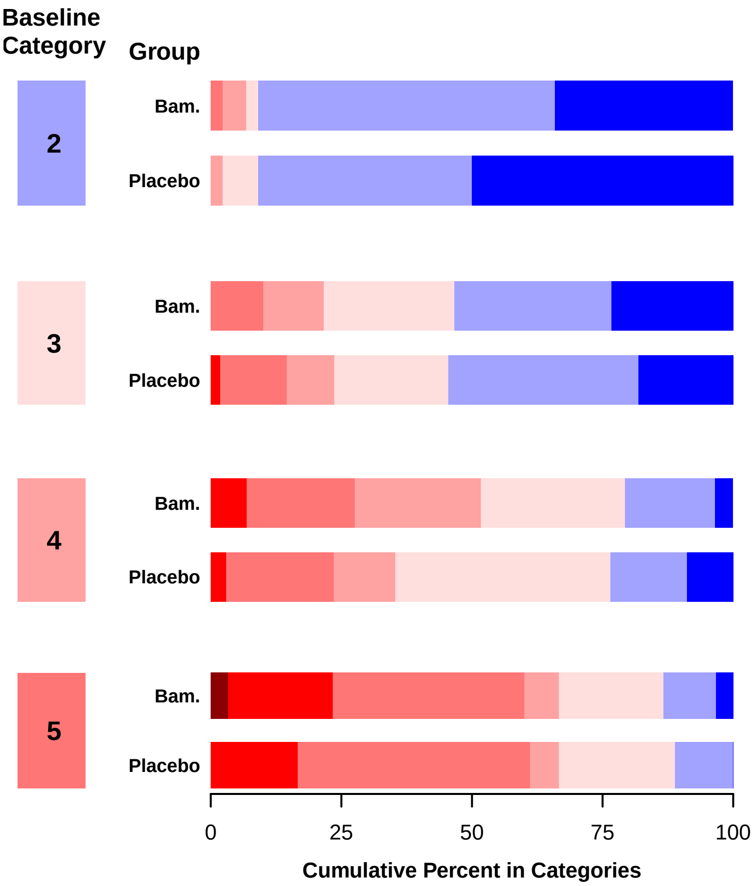

| Baseline Category | Group | N | 7 | 6 | 5 | 4 | 3 | 2 | 1 |
|---|---|---|---|---|---|---|---|---|---|
| 2 | Bam. | 44 | 0.0 | 0.0 | 2.3 | 4.5 | 2.3 | 56.8 | 34.1 |
| 2 | Placebo | 44 | 0.0 | 0.0 | 0.0 | 2.3 | 6.8 | 40.9 | 50.0 |
| 3 | Bam. | 60 | 0.0 | 0.0 | 10.0 | 11.7 | 25.0 | 30.0 | 23.3 |
| 3 | Placebo | 55 | 0.0 | 1.8 | 12.7 | 9.1 | 21.8 | 36.4 | 18.2 |
| 4 | Bam. | 29 | 0.0 | 6.9 | 20.7 | 24.1 | 27.6 | 17.2 | 3.4 |
| 4 | Placebo | 34 | 0.0 | 2.9 | 20.6 | 11.8 | 41.2 | 14.7 | 8.8 |
| 5 | Bam. | 30 | 3.3 | 20.0 | 36.7 | 6.7 | 20.0 | 10.0 | 3.3 |
| 5 | Placebo | 18 | 0.0 | 16.7 | 44.4 | 5.6 | 22.2 | 11.1 | 0.0 |

Percent in Category (columns 7–1)

**Category**
- 1 = Can independently undertake usual activities with minimal/no symptoms
- 2 = No supplemental oxygen; symptomatic and unable to independently undertake usual activities
- 3 = Supplemental oxygen < 4 L/min
- 4 = Supplemental oxygen ≥ 4 L/min
- 5 = Non−invasive ventilation or high−flow oxygen
- 6 = Invasive ventilation, ECMO, mechanical circulatory support, or renal replacement therapy
- 7 = Death

* OR of being in a better category with Bamlanivimab vs. placebo, estimated in a proportional odds model adjusted for baseline category and for study site pharmacy.

**Figure S2: Consort Diagram**

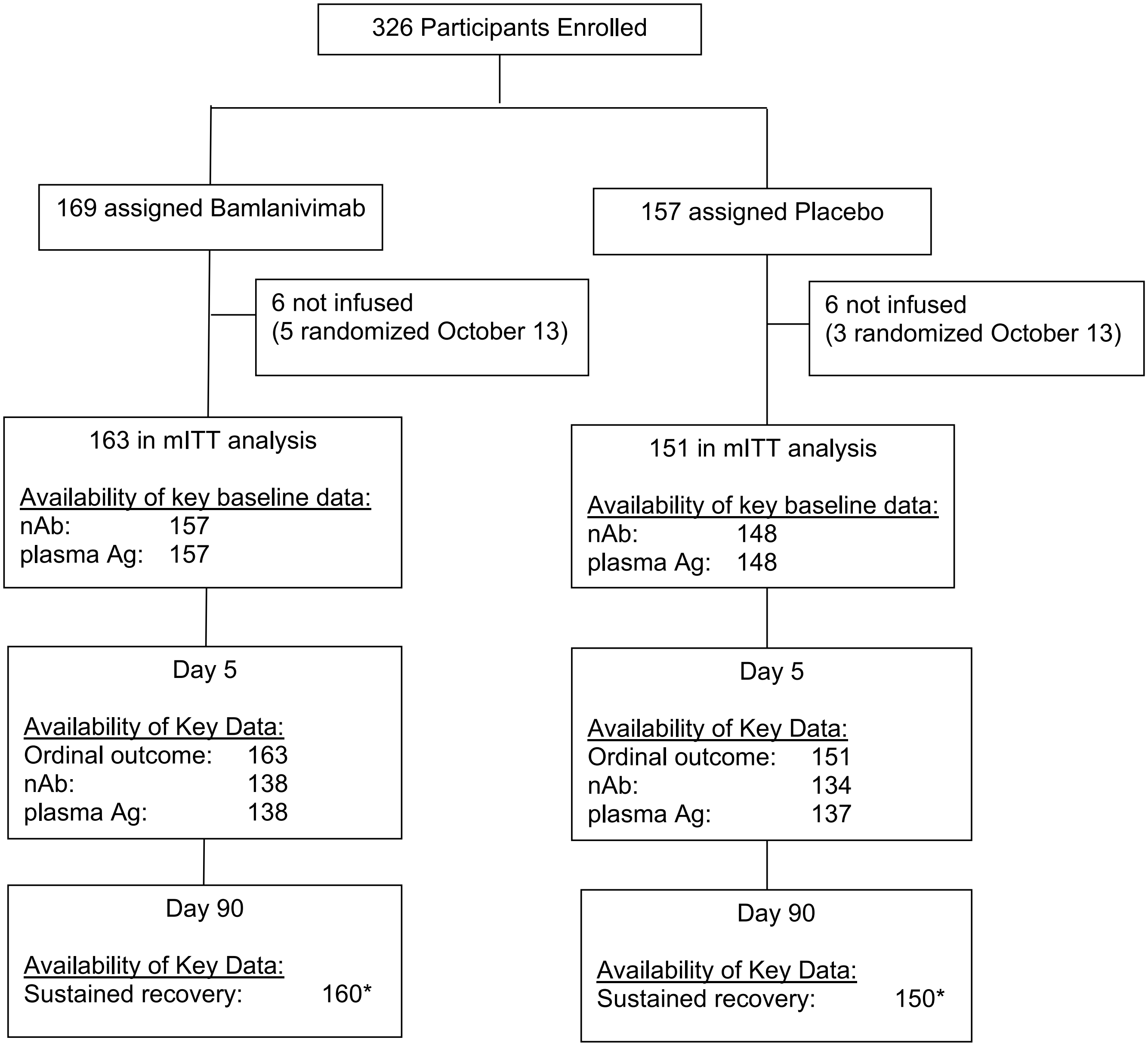

* Three participants in the Bamlanivimab group had followup for events censored at day 7, 16 and 76. One participant in the Placebo group has followup for events censored at day 79.

**Table S1: Baseline Viral RNA by Antigen Level**

| | Antigen Level(pg/mL) | | | | |
|---|---|---|---|---|---|
| **RNA (copies/mL)** | **< 3** | **3-224** | **225-999** | **1000-3724** | **3725+** |
| Neg/indeterminate | 6 (46%) | 25 (35%) | 11 (17%) | 10 (13%) | 7 (10%) |
| < 400 | 2 (15%) | 7 (10%) | 7 (11%) | 10 (13%) | 7 (10%) |
| 400-9,999 | 3 (23%) | 13 (18%) | 14 (22%) | 12 (15%) | 12 (17%) |
| 10,000-199,999 | 0 (0%) | 14 (19%) | 16 (25%) | 29 (36%) | 19 (28%) |
| ≥200,000 | 2 (15%) | 13 (18%) | 15 (24%) | 19 (24%) | 24 (35%) |
| Total | 13 (100%) | 72 (100%) | 63 (100%) | 80 (100%) | 69 (100%) |
| Correlation coefficient* | 0.26 | | | | |
| p-value | < .001 | | | | |

* Spearman; a value of 1 is imputed for negative or indeterminate viral loads

#### Figure S3: Viral Sequencing Summary, Panel A

**A.**

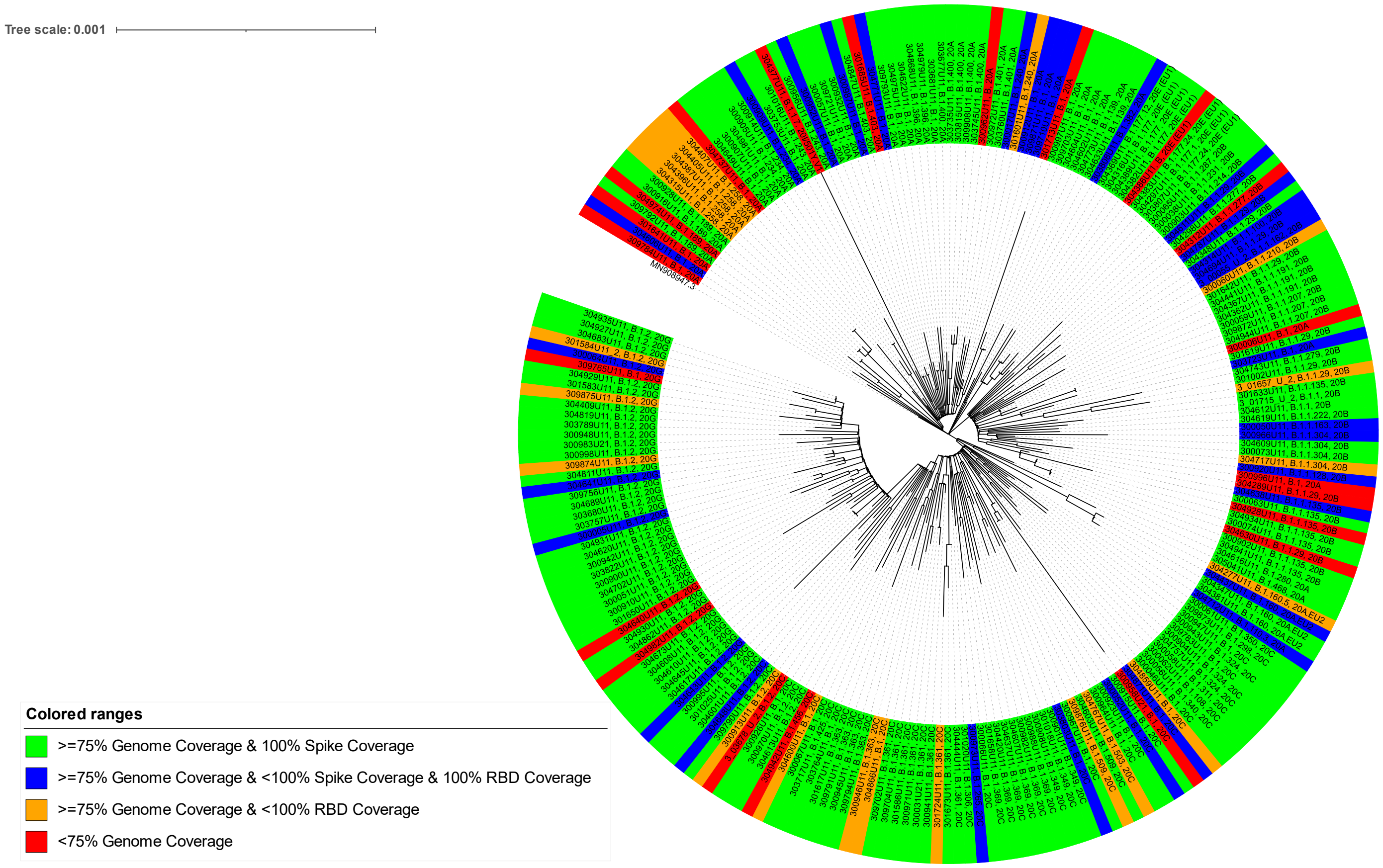

**Figure S3: Viral Sequencing Summary, Panels B-E**

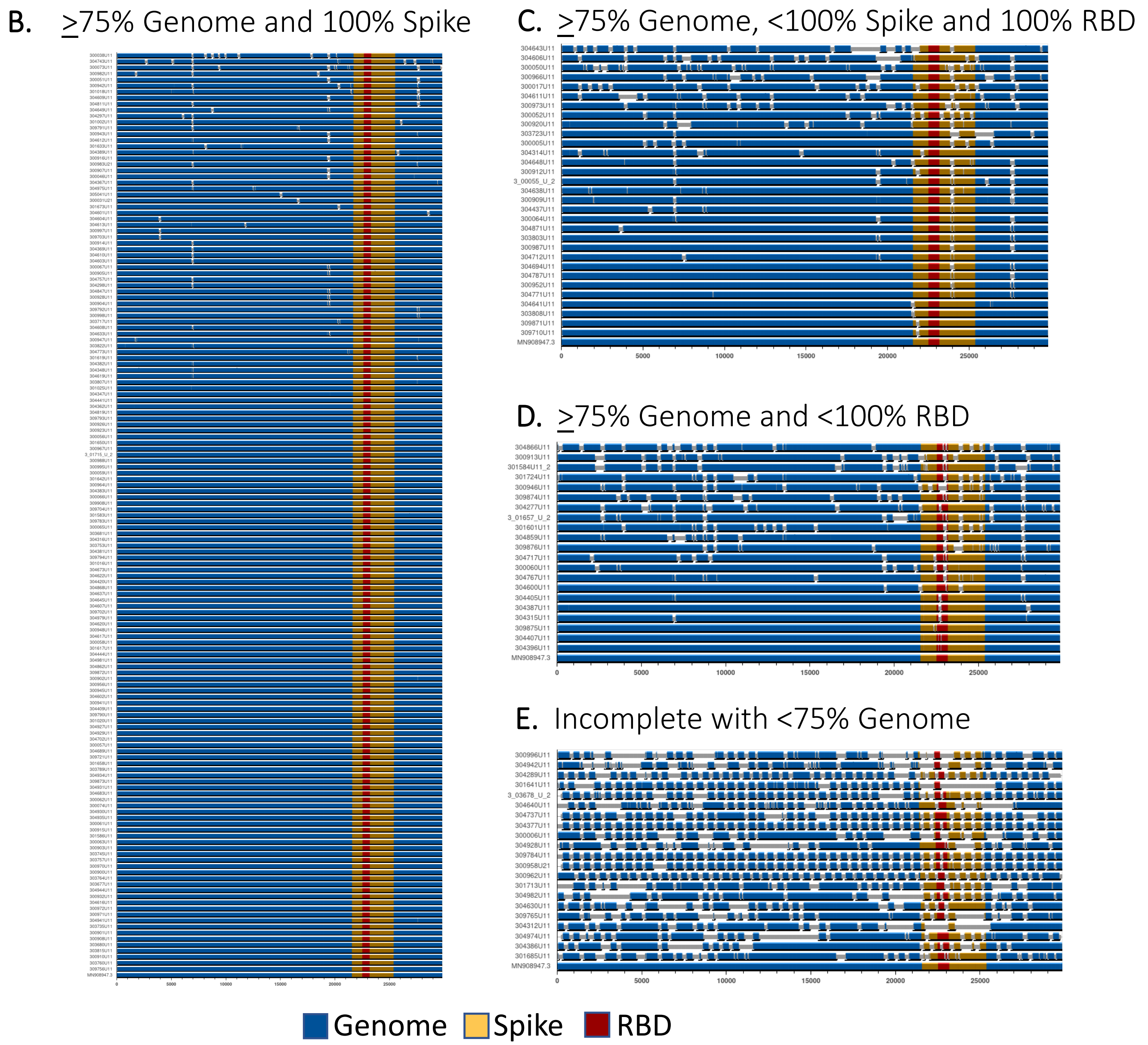

**Table S2: Baseline Characteristics by nAb Sero-status**

| | | Antibody Positive (n=152 ) | Antibody Negative (n=153 ) |
|---|---|---|---|
| Age | Median (IQR) - yr | 58 (47, 68) | 65 (50, 73) |
| Female gender | No. (%) | 64 (42%) | 69 (45%) |
| Non-white race | No. (%) | 85 (56%) | 77 (50%) |
| History of: | No. (%) | | |
| Hypertension requiring medication | | 70 (46%) | 83 (54%) |
| Diabetes requiring medication | | 44 (29%) | 43 (28%) |
| Renal impairment | | 9 (6%) | 22 (14%) |
| Asthma and/or COPD | | 21 (14%) | 23 (15%) |
| At least one co-morbidity | | 101 (66%) | 109 (71%) |
| Days since symptom onset | Median (IQR) | 8 (5, 10) | 7 (4, 8) |
| Oxygen Requirement | No. (%) | | |
| Not receiving supplemental oxygen | | 30 (20%) | 54 (35%) |
| Supplemental oxygen < 4 L/min | | 61 (40%) | 52 (34%) |
| Supplemental oxygen ≥ 4 L/min | | 36 (24%) | 28 (18%) |
| Non-invasive ventilation or HFNC | | 25 (16%) | 19 (12%) |
| Invasive ventilation or ECMO | | 0 (0%) | 0 (0%) |
| Laboratory assessments | | | |
| Plasma nucleocapside antigen | Median ng/L | 297 (37, 1924) | 2130 (796, 5060) |
| ≥ 3 ng/L (positive) | No. (%) | 138 (91%) | 152 (99%) |
| RNA positive (nasal swab) | No. (%) | 103 (70%) | 135 (90%) |
| copies/mL | Median (IQR) | 14595 (649, 131794) | 76310 (2880, 850202) |
| Anti-spike neutralizing antibodies | Median (IQR) - BI* | 61 (46, 78) | 11 (4, 19) |
| Anti-nucleocapsid antibody positive | No. (%) | 131 (86%) | 49 (32%) |
| Interleukin-6 | Median (IQR) - mg/L | 5.7 (2.4, 15.2) | 7.7 (3.1, 14.8) |
| D-dimer | Median (IQR) - ng/L | 0.98 (0.71, 1.39) | 0.84 (0.57, 1.37) |
| C-reative protein | Median (IQR) - ng/L | 196 (43, 655) | 109 (36, 461) |
| B-Lymphocytes | Median (IQR) - $10^9$ | 0.83 (0.56, 1.21) | 0.77 (0.54, 1.05) |

* = binding inhibition

**Table S3: Baseline Characteristics by Anti-N Antibody Sero-status**

| | | Antibody Positive (n=180 ) | Antibody Negative (n=125 ) |
|---|---|---|---|
| Age | Median (IQR) - yr | 59 (47, 69) | 65 (51, 74) |
| Female gender | No. (%) | 71 (39%) | 62 (50%) |
| Non-white race | No. (%) | 104 (58%) | 58 (46%) |
| History of: | No. (%) | | |
| Hypertension requiring medication | | 86 (48%) | 67 (54%) |
| Diabetes requiring medication | | 54 (30%) | 33 (26%) |
| Renal impairment | | 13 (7%) | 18 (14%) |
| Asthma and/or COPD | | 24 (13%) | 20 (16%) |
| At least one co-morbidity | | 122 (68%) | 88 (70%) |
| Days since symptom onset | Median (IQR) | 8 (6, 10) | 6 (4, 8) |
| Oxygen Requirement | No. (%) | | |
| Not receiving supplemental oxygen | | 34 (19%) | 50 (40%) |
| Supplemental oxygen < 4 L/min | | 71 (39%) | 42 (34%) |
| Supplemental oxygen ≥ 4 L/min | | 46 (26%) | 18 (14%) |
| Non-invasive ventilation or HFNC | | 29 (16%) | 15 (12%) |
| Invasive ventilation or ECMO | | 0 (0%) | 0 (0%) |
| Laboratory assessments | | | |
| Plasma nucleocapside antigen | Median ng/L | 634 (64, 2790) | 1930 (776, 4640) |
| ≥ 3 ng/L (positive) | No. (%) | 167 (93%) | 123 (98%) |
| RNA positive (nasal swab) | No. (%) | 126 (72%) | 112 (93%) |
| copies/mL | Median (IQR) | 10762 (561, 80891) | 151535 (13891, 1400018) |
| Anti-nucleocapsid antibodies | Median (IQR) - BI* | 4 (3, 5) | 0 (0, 0) |
| Anti-spike neutralizing antibody positive | No. (%) | 131 (73%) | 21 (17%) |
| Interleukin-6 | Median (IQR) - mg/L | 5.9 (2.5, 15.5) | 7.6 (3.1, 14.7) |
| D-dimer | Median (IQR) - ng/L | 0.98 (0.68, 1.39) | 0.83 (0.57, 1.37) |
| C-reative protein | Median (IQR) - ng/L | 240 (43, 663) | 98 (38, 364) |
| B-Lymphocytes | Median (IQR) - $10^9$ | 0.82 (0.56, 1.20) | 0.77 (0.53, 1.02) |

* = binding inhibition

**Table S4: Baseline Characteristics Considering Both nAb and Anti-N Antibody Sero-status**

| | | Positive nAb or Anti-N Ab (n=201 ) | Negative nAb and Anti-N Ab (n=104 ) |
|---|---|---|---|
| Age | Median (IQR) - yr | 59 (48, 69) | 67 (51, 76) |
| Female gender | No. (%) | 81 (40%) | 52 (50%) |
| Non-white race | No. (%) | 113 (56%) | 49 (47%) |
| History of: | No. (%) | | |
| Hypertension requiring medication | | 94 (47%) | 59 (57%) |
| Diabetes requiring medication | | 58 (29%) | 29 (28%) |
| Renal impairment | | 15 (7%) | 16 (15%) |
| Asthma and/or COPD | | 26 (13%) | 18 (17%) |
| At least one co-morbidity | | 134 (67%) | 76 (73%) |
| Days since symptom onset | Median (IQR) | 8 (6, 10) | 6 (4, 8) |
| Oxygen Requirement | No. (%) | | |
| Not receiving supplemental oxygen | | 41 (20%) | 43 (41%) |
| Supplemental oxygen < 4 L/min | | 78 (39%) | 35 (34%) |
| Supplemental oxygen ≥ 4 L/min | | 50 (25%) | 14 (13%) |
| Non-invasive ventilation or HFNC | | 32 (16%) | 12 (12%) |
| Invasive ventilation or ECMO | | 0 (0%) | 0 (0%) |
| Laboratory assessments | | | |
| Plasma nucleocapside antigen | Median ng/L | 683 (69, 2802) | 1941 (784, 4960) |
| ≥ 3 ng/L (positive) | No. (%) | 187 (93%) | 103 (99%) |
| RNA positive (nasal swab) | No. (%) | 142 (73%) | 96 (94%) |
| copies/mL | Median (IQR) | 13509 (583, 131112) | 193316 (13548, 1517650) |
| Interleukin-6 | Median (IQR) - mg/L | 6.7 (2.7, 16.4) | 6.5 (2.9, 12.6) |
| D-dimer | Median (IQR) - ng/L | 0.98 (0.68, 1.34) | 0.82 (0.56, 1.39) |
| C-reative protein | Median (IQR) - ng/L | 206 (42, 642) | 98 (36, 339) |
| B-Lymphocytes | Median (IQR) - $10^9$ | 0.83 (0.56, 1.20) | 0.74 (0.51, 0.98) |

* = binding inhibition

### Figure S4: nAb Levels by Study Day and Treatment Group

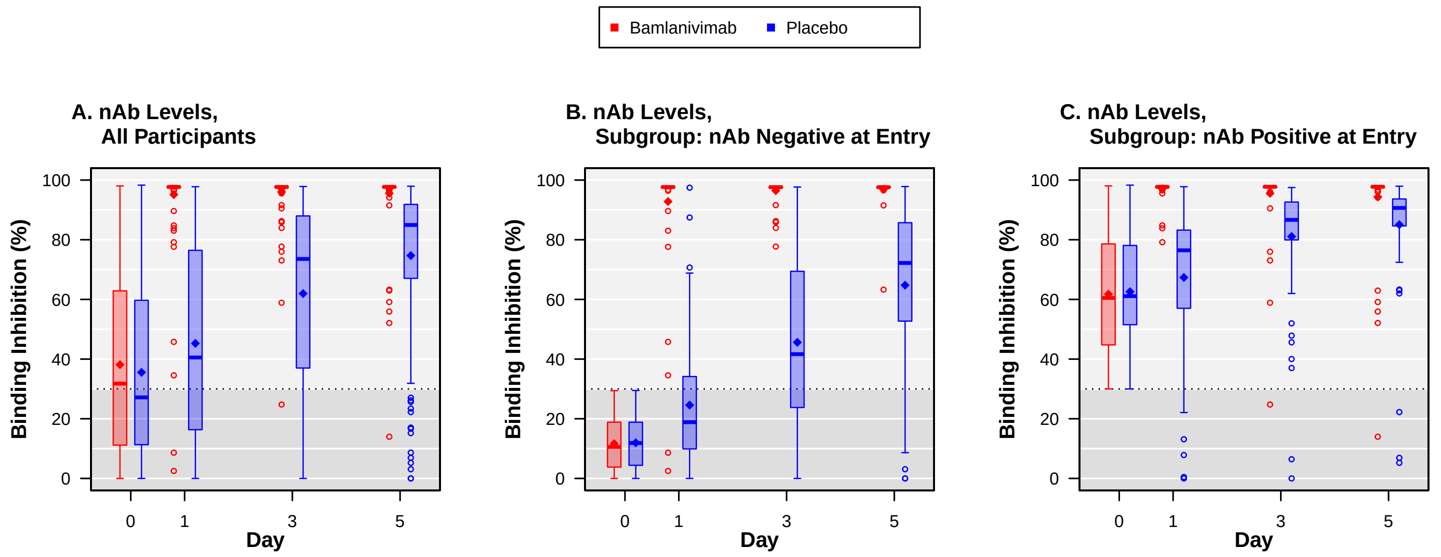

**A. All Participants**

| Number of Patients | | | | |
|---|---|---|---|---|
| Bam.: | 157 | 153 | 146 | 138 |
| Placebo: | 148 | 139 | 142 | 134 |

| Est. Mean Diff. (Bam.−Placebo) | | | |
|---|---|---|---|
| Est. Diff.: | 50 | 33 | 21 |
| 95% CI: | (45, 54) | (28, 37) | (16, 25) |
| P−value: | <0.001 | <0.001 | <0.001 |

**B. Subgroup: nAb Negative at Entry**

| Number of Patients | | | | |
|---|---|---|---|---|
| Bam.: | 74 | 71 | 65 | 61 |
| Placebo: | 79 | 72 | 77 | 69 |

| Est. Mean Diff. (Bam.−Placebo) | | | |
|---|---|---|---|
| Est. Diff.: | 68 | 51 | 32 |
| 95% CI: | (62, 75) | (44, 58) | (25, 39) |
| P−value: | <0.001 | <0.001 | <0.001 |

**C. Subgroup: nAb Positive at Entry**

| Number of Patients | | | | |
|---|---|---|---|---|
| Bam.: | 83 | 78 | 78 | 74 |
| Placebo: | 69 | 64 | 63 | 62 |

| Est. Mean Diff. (Bam.−Placebo) | | | |
|---|---|---|---|
| Est. Diff.: | 30 | 14 | 9 |
| 95% CI: | (25, 36) | (10, 19) | (4, 15) |
| P−value: | <0.001 | <0.001 | <0.001 |

### Figure S5: Anti-N Antibody Levels by Study Day and Treatment Group

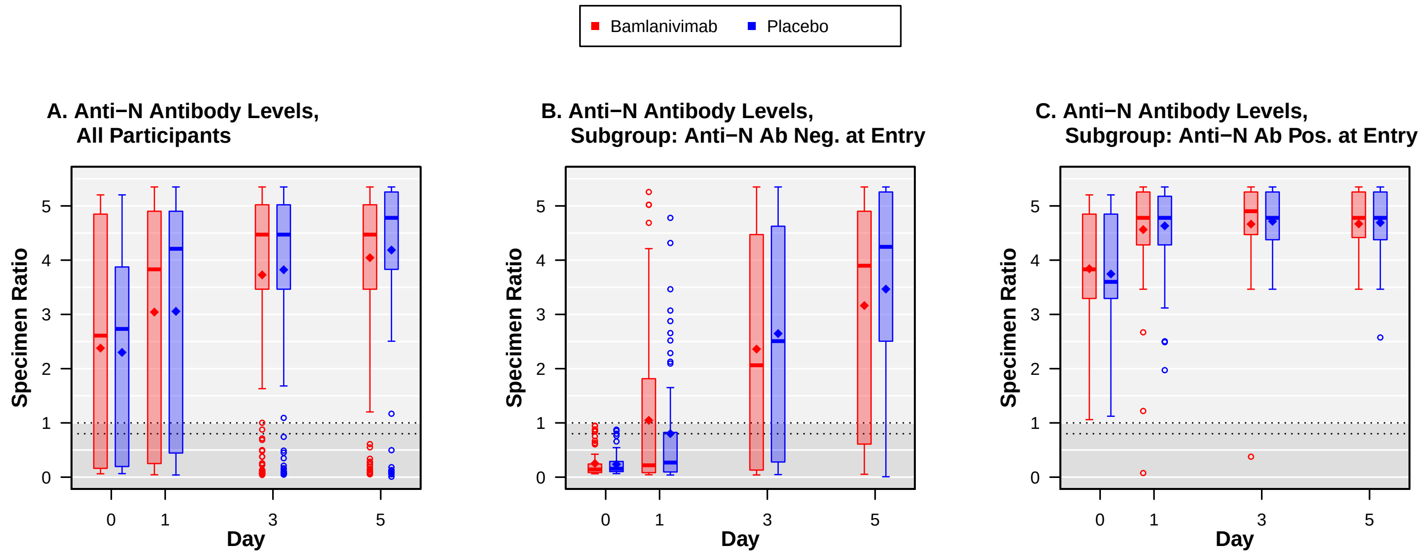

**A. All Participants**

| Number of Patients | | | | |
|---|---|---|---|---|
| Bam.: | 157 | 155 | 147 | 138 |
| Placebo: | 148 | 140 | 141 | 136 |

| Est. Mean Diff. (Bam.−Placebo) | | | |
|---|---|---|---|
| Est. Diff.: | −0.007 | −0.19 | −0.14 |
| 95% CI: | (−0.30, 0.28) | (−0.53, 0.16) | (−0.47, 0.19) |
| P−value: | 0.96 | 0.29 | 0.41 |

**B. Subgroup: Anti−N Ab Neg. at Entry**

| Number of Patients | | | | |
|---|---|---|---|---|
| Bam.: | 64 | 63 | 57 | 53 |
| Placebo: | 61 | 57 | 59 | 54 |

| Est. Mean Diff. (Bam.−Placebo) | | | |
|---|---|---|---|
| Est. Diff.: | 0.18 | −0.33 | −0.38 |
| 95% CI: | (−0.12, 0.47) | (−0.99, 0.33) | (−1.11, 0.36) |
| P−value: | 0.23 | 0.32 | 0.31 |

**C. Subgroup: Anti−N Ab Pos. at Entry**

| Number of Patients | | | | |
|---|---|---|---|---|
| Bam.: | 64 | 63 | 57 | 53 |
| Placebo: | 61 | 57 | 59 | 54 |

| Est. Mean Diff. (Bam.−Placebo) | | | |
|---|---|---|---|
| Est. Diff.: | −0.08 | −0.053 | −0.023 |
| 95% CI: | (−0.32, 0.16) | (−0.26, 0.15) | (−0.21, 0.16) |
| P−value: | 0.51 | 0.61 | 0.81 |

**Table S5: Antigen Levels Below Selected Levels by Study Day and Treatment Group**

**Antigen Levels below the Level of Quantification***

| Visit | Banlanivimab N Pts | Banlanivimab N | Banlanivimab % | Placebo N Pts | Placebo N | Placebo % | Treatment Difference** OR | Treatment Difference** 95% CI | Treatment Difference** P-value |
|---|---|---|---|---|---|---|---|---|---|
| Day 0 | 157 | 9 | 5.7 | 148 | 6 | 4.1 | | | |
| Day 1 | 155 | 12 | 7.7 | 141 | 5 | 3.5 | 3.50 | 0.96, 12.76 | 0.06 |
| Day 3 | 147 | 29 | 19.7 | 142 | 22 | 15.5 | 1.46 | 0.64, 3.30 | 0.37 |
| Day 5 | 138 | 61 | 44.2 | 137 | 59 | 43.1 | 1.40 | 0.74, 2.66 | 0.30 |

**Antigen Levels < 1000 ng/L**

| Visit | Banlanivimab N Pts | Banlanivimab N | Banlanivimab % | Placebo N Pts | Placebo N | Placebo % | Treatment Difference** OR | Treatment Difference** 95% CI | Treatment Difference** P-value |
|---|---|---|---|---|---|---|---|---|---|
| Day 0 | 157 | 71 | 45.2 | 148 | 81 | 54.7 | | | |
| Day 1 | 155 | 86 | 55.5 | 141 | 86 | 61.0 | 1.07 | 0.52, 2.21 | 0.85 |
| Day 3 | 147 | 137 | 93.2 | 142 | 132 | 93.0 | 1.05 | 0.37, 2.94 | 0.93 |
| Day 5 | 138 | 135 | 97.8 | 137 | 134 | 97.8 | 1.24 | 0.23, 6.64 | 0.80 |

* LOQ = 3.0 by Quanterix antigen test

** Odds ratio from logistic model with adjustment for baseline $\log_{10}$ antigen levels.

**Figure S6: Antigen Levels by Study Day and Treatment Group and by nAb at Entry**

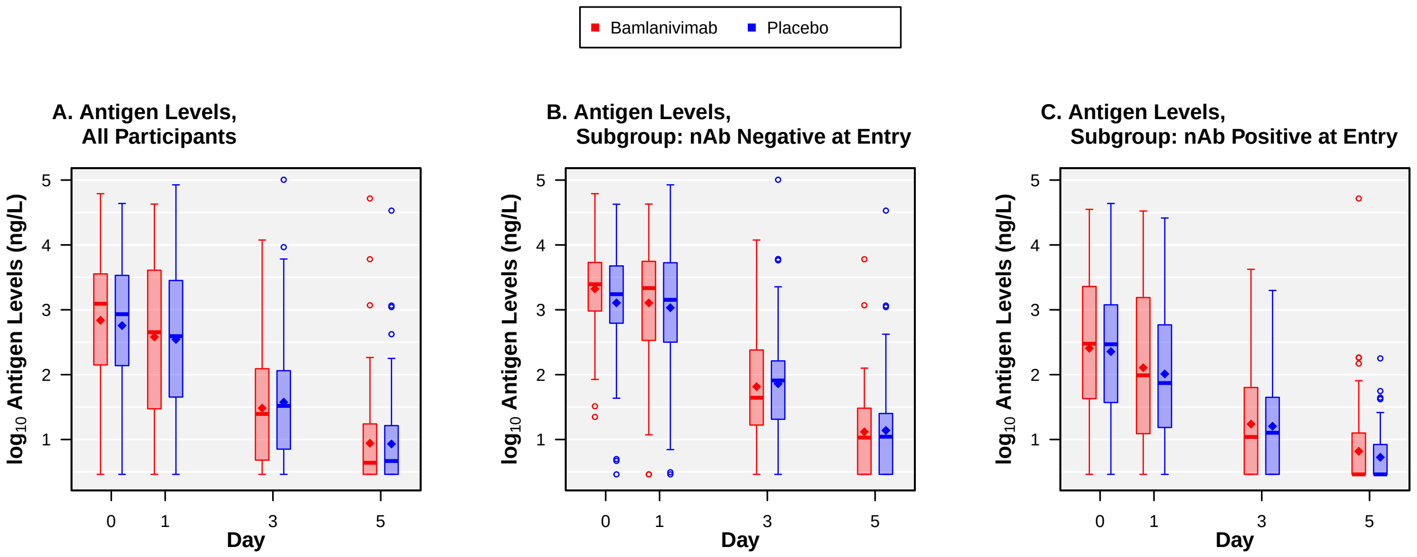

**A. Antigen Levels, All Participants**

| Number of Patients | 0 | 1 | 3 | 5 |
|---|---|---|---|---|
| **Bam.:** | 157 | 155 | 147 | 138 |
| **Placebo:** | 148 | 141 | 142 | 137 |

| Est. Mean Diff. (Bam.−Placebo) | 1 | 3 | 5 |
|---|---|---|---|
| Est. Diff.: | −0.073 | −0.092 | −0.021 |
| 95% CI: | (−0.22, 0.07) | (−0.23, 0.05) | (−0.16, 0.11) |
| P−value: | 0.32 | 0.19 | 0.76 |

**B. Antigen Levels, Subgroup: nAb Negative at Entry**

| Number of Patients | 0 | 1 | 3 | 5 |
|---|---|---|---|---|
| **Bam.:** | 74 | 71 | 66 | 61 |
| **Placebo:** | 79 | 74 | 77 | 70 |

| Est. Mean Diff. (Bam.−Placebo) | 1 | 3 | 5 |
|---|---|---|---|
| Est. Diff.: | −0.094 | −0.20 | −0.12 |
| 95% CI: | (−0.30, 0.11) | (−0.41, 0.006) | (−0.34, 0.09) |
| P−value: | 0.36 | 0.06 | 0.27 |

**C. Antigen Levels, Subgroup: nAb Positive at Entry**

| Number of Patients | 0 | 1 | 3 | 5 |
|---|---|---|---|---|
| **Bam.:** | 83 | 80 | 78 | 74 |
| **Placebo:** | 69 | 64 | 63 | 64 |

| Est. Mean Diff. (Bam.−Placebo) | 1 | 3 | 5 |
|---|---|---|---|
| Est. Diff.: | −0.016 | 0.017 | 0.066 |
| 95% CI: | (−0.22, 0.19) | (−0.16, 0.20) | (−0.10, 0.23) |
| P−value: | 0.87 | 0.85 | 0.42 |

#### Figure S7: Sustained Recovery Outcome by Baseline Demographic Subgroups

*No. (%) of Pts with Sustained Recovery*

| Subgroup | Pct. in Subgrp | Bamlanivimab Pts | Bamlanivimab N | Bamlanivimab (%) | Placebo Pts | Placebo N | Placebo (%) | RRR | P for Interact. |
|---|---|---|---|---|---|---|---|---|---|
| **Age (years)*** | | | | | | | | | 0.517 |
| < 50 | 25 | 39 | 38 | (97) | 40 | 38 | (95) | 1.08 | |
| 50−59 | 22 | 30 | 28 | (93) | 39 | 35 | (90) | 1.02 | |
| 60−69 | 23 | 45 | 37 | (82) | 28 | 26 | (93) | 0.69 | |
| 70−79 | 20 | 38 | 34 | (89) | 25 | 22 | (88) | 1.15 | |
| ≥ 80 | 10 | 11 | 7 | (64) | 19 | 15 | (79) | 0.77 | |
| **Sex** | | | | | | | | | 0.833 |
| Male | 56 | 97 | 84 | (87) | 80 | 71 | (89) | 0.95 | |
| Female | 44 | 66 | 60 | (91) | 71 | 65 | (92) | 1.00 | |
| **Race** | | | | | | | | | 0.340 |
| Black | 21 | 33 | 31 | (94) | 34 | 31 | (91) | 0.79 | |
| Hispanic | 24 | 41 | 36 | (88) | 33 | 28 | (85) | 1.28 | |
| White/Other | 55 | 89 | 77 | (87) | 84 | 77 | (92) | 0.92 | |
| **Days since symptom onset*** | | | | | | | | | 0.230 |
| ≤ 5 | 32 | 56 | 49 | (88) | 45 | 39 | (87) | 1.07 | |
| 6−8 | 32 | 47 | 43 | (91) | 53 | 48 | (91) | 1.17 | |
| ≥ 9 | 36 | 60 | 52 | (87) | 53 | 49 | (92) | 0.80 | |
| **Region** | | | | | | | | | 0.345 |
| USA | 88 | 143 | 124 | (87) | 132 | 117 | (89) | 0.94 | |
| Europe or Asia | 12 | 20 | 20 | (100) | 19 | 19 | (100) | 1.28 | |
| **Type of residence** | | | | | | | | | 0.967 |
| Independent living | 95 | 157 | 139 | (89) | 142 | 129 | (91) | 0.95 | |
| Other | 5 | 6 | 5 | (83) | 9 | 7 | (78) | 0.94 | |
| **Borg respiratory scale*** | | | | | | | | | 0.205 |
| 0−1 | 31 | 53 | 45 | (85) | 41 | 39 | (95) | 0.66 | |
| 2−3 | 38 | 56 | 56 | (100) | 60 | 55 | (92) | 1.54 | |
| ≥ 4 | 31 | 51 | 41 | (80) | 44 | 38 | (86) | 0.88 | |
| **NEW score*** | | | | | | | | | 0.975 |
| ≤ 2 | 30 | 42 | 39 | (93) | 53 | 50 | (94) | 0.78 | |
| 3−5 | 44 | 81 | 74 | (91) | 58 | 52 | (90) | 1.20 | |
| ≥ 6 | 25 | 40 | 31 | (78) | 40 | 34 | (85) | 0.87 | |
| **Baseline pulmonary category*** | | | | | | | | | 0.863 |
| Not on supplemental $O_2$ | 28 | 44 | 42 | (95) | 44 | 42 | (95) | 0.87 | |
| Supp. $O_2$ , flow rate < 4 L/min | 37 | 60 | 58 | (97) | 55 | 51 | (93) | 1.39 | |
| Supp. $O_2$ , flow rate ≥ 4 L/min | 20 | 29 | 24 | (83) | 34 | 31 | (91) | 0.83 | |
| HFNC/non−invasive vent. | 15 | 30 | 20 | (67) | 18 | 12 | (67) | 1.00 | |
| **Overall** | 100 | 163 | 144 | (88) | 151 | 136 | (90) | 0.99 | |

**RRR(Bam./Placebo) (95% CI)**

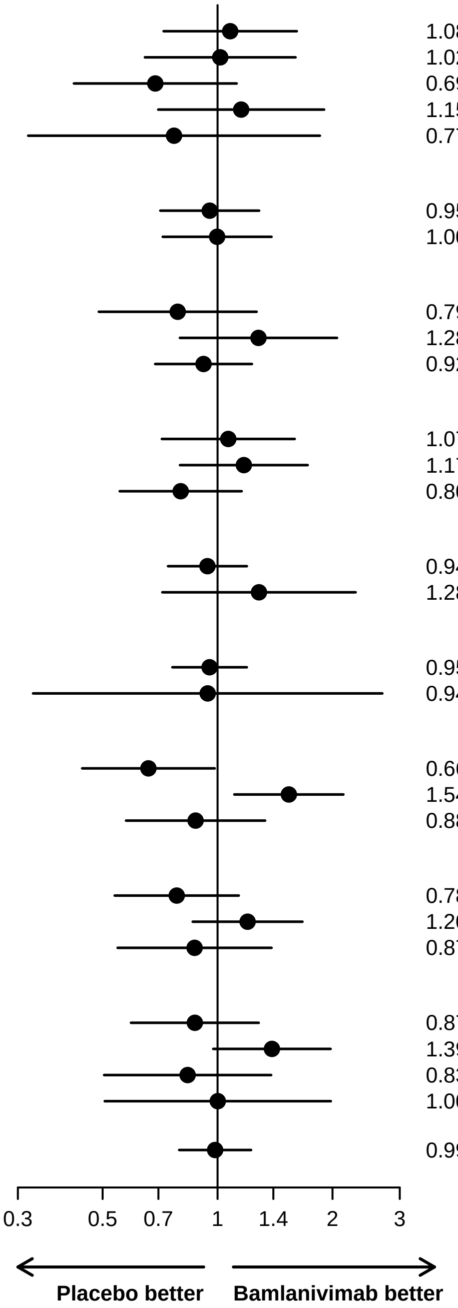

* The interaction p−value is for trend, using the continuous subgroup variable

#### Figure S8: Sustained Recovery Outcome by Anti-N Antibody Subgroups

*No. (%) of Pts with Sustained Recovery*

| Subgroup | Pct. in Subgrp | Bamlanivimab Pts | Bamlanivimab N | Bamlanivimab (%) | Placebo Pts | Placebo N | Placebo (%) | RRR | P for Interact. |
|---|---|---|---|---|---|---|---|---|---|
| **Anti−N Ab (Ab)** | | | | | | | | | 0.050 |
| Positive (Ab+) | 59 | 93 | 81 | (87) | 87 | 81 | (93) | 0.81 | |
| Negative (Ab−) | 41 | 64 | 58 | (91) | 61 | 52 | (85) | 1.27 | |
| **Antigen (Ag)** | | | | | | | | | 0.046 |
| < 1000 pg/mL (low) | 50 | 71 | 65 | (92) | 81 | 78 | (96) | 0.80 | |
| ≥ 1000 pg/mL (high) | 50 | 86 | 74 | (86) | 67 | 55 | (82) | 1.26 | |
| **Viral RNA** | | | | | | | | | 0.180 |
| < 10,000 cp/mL (low)* | 49 | 75 | 67 | (89) | 72 | 68 | (94) | 0.85 | |
| ≥ 10,000 cp/mL (high) | 51 | 79 | 69 | (87) | 72 | 61 | (85) | 1.15 | |
| **Antigen by Ab** | | | | | | | | | 0.059 |
| Ag high, Ab− | 27 | 47 | 42 | (89) | 35 | 27 | (77) | 1.66 | |
| Ag high, Ab+ | 23 | 39 | 32 | (82) | 32 | 28 | (88) | 0.94 | |
| Ag low, Ab− | 14 | 17 | 16 | (94) | 26 | 25 | (96) | 0.96 | |
| Ag low, Ab+ | 36 | 54 | 49 | (91) | 55 | 53 | (96) | 0.74 | |
| **Viral RNA by Ab** | | | | | | | | | 0.041 |
| RNA high, Ab− | 29 | 44 | 42 | (95) | 43 | 35 | (81) | 1.63 | |
| RNA high, Ab+ | 22 | 35 | 27 | (77) | 29 | 26 | (90) | 0.74 | |
| RNA low, Ab− | 11 | 17 | 13 | (76) | 17 | 16 | (94) | 0.73 | |
| RNA low, Ab+ | 38 | 57 | 53 | (93) | 55 | 52 | (95) | 0.90 | |
| **Antigen and Viral RNA**** | | | | | | | | | 0.090 |
| Ag low & RNA low | 30 | 45 | 42 | (93) | 43 | 42 | (98) | 0.73 | |
| Ag high **or** RNA high | 70 | 108 | 93 | (86) | 101 | 87 | (86) | 1.11 | |
| **Antigen and Viral RNA by Ab **** | | | | | | | | | 0.064 |
| Ag high **or** RNA high, Ab− | 37 | 56 | 51 | (91) | 53 | 44 | (83) | 1.43 | |
| Ag high **or** RNA high, Ab+ | 34 | 52 | 42 | (81) | 48 | 43 | (90) | 0.84 | |
| Ag low & RNA low, Ab− | 4 | 5 | 4 | (80) | 7 | 7 | (100) | 0.52 | |
| Ag low & RNA low, Ab+ | 26 | 40 | 38 | (95) | 36 | 35 | (97) | 0.77 | |
| **Overall** | 100 | 163 | 144 | (88) | 151 | 136 | (90) | 0.99 | |

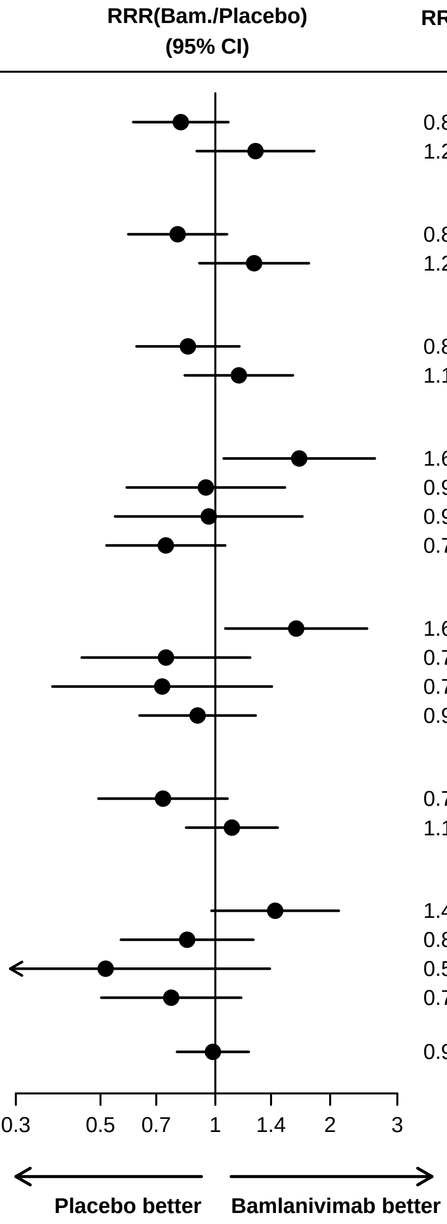

* Viral RNA < 10,000 copies/mL, negative, or indeterminate
** Ag low: Antigen <1000 pg/mL; RNA low: RNA < 10K

**Figure S9: Sustained Recovery Outcome by Subgroups Defined by nAb and Anti-N Antibodies**

*No. (%) of Pts with Sustained Recovery*

| Subgroup | Pct. in Subgrp | Bamlanivimab Pts | Bamlanivimab N | Bamlanivimab (%) | Placebo Pts | Placebo N | Placebo (%) | RRR | P for Interact. |
|---|---|---|---|---|---|---|---|---|---|
| **Neutralizing Ab and anti−N Ab** | | | | | | | | | 0.008 |
| Ab+ by either assay (Ab+) | 66 | 105 | 90 | (86) | 96 | 90 | (94) | 0.78 | |
| Ab− by both assays (Ab−) | 34 | 52 | 49 | (94) | 52 | 43 | (83) | 1.47 | |
| **Antigen (Ag)** | | | | | | | | | 0.046 |
| < 1000 pg/mL (low) | 50 | 71 | 65 | (92) | 81 | 78 | (96) | 0.80 | |
| ≥ 1000 pg/mL (high) | 50 | 86 | 74 | (86) | 67 | 55 | (82) | 1.26 | |
| **Viral RNA** | | | | | | | | | 0.180 |
| < 10,000 cp/mL (low)* | 49 | 75 | 67 | (89) | 72 | 68 | (94) | 0.85 | |
| ≥ 10,000 cp/mL (high) | 51 | 79 | 69 | (87) | 72 | 61 | (85) | 1.15 | |
| **Antigen by Antibody** | | | | | | | | | 0.014 |
| Ag high, Ab− (both assays) | 22 | 38 | 35 | (92) | 30 | 22 | (73) | 1.93 | |
| Ag high, Ab+ (any assay) | 28 | 48 | 39 | (81) | 37 | 33 | (89) | 0.90 | |
| Ag low, Ab− (both assays) | 12 | 14 | 14 | (100) | 22 | 21 | (95) | 1.18 | |
| Ag low, Ab+ (any assay) | 38 | 57 | 51 | (89) | 59 | 57 | (97) | 0.73 | |
| **Viral RNA by Antibody** | | | | | | | | | 0.004 |
| RNA high, Ab− (both assays) | 25 | 39 | 38 | (97) | 35 | 27 | (77) | 2.01 | |
| RNA high, Ab+ (any assay) | 26 | 40 | 31 | (78) | 37 | 34 | (92) | 0.68 | |
| RNA low, Ab− (both assays) | 9 | 12 | 10 | (83) | 16 | 15 | (94) | 0.72 | |
| RNA low, Ab+ (any assay) | 40 | 62 | 56 | (90) | 56 | 53 | (95) | 0.88 | |
| **Antigen and Viral RNA**** | | | | | | | | | 0.097 |
| Ag low & RNA low | 29 | 45 | 42 | (93) | 43 | 42 | (98) | 0.73 | |
| Ag high **or** RNA high | 71 | 111 | 96 | (86) | 102 | 88 | (86) | 1.10 | |
| **Antigen and Viral RNA by Antibody (nAb and Anti−N Ab)**** | | | | | | | | | 0.028 |
| Ag high **or** RNA high, Ab− | 31 | 49 | 46 | (94) | 45 | 36 | (80) | 1.60 | |
| Ag high **or** RNA high, Ab+ | 40 | 62 | 50 | (81) | 57 | 52 | (91) | 0.81 | |
| Ag low & RNA low, Ab− | 3 | 3 | 3 | (100) | 7 | 7 | (100) | 0.82 | |
| Ag low & RNA low, Ab+ | 26 | 42 | 39 | (93) | 36 | 35 | (97) | 0.73 | |
| **Overall** | 100 | 163 | 144 | (88) | 151 | 136 | (90) | 0.99 | |

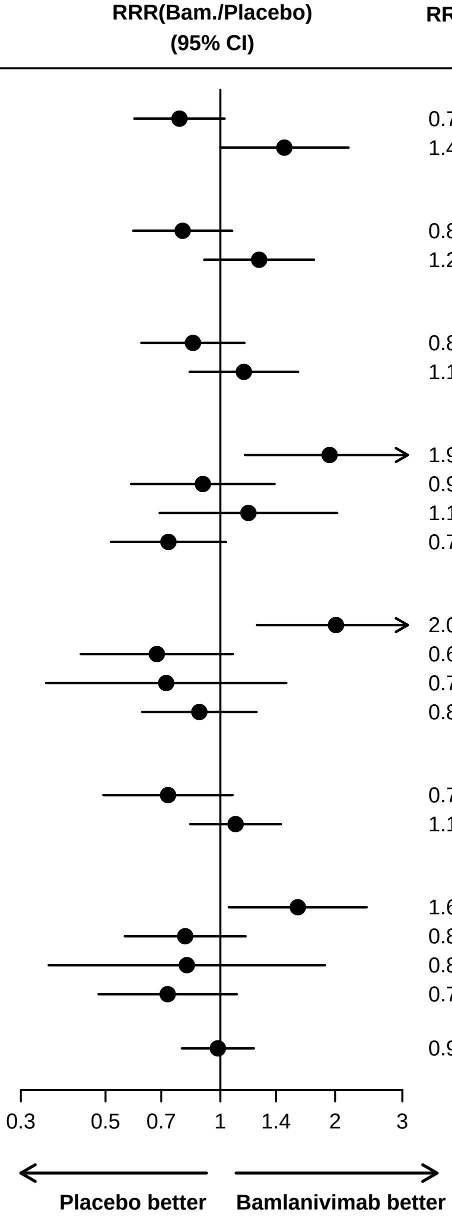

* Viral RNA < 10,000 copies/mL, negative, or indeterminate
** Ag low: Antigen <1000 pg/mL; RNA low: RNA < 10K

Table S6: Number of Participants with Safety Outcomes by Treatment Group

| Event | Bamlanivimab | Placebo | OR or HR | 95% CI | p-value |
|---|---|---|---|---|---|
| Infusion reaction [a] | 23 (14.1%) | 14 (9.3%) | 1.62 | 0.79, 3.32 | .19 |
| Day 5 composite safety outcome [b] | 33 (20.2%) | 21 (13.9%) | 1.66 | 0.87, 3.16 | .13 |
| Day 5 expanded safety outcome [c] | 45 (27.6%) | 28 (18.5%) | 1.83 | 1.01, 3.29 | .05 |
| Day 28 composite safety outcome [d] | 42 (25.8%) | 32 (21.2%) | 1.28 | 0.81, 2.02 | .30 |
| Day 28 expanded safety outcome [e] | 52 (31.9%) | 42 (27.8%) | 1.21 | 0.81, 1.82 | .35 |
| Day 90 composite safety outcome [f] | 45 (27.6%) | 41 (27.2%) | 1.05 | 0.68, 1.60 | .83 |
| Death [g] | 13 (8.0%) | 11 (7.3%) | 1.09 | 0.49, 2.43 | .84 |

[a] Infusion reactions during or within 2 hours following the infusion; odds ratio (OR) estimated from logistic model adjusted for site pharmacy.

[b] Death, SAE, or grade 3/4 AE through Day 5; odds ratio (OR) estimated from logistic model adjusted for site pharmacy.

[c] Death, SAE, or grade 3/4 AE, organ failure, or serious infection through Day 5; odds ratio (OR) estimated from logistic model adjusted for site pharmacy.

[d] Death, SAE, or grade 3/4 AE through Day 28; hazard ratio (HR) estimated from Cox model with stratification by site pharmacy.

[e] Death, SAE, or grade 3/4 AE, organ failure, or serious infection through Day 28; hazard ratio (HR) estimated from Cox model with stratification by site pharmacy.

[f] Death, SAE, organ failure, or serious infection through Day 90; hazard ratio (HR) estimated from Cox model with stratification by site pharmacy.

[g] Death through Day 90; hazard ratio (HR) estimated from Cox model with stratification by site pharmacy.

**Table S7: Safety Outcomes by nAb Subgroups**

| Event | Bamlanivimab | Placebo | OR or HR | 95% CI | Interaction* |
|---|---|---|---|---|---|
| Infusion reaction [a] | | | | | .54 |
| Antibody positive | 9 (10.8%) | 3 (4.3%) | 2.68 | 0.69, 10.30 | |
| Antibody negative | 14 (18.9%) | 10 (12.7%) | 1.61 | 0.67, 3.89 | |
| Day 5 composite safety outcome [b] | | | | | .94 |
| Antibody positive | 15 (18.1%) | 9 (13.0%) | 1.47 | 0.60, 3.60 | |
| Antibody negative | 16 (21.6%) | 12 (15.2%) | 1.54 | 0.67, 3.52 | |
| Day 5 expanded safety outcome [c] | | | | | .94 |
| Antibody positive | 23 (27.7%) | 13 (18.8%) | 1.65 | 0.76, 3.57 | |
| Antibody negative | 20 (27.0%) | 15 (19.0%) | 1.58 | 0.74, 3.38 | |
| Day 28 composite safety outcome [d] | | | | | .62 |
| Antibody positive | 20 (24.1%) | 12 (17.4%) | 1.42 | 0.69, 2.91 | |
| Antibody negative | 20 (27.0%) | 20 (25.3%) | 1.12 | 0.60, 2.08 | |
| Day 28 expanded safety outcome [e] | | | | | .45 |
| Antibody positive | 26 (31.3%) | 16 (23.2%) | 1.42 | 0.76, 2.65 | |
| Antibody negative | 24 (32.4%) | 26 (32.9%) | 1.04 | 0.60, 1.81 | |
| Day 90 composite safety outcome [f] | | | | | .03 |
| Antibody positive | 26 (31.3%) | 13 (18.8%) | 1.79 | 0.92, 3.48 | |
| Antibody negative | 18 (24.3%) | 28 (35.4%) | 0.67 | 0.37, 1.21 | |
| Death [g] | | | | | .04 |
| Antibody positive | 8 (9.6%) | 2 (2.9%) | 3.52 | 0.75, 16.58 | |
| Antibody negative | 4 (5.4%) | 9 (11.4%) | 0.46 | 0.14, 1.48 | |

* p-value for interaction between treatment group and antibody interpretation

[a] Infusion reactions during or within 2 hours following the infusion; odds ratio (OR) estimated from logistic model.

[b] Death, SAE, or grade 3/4 AE through Day 5; odds ratio (OR) estimated from logistic model

[c] Death, SAE, or grade 3/4 AE, organ failure, or serious infection through Day 5; odds ratio (OR) estimated from logistic model

[d] Death, SAE, or grade 3/4 AE through Day 28; hazard ratio (HR) estimated from Cox model

[e] Death, SAE, or grade 3/4 AE, organ failure, or serious infection through Day 28; hazard ratio (HR) estimated from Cox model

[f] Death, SAE, organ failure, or serious infection through Day 90; hazard ratio (HR) estimated from Cox model

[g] Death through Day 90; hazard ratio (HR) estimated from Cox model

Table S8: Components of the Day 90 Safety Outcome by nAb Subgroups

| | Bamlanivimab | | Placebo | | Hazard Ratio* | | |
|---|---|---|---|---|---|---|---|
| **All Participants** | | | | | | | |
| | **Bamlanivimab (n= 163 )** | | **Placebo (n= 151 )** | | **Hazard Ratio*** | | |
| **Events through Day 90** | **Pts** | **Pct.** | **Pts** | **Pct.** | **HR** | **95% CI** | **P-value** |
| SAE | 9 | 5.5 | 12 | 7.9 | 0.70 | 0.29, 1.65 | .41 |
| Death | 13 | 8.0 | 11 | 7.3 | 1.09 | 0.49, 2.43 | .84 |
| SAE or Death | 22 | 13.5 | 20 | 13.2 | 1.03 | 0.56, 1.89 | .93 |
| Organ Failure | 37 | 22.7 | 31 | 20.5 | 1.14 | 0.71, 1.83 | .60 |
| Any of Above | 45 | 27.6 | 41 | 27.2 | 1.05 | 0.68, 1.60 | .83 |
| **Among Participants with Negative nAb at Baseline** | | | | | | | |
| | **Bamlanivimab (n= 74 )** | | **Placebo (n= 79 )** | | **Hazard Ratio*** | | |
| SAE | 4 | 5.4 | 8 | 10.1 | 0.51 | 0.15, 1.70 | .27 |
| Death | 4 | 5.4 | 9 | 11.4 | 0.46 | 0.14, 1.48 | .19 |
| SAE or Death | 8 | 10.8 | 15 | 19.0 | 0.55 | 0.23, 1.29 | .17 |
| Organ Failure | 15 | 20.3 | 20 | 25.3 | 0.78 | 0.40, 1.53 | .48 |
| Any of Above | 18 | 24.3 | 28 | 35.4 | 0.67 | 0.37, 1.21 | .18 |
| **Among Participants with Positive nAb at Baseline** | | | | | | | |
| | **Bamlanivimab (n= 83 )** | | **Placebo (n= 69 )** | | **Hazard Ratio*** | | |
| SAE | 5 | 6.0 | 4 | 5.8 | 1.10 | 0.30, 4.11 | .88 |
| Death | 8 | 9.6 | 2 | 2.9 | 3.52 | 0.75, 16.58 | .11 |
| SAE or Death | 13 | 15.7 | 5 | 7.2 | 2.29 | 0.82, 6.43 | .11 |
| Organ Failure | 22 | 26.5 | 11 | 15.9 | 1.78 | 0.86, 3.68 | .12 |
| Any of Above | 26 | 31.3 | 13 | 18.8 | 1.79 | 0.92, 3.48 | .09 |

* Hazard ratio by Cox regression. Overall is stratified by site pharmacy.

#### Figure S10: Plasma IL-6 and CRP Levels by Treatment Group and by nAb at Entry

■ Bamlanivimab ■ Placebo

**A. IL−6,**
**All Participants**

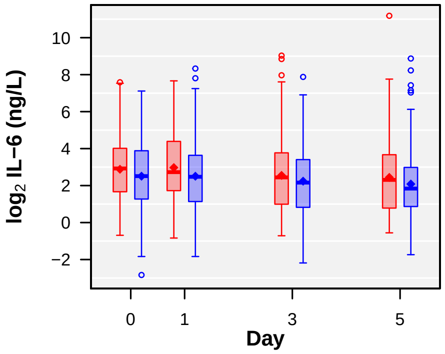

| | | | | |
|---|---|---|---|---|
| **Number of Patients** | | | | |
| **Bam.:** | 156 | 153 | 146 | 137 |
| **Placebo:** | 148 | 141 | 142 | 136 |

| | | | |
|---|---|---|---|
| **Est. Mean Diff. (Bam.−Placebo)** | | | |
| Est. Diff.: | 0.19 | 0.11 | 0.17 |
| 95% CI: | (−0.14, 0.53) | (−0.27, 0.49) | (−0.27, 0.61) |
| P−value: | 0.26 | 0.56 | 0.45 |

**B. IL−6,**
**Subgroup: nAb Negative at Entry**

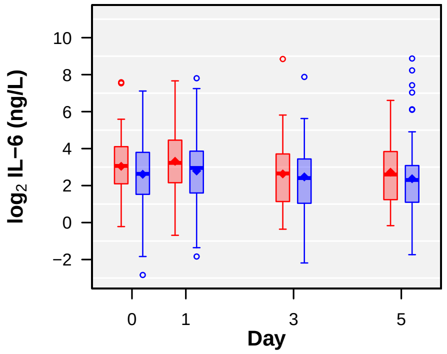

| | | | | |
|---|---|---|---|---|
| **Number of Patients** | | | | |
| **Bam.:** | 74 | 71 | 66 | 61 |
| **Placebo:** | 79 | 74 | 77 | 70 |

| | | | |
|---|---|---|---|
| **Est. Mean Diff. (Bam.−Placebo)** | | | |
| Est. Diff.: | 0.22 | −0.14 | 0.06 |
| 95% CI: | (−0.22, 0.65) | (−0.63, 0.36) | (−0.58, 0.70) |
| P−value: | 0.33 | 0.59 | 0.85 |

**C. IL−6,**
**Subgroup: nAb Positive at Entry**

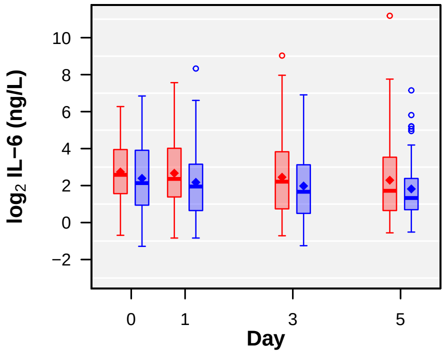

| | | | | |
|---|---|---|---|---|
| **Number of Patients** | | | | |
| **Bam.:** | 82 | 78 | 77 | 73 |
| **Placebo:** | 69 | 64 | 63 | 63 |

| | | | |
|---|---|---|---|
| **Est. Mean Diff. (Bam.−Placebo)** | | | |
| Est. Diff.: | 0.23 | 0.39 | 0.32 |
| 95% CI: | (−0.28, 0.75) | (−0.21, 0.98) | (−0.29, 0.94) |
| P−value: | 0.37 | 0.20 | 0.30 |

**D. CRP,**
**All Participants**

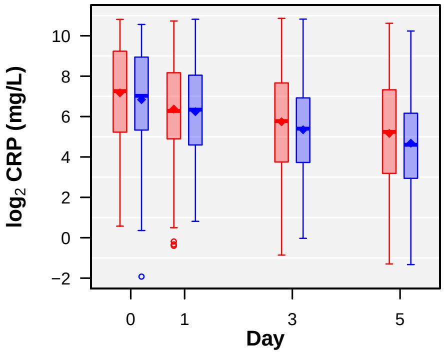

| | | | | |
|---|---|---|---|---|
| **Number of Patients** | | | | |
| **Bam.:** | 149 | 150 | 143 | 133 |
| **Placebo:** | 146 | 135 | 140 | 131 |

| | | | |
|---|---|---|---|
| **Est. Mean Diff. (Bam.−Placebo)** | | | |
| Est. Diff.: | −0.18 | 0.23 | 0.34 |
| 95% CI: | (−0.62, 0.26) | (−0.25, 0.70) | (−0.22, 0.90) |
| P−value: | 0.43 | 0.34 | 0.24 |

**E. CRP,**
**Subgroup: nAb Negative at Entry**

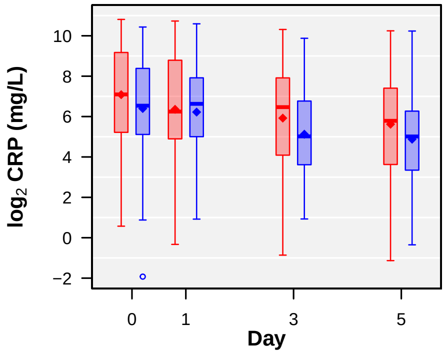

| | | | | |
|---|---|---|---|---|
| **Number of Patients** | | | | |
| **Bam.:** | 69 | 69 | 64 | 59 |
| **Placebo:** | 78 | 71 | 75 | 68 |

| | | | |
|---|---|---|---|
| **Est. Mean Diff. (Bam.−Placebo)** | | | |
| Est. Diff.: | −0.28 | 0.48 | 0.48 |
| 95% CI: | (−0.93, 0.37) | (−0.22, 1.18) | (−0.33, 1.30) |
| P−value: | 0.39 | 0.18 | 0.24 |

**F. CRP,**
**Subgroup: nAb Positive at Entry**

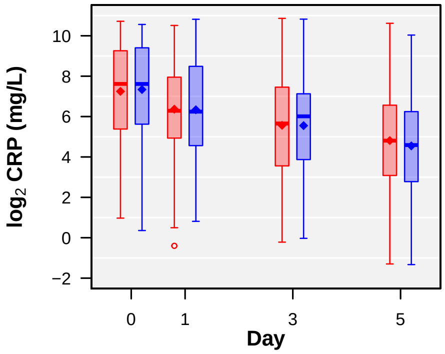

| | | | | |
|---|---|---|---|---|
| **Number of Patients** | | | | |
| **Bam.:** | 80 | 77 | 77 | 71 |
| **Placebo:** | 68 | 62 | 63 | 60 |

| | | | |
|---|---|---|---|
| **Est. Mean Diff. (Bam.−Placebo)** | | | |
| Est. Diff.: | −0.044 | 0.056 | 0.30 |
| 95% CI: | (−0.65, 0.56) | (−0.60, 0.72) | (−0.49, 1.08) |
| P−value: | 0.89 | 0.87 | 0.46 |

#### Figure S11: Plasma D-dimer Levels by nAb at Entry

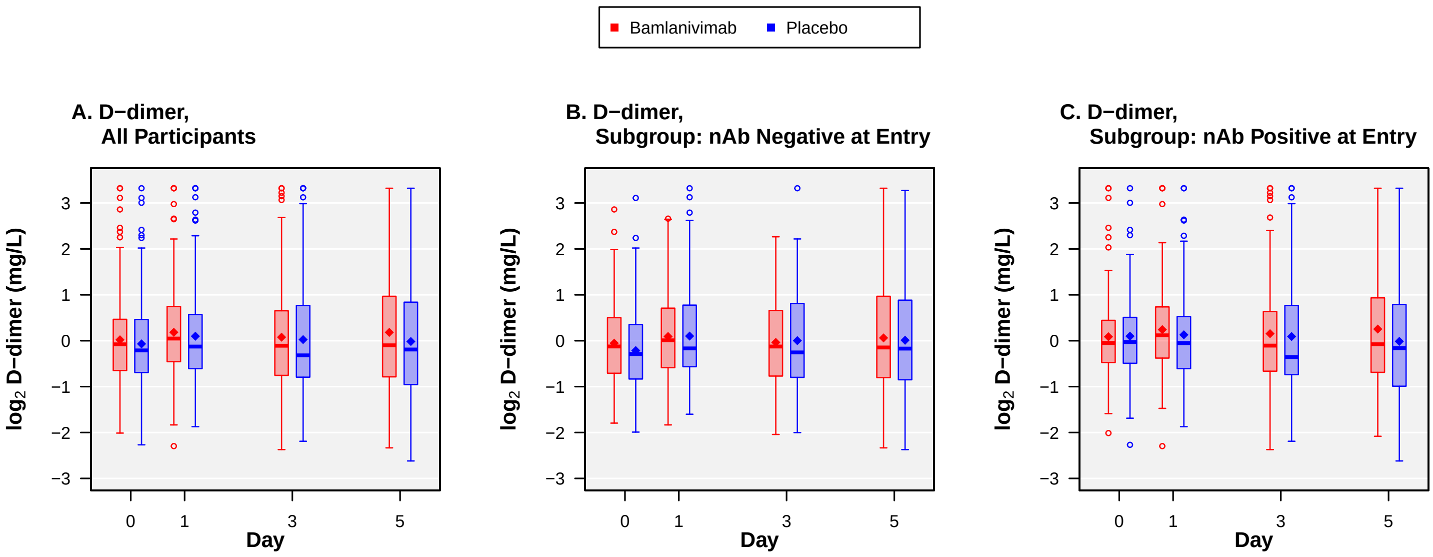

**A.**

| **Number of Patients** | 0 | 1 | 3 | 5 |
|---|---|---|---|---|
| **Bam.:** | 156 | 153 | 146 | 137 |
| **Placebo:** | 148 | 141 | 141 | 135 |

| **Est. Mean Diff. (Bam.−Placebo)** | 1 | 3 | 5 |
|---|---|---|---|
| Est. Diff.: | 0.013 | −0.045 | 0.084 |
| 95% CI: | (−0.13, 0.16) | (−0.24, 0.15) | (−0.16, 0.33) |
| P−value: | 0.87 | 0.65 | 0.50 |

**B.**

| **Number of Patients** | 0 | 1 | 3 | 5 |
|---|---|---|---|---|
| **Bam.:** | 74 | 71 | 66 | 61 |
| **Placebo:** | 79 | 74 | 77 | 70 |

| **Est. Mean Diff. (Bam.−Placebo)** | 1 | 3 | 5 |
|---|---|---|---|
| Est. Diff.: | −0.13 | −0.22 | −0.03 |
| 95% CI: | (−0.35, 0.088) | (−0.47, 0.032) | (−0.40, 0.34) |
| P−value: | 0.24 | 0.09 | 0.87 |

**C.**

| **Number of Patients** | 0 | 1 | 3 | 5 |
|---|---|---|---|---|
| **Bam.:** | 82 | 78 | 77 | 73 |
| **Placebo:** | 69 | 64 | 62 | 62 |

| **Est. Mean Diff. (Bam.−Placebo)** | 1 | 3 | 5 |
|---|---|---|---|
| Est. Diff.: | 0.18 | 0.14 | 0.21 |
| 95% CI: | (−0.0059, 0.37) | (−0.17, 0.44) | (−0.13, 0.55) |
| P−value: | 0.06 | 0.38 | 0.23 |

### Addendum to Statistical Analysis Plan (Version 1.0) Therapeutics for Inpatients with COVID-19 (TICO) ACTIV-3 INSIGHT 014

**14 April 2021**

The Statistical Analysis Plan (SAP) for the ACTIV-3 substudy of LY-CoV555 and its matching placebo is described in Version 1.0 of the SAP. This addendum specifies statistical analyses of SARS-CoV-2 antibody and antigen levels measured in plasma, and SARS-CoV-2 qualitative and quantitative measurements of SARS-CoV-2 RNA in viral transport media (also referred to as RNA) from a mid-turbinate swab sample.

**Study Population:**
The study population will include all participants in the TICO trial of LY-CoV555 who have received any of the investigational agent/placebo (modified intention-to-treat [mITT]) (314 of 326 randomized participants). This is the study population for whom the main study results were reported in the initial NEJM publication.[1] Participants without a baseline specimen collected will be excluded for this study population. Currently, 297 participants have baseline antigen results, 295 have baseline antibody results and 304 have baseline RNA results. The number with antigen and antibody results will increase by a small amount based on recently received specimens at ABML.

#### 1. Assays and Data Collection

##### 1.1 SARS-CoV-2 Antibody and Antigen Data

SARS-CoV-2 antibody and antigen levels were measured in plasma specimens collected at baseline (Day 0), Day 1, Day 3, and Day 5. Antibody and antigen levels were determined centrally, by the NIH, NIAID laboratory.

Antibody levels in plasma were measured using two assays:

- SARS-CoV-2 antibody assay by **Bio-Rad**, measuring total (IgA, IgG, and IgM) anti-nucleoprotein (NP) (Platelia SARS-CoV-2 Total Ab, Bio-Rad, Hercules, CA, USA).

  Results of this antibody measurement are defined in terms of “specimen ratios”. Specimen ratios are defined as the specimen optical density (OD) divided by the optical density of the cut-off control R4 ($OD_M R4$). According to the manufacturer, specimen ratios less than 0.8 are considered negative, those with a specimen ratio between 0.8 and 1.0 are considered equivocal, and those $\geq$ 1.0 are considered positive for the presence of anti-SARS-CoV-2 antibodies.

  **Comment:** Equivocal Bio-Rad antibody levels will be combined with negative levels for all analyses unless otherwise stated.

- SARS-CoV-2 anti-spike neutralizing antibody (nAb) surrogate by **GenScript**, measuring a subset of antibodies capable of inhibiting binding by spike proteins (SARS-CoV-2 Surrogate Virus Neutralization Test [sVNT], GenScript, Piscataway, NJ, USA).

  Neutralizing antibody levels by GenScript are expressed as percent binding inhibition. Specimens with levels less than 30% are considered nAb negative (30% is the manufacturer's cutoff for positivity).

**Comments:**
- For the purpose of comparing natural immunity and whether nMAbs have potential benefit in those with such, the GenScript assay will be used. This attempts to quantify neutralizing titers, whereas the Bio-Rad assay captures any type of antibody against NP (a section of virus not causing neutralization).
- Conversely, for the purpose of understanding whether antibody production by the host in some way affects antigen levels, the Bio-Rad assay is preferred because it identifies antibodies against the same virus antigen as is quantified by the Quanterix antigen assay.

Levels of SARS-CoV-2 nucleoprotein antigen in plasma were measured using an assay made by **Quanterix** (Simoa® SARS-CoV-2 N Protein Advantage, Quanterix, Bellerica, MA, USA). For this assay, antigen levels of < 3 pg/mL (the lower level of quantification) are considered "antigen negative".

**Comments:**
- When analyzed as continuous variable, Quanterix antigen levels <3 will be set to 2.9 pg/mL.
- When analyzed as continuous variable, antigen levels will be $\log_{10}$-transformed.

Baseline antibody levels (both assays) and antigen levels, obtained for the TICO LY-Cov555 trial, are summarized in Table A-1 in the Appendix. Out of 295 participants with antibody data, 112 (38%) were antibody negative by Bio-Rad; 147 (50%) were nAb negative by GenScript; and 13 (4%) of 297 participants had negative antigen levels by Quanterix.

Associations between plasma antigen levels and plasma antibody levels at baseline are summarized in Table A-2. For both assays, antigen levels are lower among participants who are antibody positive.

Table A-3 summarizes the bivariate distribution of baseline antibody and antigen levels for those who were antibody positive. Higher antibody levels among antibody positive participants were associated with lower antigen levels. This association was stronger for GenScript (anti-NP) and NP antigen levels (rank correlation = -0.50) than for the association between Bio-RAD (anti-spike) and NP antigen levels (rank correlation = -0.13).

Table A-4 summarizes antibody and antigen levels and percent positive according to days from symptom onset. Antibody levels are higher among those participants with longer symptom duration; antigen levels don't follow a linear trend, antigen levels appear highest 5-7 days after symptom onset.

These tables informed subgroups analyses described in section 2.2 below.

#### 1.2 SARS-CoV-2 RNA Data

Qualitative and quantitative assessments of SARS-CoV-2 RNA in viral transport media (proxy for viral load) by RT-PCR from mid-turbinate nasal swabs were determined at baseline. The qualitative and quantitative assessments were made centrally by ABML.

- *Qualitative RT-PCR analysis:* Extraction, master mix preparation, and RT-PCR were performed as described in the CDC 2019-Novel Coronavirus Real-Time RT-PCR Diagnostic Panel. RT-PCR was performed on an Applied Biosystems QuantStudio 7 Flex. Ct scores <40 for both nCoV N1 and nCoV N2 probe sets are scored as positive for the presence of SARS-CoV-2 RNA.
- *Quantitative RT-PCR analysis:* Quantitative RT-PCR analysis of the samples used the same RNA extracts prepared for the qualitative assay. Assay conditions were the same as outlined in the CDC protocol except the RNaseP probe was not used.

Samples were available for 304 participants, collected at baseline. Of these, 247 (81%) were SARS-CoV-2 positive, 54 (18%) were negative, and 3 (1%) were indeterminate. The distribution of levels (copies/µL) which were positive for SARS-CoV-2 are given in Table 1 below. This summary was provided by ABML and categories may be modified when the raw data are received.

**Comments:**

- For most analyses, RNA levels in viral transport media will be categorized as negative, <1000, 1,000-50,000, and > 50,000 copies/µL. Indeterminate levels may be combined with negative levels for selected analyses.
- When analyzed as continuous variable, RNA levels to a level TBD when raw data are received.
- When analyzed as continuous variable, RNA levels will be $\log_{10}$-transformed.

**Table 1. SARS-CoV-2 Qualitative RT-PCR Results at Baseline, Among SARS-CoV-2 Positive Participants**

| Bin | SARS-CoV-2 copies/µL | Number of PIDs |
|---|---|---|
| 1 | <1,000 | 92 |
| 2 | 1,000 - 50 K | 99 |
| 3 | 50 K - 200 K | 26 |
| 4 | 200 K - 400 K | 6 |
| 5 | 400 K - 1 M | 4 |
| 6 | 1 M - 4 M | 9 |
| 7 | 4 M - 50 M | 10 |
| 8 | >50 M | 1 |

##### 1.3 Biomarkers of Inflammation and Coagulation

Interleukin-6 (IL-6), D-dimer and hsCRP will be assessed centrally from stored plasma samples at baseline (Day 0), Day 1, 3, and 5.

**Comments:**
- When analyzed as continuous variable, these biomarkers will be $\log_2$-transformed.

#### 2 Associations between Clinical Outcomes for LY-CoV555 and Baseline Antibody and Antigen Levels

Associations between clinical outcomes for LY-CoV555 and baseline SARS-CoV-2 antibody and antigen levels will be assessed using subgroup analyses, as described in Section 8.6 of the SAP. Subgroups will also be considered by SARS-CoV-2 RNA levels.

The overall results and the subgroup findings by days since symptoms onset of the trial of LY-Cov555 in *hospitalized patients* did not reveal a treatment difference in either the intermediate pulmonary ordinal outcome at Day 5 or in the primary endpoint of sustained recovery. However, a growing body of evidence supports the possibility that anti-SARS-CoV-2 monoclonal antibodies may alter viral pathogenesis when given early following infection, before an immune response to the infection has been initiated.

Summaries of baseline antibody and antigen levels according to days since symptom onset revealed a trend – those with longer duration of symptoms had higher antibody levels than those with a shorter duration of symptoms. For example, 37% of participants with duration of symptoms < 5 days were antibody positive by the GenScript assay; this percentage was 71% for those enrolled with duration of symptoms of 10 or more days at the time of randomization.(Table A-4). This suggests that subgroup analyses that take into account antibody levels may be more informative that those by duration of symptoms.

Thus, in spite of no overall treatment effect and no evidence of a treatment by subgroup interaction according to days since symptom onset, subgroup analyses that consider baseline antigen and antibody levels were undertaken. Our general hypothesis is that the population enrolled in the LY-CoV55 trial was heterogenous and that there may be subgroups defined by antigen, antibody and RNA levels for which the treatment may be beneficial. Specific hypotheses are below. Findings from these exploratory analyses will be considered hypothesis generating and will be used to define analyses for other monoclonal antibodies studied in TICO (ACTIV-3).

**Hypothesis 1:** Patients with negative or lower positive neutralizing antibody levels (GenScript) will benefit more from the investigational agent compared to placebo than patients with higher antibody levels. Furthermore, those with lower neutralizing antibody levels AND with higher antigen levels, will benefit more from the investigational agent compared to placebo than other subgroups categorized by both antibody and antigen levels.

**Hypothesis 2:** Patients with lower neutralizing antibody levels (Genscript) AND with higher levels of RNA in nasal turbinates will benefit more from the investigational agent compared to placebo than other subgroups categorized by both antibody and RNA levels.

While our primary hypotheses consider the neutralizing antibody levels, we will test similar hypotheses for the total antibody levels.

##### 2.1 Clinical Outcomes Considered for Subgroup Analyses

Subgroup analyses will be performed for the primary and intermediate efficacy outcomes, and a composite safety outcome. Below we are listing the clinical outcomes, and the statistical methodology:

- *Time to recovery (primary efficacy outcome).* Within each baseline subgroup, median time to recovery will be estimated (by treatment group) using Aalen-Johansen estimates for the cumulative incidence function (CIF). Recovery rate ratios (RR) comparing the treatment groups within each subgroup will be estimated with 95% CIs using Fine-Gray models containing the treatment group indicator. The p-value for a differential treatment effect across subgroups will be estimated as described below.
- *Ordinal Pulmonary outcome on Day 5 (intermediate efficacy outcome):* Within each baseline subgroup, the mean category level will be estimated (by treatment group). Odds ratios (OR) comparing the treatment groups within each subgroup for the odds of being in a better category will be estimated with 95% CIs using a proportional odds model containing the treatment group indicator. The p-value for a differential treatment effect across subgroups will be estimated as described below.
- *Composite of grade 3 and 4 events, SAEs, organ failure, serious infections, or death through Day 28.* Within each baseline subgroup, the number and % of participants with the event will be calculated (by treatment group). Hazard ratios (HR) comparing the treatment groups for time to first event within each subgroup will be estimated with 95% CIs using a Cox proportional hazards regression model containing the treatment group indicator. The p-value for a differential treatment effect across subgroups will be estimated as described below.

**Tests for differential treatment effects across subgroups:** As described in the current SAP, tests for differential treatment effects will be carried out, by testing for interactions between the subgroup indicator and treatment indicator variables in the corresponding joint models across subgroups. In addition to the interactions based on the categorical subgroup indicators, the interaction effect will be estimated in models that include the subgrouping variable (baseline antibody level, antigen level, or RNA) as continuous variables, $\log_{10}$-transformed if necessary, for interaction tests; the latter will be the primary interaction test.

**Stratification and covariates:**

- Models for the analysis of the ordinal pulmonary ordinal outcome will contain the indicators for the baseline pulmonary categories as covariates.
- Because of potentially low sample sizes in subgroup analyses, models will not be stratified by study pharmacy.

#### 2.2 Definition of Baseline Subgroups by Antigen and Antibody Levels and by SARS-CoV-2 RNA levels.

Distributions of antibody and antigen levels at baseline are summarized in Table A-1 on page 14, and the joint distributions of antibody and antigen levels are described in Tables A-2 and A-3 on pages 15-16. Based on these distributions, we formulated the following subgroups defined by each antibody assay alone, by antigen level alone, and by antibody and antigen level jointly:

1. Antibody positive vs antibody negative (**Bio-Rad**). Those in the antibody negative group have a specimen ratio less than 0.8; those antibody positive have specimen ratios ≥ 1.0. Four groups will be defined, above and below the median of 3.40 for positives, and above and below the median for negatives (<0.14 vs. ≥0.14). In this analysis of 4 groups, those with equivocal results (9 patients) will be combined with those negative and above 0.14.
2. Antibody positive vs antibody negative (**GenScript**). Those in the antibody negative group have a binding inhibition percent < 30%; those antibody positive are all ≥ 30%. Fifty percent of participants are antibody negative (Table A-1). Approximate quartiles will also be considered (<13%, 13-29.9, 30-59.9, and ≥ 60%). The first 2 quartiles are for antibody negative patients and the 3rd and 4th quartiles are for antibody positive patients.
3. **Antigen** level < 1000 pg/mL vs antigen level ≥ 1000 pg/mL; the cut-point of 1000 pg/mL is close to the median of 994 pg/mL. The group < 1000 pg/mL includes all those classified as antigen negative.
   Approximate quartiles will also be considered (< 100 pg/mL, 100-999.9, 1000-2999.9, and ≥ 3000 pg/mL). The group < 100 pg/mL includes all those classified as antigen negative.
4. **Bivariate combinations of antigen and antibody levels:** To understand the joint relationship of antibody and antigen levels on major outcomes the following 4 groups will be formed: i) antibody negative and antigen ≥ 1000 pg/mL(approximate median); ii) antibody positive and antigen ≥ 1000 pg/mL; iii) antibody negative and antigen < 1000 pg/mL; and iv) antibody positive and antigen < 1000 pg/mL. This subgroup analysis will be carried out for both the Bio-Rad and GenScript assays. Among 295 patients in Table A-2, the percentage in groups i) through iv) are 26%, 24%, 15%, and 35% using the Bio-Rad antibody assay (9 patients with equivocal results are included in the antibody negative group). For the GenScript antibody assay, these percentages are 33%, 17%, 17%, and 33%, respectively.

   We will also separately perform this analysis defining antibody positivity as being positive for EITHER assay, and negative otherwise. Counting the 9 patients with equivocal results as “antibody negative”, 194 of the 295 patients (66%) were positive on EITHER assay. Of the 295 patients, 128 (43%) were positive on both assays; 101 were negative on both (34%); 46 (16%) were positive on the Bio-Rad assay and negative on GenScript; and 20 (7%) were negative on the Bio-RAD assay and positive on the Genscript assay.

As indicated in the hypothesis, we expect the investigational agent to have the most favorable response compared to placebo for subgroup i) (antibody negative and high antigen level). The least favorable response for the investigational agent compared to placebo is expected among those in group iv) (antibody positive and low antigen level).

5. Subgroups by **SARS-CoV-2 RNA** level (proxy for viral load from NP swab): negative, <1000, 1000-50K, and >50K copies/µL.

6. Bivariate combinations of SARS-CoV-2 RNA and antigen levels: To understand the joint relationship of RNA and antigen levels on major outcomes, the following groups will be formed: *TBD when RNA levels are provided*.

### 3 Antibody and Antigen Levels During Follow-up

Plasma antibody and antigen levels are available at baseline, Days 1, 3, and 5. To compare treatment groups for longitudinal changes in antibody and antigen levels, the following analyses will be performed. This section expands on Section 10 of the SAP version 1.

#### 3.1 Antigen levels through follow-up

**Hypothesis:** Over the first 5 days, antigen levels will decline faster in those receiving the investigational agent compared to those receiving placebo.

Our primary analysis to address this hypothesis will be to compare treatment groups for the percent antigen negative at Day 5. Additional secondary analyses are described in this section, based on three main approaches for modeling changes in antibody levels over time: as percent of participants who are antibody negative (binary outcome) at each time point, as ordinal categorical outcomes, and as continuous outcomes (estimating the rate of change over time).

Table A-4 on page 18 summarizes the observed plasma antigen levels at baseline, Days 1, 3, and 5, pooled across treatment groups. At each time point, distributions of participants across the following categories were summarized: <3 pg/mL (antigen negative), 3-9.9, 10-99.9, 100-999.9, 1000-2999.9, 3000+ pg/mL.

Antigen levels over time (Baseline, Days 1, 3, and 5) will be described using the following summary statistics, by treatment group:

- Side-by-side box plots of antigen levels
- Number and percent of participants in each of the following antigen categories: <3 (negative), 3-9.9, 10-99.9, 100-999.9, 1000-2999.9, 3000+ pg/mL (ordinal outcome).
- Geometric mean titers (GMT) of the antigen levels

The **primary analysis** will be to compare treatment groups for the *proportion of participants with negative antigen levels on day 5*. The proportion of participants who are antigen negative will be compared between treatment groups using a logistic regression model that includes the treatment indicator, and the baseline $\log_{10}$-transformed antigen level as continuous covariate. An unadjusted analysis will also be performed.

**Secondary analyses**
Treatment groups will be compared for the following secondary outcomes:

- *Percent of participants who are antigen negative (<3 pg/mL) (Days 1 and 3):* The proportion of participants who are antigen negative will be compared between treatment groups using logistic regression models, similar to the primary analysis.
  - In addition, treatment groups will be compared using a longitudinal GEE model for binary outcomes (logistic link function), to estimate an overall treatment effect (weighted average over time).

    **Comment:** The longitudinal model considers antigen status (negative vs positive) on Days 0, 1, 3, and 5 as response, and includes the treatment indicator, time as continuous variable, a “follow-up visit” indicator (=0 for baseline visits, =1 otherwise), the treatment by follow-up indicator interaction, and the baseline $\log_{10}$-transformed antigen levels as (fixed-effect) covariate. The treatment effect is estimated through the treatment by follow-up interaction.

- *Change in geometric mean titers (GMT) of antigen from baseline to Days 1, 3, and 5:* Analyses will use separate models for each follow-up day, as opposed to one longitudinal model. Treatment groups will be compared using ANCOVA models for the change in $\log_{10}$-transformed antigen levels from baseline to the given day; models contain the treatment indicator, and include the baseline $\log_{10}$-transformed antigen level as continuous covariate. The treatment difference (and 95% CI) will be estimated in the ANCOVA model, and presented as ratio of fold-change on the original scale. For each treatment group, the mean change from baseline for the $\log_{10}$-transformed antigen levels will be estimated and presented as fold-change on the original scale.

- *Ordered categories of antigen levels, on Days 1, 3, and 5:* Treatment groups will be compared for the probability of being in a better category on the given follow-up day using proportional odds models; the treatment effect is estimated as summary OR with 95% CI, models contain the treatment indicator, and include the baseline $\log_{10}$-transformed antigen level as continuous covariate.

- Treatment groups will be compared for the mean *slope of the $\log_{10}$-transformed* *antigen levels over time*, using longitudinal linear mixed models. The slope will be expressed as fold-change in GMT antigen levels (original scale).

> **Comment:** The linear mixed effects model for comparing slopes includes as response the $log_{10}$-transformed antigen levels at baseline and through follow-up, random intercepts and slopes (subject-specific, uncorrelated intercepts and slopes), and as independent (fixed effects) variables the treatment group indicator, time as continuous variable, and their interaction. The treatment effect (comparing slopes) is represented by the interaction between the treatment group indicator and the continuous time variable.

**Subgroup analyses:**
For the **primary antigen outcome** (proportion of participants who are antigen negative on Day 5), subgroup analyses will be conducted for the following baseline subgroups (as defined in section 2.2):

- Baseline antigen level
- Antibody positive by BioRad
- Neutralizing antibody positive by GenScript
- Antibody positive by one or both assays (positive by either versus negative on both)
- Subgroups formed by the 4 combinations Bio-Rad antibody levels (positive vs. negative) and antigen levels (< 1000 vs. $\geq$ 1000 pg/mL)
- Subgroups formed by the 4 combinations GenScript neutralizing antibody levels (positive vs. negative) and antigen levels (< 1000 vs. $\geq$ 1000 pg/mL)
- Pulmonary (ordered) category
- Time since symptom onset
- Age
- Immunosuppressive condition or immunosuppressive treatment at baseline
- Prior use of remdesivir at time of randomization

Subgroup analyses will be conducted using logistic regression models similar to those described in section 2.1 above.

> **Comment:** Within each subgroup, the OR of being antigen negative on day 5 comparing the investigational agent to placebo will be estimated using logistic regression models that contain the treatment indicator. In selected analyses, baseline antigen levels will also be included as a continuous covariate. Heterogeneity of the treatment effect across subgroups will be assessed by testing for the interaction between the treatment indicator and subgrouping variable (continuous if possible) in a joint model.

#### 3.2 Antibody levels by Bio-Rad and GenScript through follow-up

Analyses are largely similar to those described for the antigen levels, and will be specified at a later date when pooled (both treatment groups combined) follow-up levels are available.

### 4 IL-6, D-dimer, and hsCRP Levels During Follow-up

IL-6 and D-dimer levels will be available at baseline (Day 0), Days 1, 3, and 5.

**Hypothesis:**

- Infusion of high titers of neutralizing monoclonal antibodies as compared with placebo, leads to an increase in levels of host reactive biomarkers (e.g., IL-6, D-dimer, and hsCRP) over the first 2-4 days after time of infusion.
- The increase is higher in persons with high versus low baseline antigen levels (as proxy for total body viral replication).

The **primary analysis** will be:

- To compare treatment groups for the *change in IL-6 and D-dimer levels from baseline to Day 3*.
- To compare treatment groups of *change in IL-6 and D-dimer levels from baseline to Day 3* within subgroups by baseline antigen levels, and to test for heterogeneity of the treatment difference across the subgroups.

Treatment groups will be compared using ANCOVA models for the change in $\log_2$-transformed biomarker levels from baseline to Day 3; models contain the treatment indicator, and include the baseline $\log_2$-transformed biomarker level as continuous covariate. The treatment difference (and 95% CI) will be estimated in the ANCOVA model, and presented as ratio of fold-changes on the original scale. For each treatment group, the mean change from baseline for the $\log_2$-transformed biomarker levels will be estimated and presented as fold-change on the original scale.

The subgroup analysis will use similar models; methods were described in detail in section 3.1 for antigen levels.

As secondary analyses, trajectories of IL-6 and D-dimer over time will be described, with pointwise comparisons between treatment groups at Days 1, 3, and 5. IL-6 and D-dimer will be $\log_2$-transformed for the analyses, and results will be presented as GMT ratios (or ratios of GMT fold-changes from baseline) on the original scale. Analyses use ANCOVA models as described in section 3.1 for antigen levels.

### 5 Missing Data and Data Collection Outside of Visit Window

For patients who were discharged from the hospital, it was sometimes not possible for the participant to come back to the hospital for a blood draw on the day the specimen was to be obtained (Day 1, 3 or 5). For the Day 1 visit, 2 participants, for Day 3, 3 participants, and for Day 5, 7 participants, blood was obtained on the day before or after the protocol specified visit. (Table A-6 on page 19)

This is ignored in all of the analyses.

##### 5.1 Missing Antigen Data

Table A-7 on page 20 shows the pattern of missingness of antigen data over time; 225 of 297 participants have complete antigen data. Currently, the percentages with missing data at Day 1, 3 and 5 are 7.4%, 8.4%, and 14.1%, respectively. Most of the missing Day 5 data is for discharged participants (32/42 = 76%). There are now an additional 48 specimens which can sent to the laboratory. With these specimens, the percentages with missing data at Days 1, 3, 5 and will be 3.3%, 5.0%, and 9.3%.

We will assume that the data are missing at random, and will refrain from imputing missing data for the primary analysis. As a **sensitivity analysis**, missing data will be imputed, using multiple imputation based on baseline and follow-up antigen results that precede the missing data and on the participants' hospitalization status at the visit for which the antigen data is missing.

#### 6 Cross-Sectional and Epidemiological Analyses

A number of other analyses besides those focused on the randomized comparison of LY-CoV555 with placebo will be carried out. Some of these are outlined below.

##### 6.1 Correlations between SARS-CoV-2 Antigen Levels in Plasma, SARS-CoV-2 RNA Levels from NP Swabs, and Biomarkers

We will describe correlations between SARS-CoV-2 antigen levels in plasma and SARS-CoV-2 RNA levels from NP swabs at baseline and follow-up visits, by calculating Spearman's correlation coefficients, and cross-tabulation of percent antigen negative/positive versus RNA negative/positive.

To assess whether SARS-COV-2 antigens in the blood or RNA levels from mid-turbinate NP swabs are better markers of the current disease stage and for prognosis, two hypotheses will be tested, given below and in section 6.2.

**Hypothesis:** Correlations between baseline plasma antigen levels and biomarkers of host reaction to the virus infection (e.g. IL-6, D-dimer, and hsCRP) are stronger than correlations between RNA levels from mid-turbinate NP swabs and the same biomarkers.

To address this hypothesis, we will perform cross-sectional analyses of baseline data; the difference median biomarker levels (mean log2-transformed IL-6 and D-dimer) will be compared between those who are antigen positive versus antigen negative, and between those who are RNA-positive versus RNA-negative. In addition to comparing the point estimates, we will assess whether the confidence intervals overlap.
In addition, we will calculate Spearman's correlation coefficients between plasma antigen levels with each of the biomarkers, and between RNA levels and biomarkers.

#### 6.2 Associations Between Clinical Outcomes and Baseline Antibody, Antigen, and RNA Levels

In addition to the analyses that are focused on treatment differences between the investigational agent and placebo, epidemiological analyses will be performed to better understand the prognostic importance of baseline levels of antibody, antigen, and RNA levels with clinical outcomes.

The following clinical outcomes will be considered:

- Time to recovery (primary efficacy outcome).
- Ordinal Pulmonary outcome on Day 5 (intermediate efficacy outcome
- Composite of grade 3 and 4 events, SAEs, organ failure, serious infections, or death through Day 28.

In the TICO trial assessing LY-CoV555, there is no evidence for a treatment difference with respect to any of the clinical outcomes considered for the subgroup analyses. To assess the prognostic importance of baseline antigen and antibody levels, we will pool the treatment groups (investigational agent and placebo) and assess associations between antibody and antigen levels and each of the clinical outcomes using the corresponding regression models (Fine-Gray, proportional odds, logistic regression, and Cox proportional hazards models).

Univariate associations will be estimated using the antibody, antigen, and RNA data ($log_{10}$-transformed as necessary) as independent predictors. Joint associations of antibody and antigen/RNA levels with the risk of clinical outcomes will be estimated using the joint baseline subgroups as described in section 2.2 above. In addition to the univariate analyses, models will be adjusted for age, time since symptom onset, and presence of an immunosuppressive condition or use of an immunosuppressive treatment.

In addition, epidemiological analyses will be performed to compare the relative predictive value of SARS-CoV-2 antigen levels in the blood with RNA levels with respect to clinical outcomes.

**Hypothesis:** In the control group, antigen levels in the blood will correlate better with clinical outcomes than RNA levels in viral transport media from mid-turbinate NP swabs.

To address this hypothesis, we will use regression models for the clinical outcomes (as described above) containing each of the two predictors (plasma antigen and RNA levels) with the model containing both predictors jointly, using likelihood ratio tests. If adding antigen levels to using RNA levels results in a (statistically significant) better model fit compared to using RNA levels alone, but not vice versa, we would consider antigen levels as a better predictor than RNA levels. This analysis will be restricted to participants who received placebo.

### Appendix: Data Tables

#### Table A-1. Baseline antibody and antigen data

**Baseline Antibody and Antigen Data**

| BioRad Sample/Cutoff Ratio | Interpretation: Antibody Positive | Interpretation: Antibody Negative | Equivocal | Total |
|---|---|---|---|---|
| no. (%) | 174 (59%) | 112 (38%) | 9 (3%) | 295 (100%) |
| min, max | 1.06, 5.20 | 0.06, 0.76 | 0.80, 0.95 | 0.06, 5.20 |
| median (IQR) | 3.40 (3.29, 4.85) | 0.14 (0.09, 0.22) | 0.88 (0.85, 0.88) | 2.61 (0.17, 4.63) |
| mean ±SD | 3.79 ± 1.23 | 0.20 ± 0.16 | 0.87 ± 0.05 | 2.34 ± 1.99 |
| **GenScript Antibody Binding Inhibition (%)** | **Antibody Positive** | **Antibody Negative** | | **Total** |
| no. (%) | 148 (50%) | 147 (50%) | | 295 (100%) |
| min, max | 30, 98 | -11, 29 | | -11, 98 |
| median (IQR) | 61 (46, 78) | 11 (4, 19) | | 30 (11, 61) |
| mean ±SD | 61.7 ± 19.7 | 11.6 ± 9.6 | | 36.7 ± 29.5 |
| **Quanterix Antigen Concentration (pg/mL)** | **Antigen Positive** | **Antigen Negative** | | **Total** |
| no. (%) | 284 (96%) | 13 (4%) | | 297 (100%) |
| min*, max | 3.2, 61631 | 0.0, 2.7 | | 0.0, 61631 |
| median (IQR) | 1183 (213, 3645) | 0 (0, 2) | | 994 (141, 3430) |
| mean ±SD | 3355.7 ± 6832.6 | 0.7 ± 1.1 | | 3208.9 ± 6716.1 |
| median (IQR) $\log_{10}$ | 3.1 (2.3, 3.6) | 0.5 (0.5, 0.5) | | 3.0 (2.1, 3.5) |
| mean ±SD $\log_{10}$ | 2.90 ± 0.90 | 0.46 ± 0.00 | | 2.79 ± 1.01 |

* a value of 2.9 will be imputed for a negative antigen result in analyses

Program Name = AnAbStats Run date = 11APR2021

**Table A-2. Baseline Antigen Data by Antibody Positive or Negative**
(GenScript indeterminate antibody values are included as “negative”)

**Baseline Antigen Data by Antibody Interpretation**

| | BioRad Interpretation | | GenScript Interpretation | |
|---|---|---|---|---|
| **Quanterix Antigen** | **Antibody Positive** | **Antibody Negative*** | **Antibody Positive** | **Antibody Negative** |
| < 100 pg/mL | 50 (29%) | 12 (10%) | 52 (35%) | 10 (7%) |
| 100-999 pg/mL | 54 (31%) | 31 (26%) | 46 (31%) | 39 (27%) |
| 1000-2999 pg/mL | 29 (17%) | 38 (31%) | 23 (16%) | 44 (30%) |
| 3000+ pg/mL | 41 (24%) | 40 (33%) | 27 (18%) | 54 (37%) |
| Total | 174 (100%) | 121 (100%) | 148 (100%) | 147 (100%) |
| Median (IQR) | 664 (64, 2802) | 1861 (744, 4190) | 300 (37, 1924) | 2130 (796, 5060) |
| mean ±SD $\log_{10}$ | 2.58 ± 1.07 | 3.11 ± 0.84 | 2.40 ± 1.09 | 3.20 ± 0.74 |

* includes equivocal

Program Name = AbxAg Run date = 11APR2021

#### Table A-3. Bivariate distribution of antigen levels versus antibody levels (tertiles) at baseline, among participants who are antibody positive

**Baseline Antigen Data by Antibody Tertiles among Antibody Positive Samples**

| Quanterix Antigen | BioRad Positive Tertile 1 1-3.29 S/C ratio | BioRad Positive Tertile 2 3.30-4.847 S/C ratio | BioRad Positive Tertile 3 4.848+ S/C ratio |
|---|---|---|---|
| < 100 pg/mL | 12 (20%) | 18 (40%) | 20 (29%) |
| 100-999 pg/mL | 20 (33%) | 7 (16%) | 27 (40%) |
| 1000-2999 pg/mL | 13 (21%) | 6 (13%) | 10 (15%) |
| 3000+ pg/mL | 16 (26%) | 14 (31%) | 11 (16%) |
| Total | 61 (100%) | 45 (100%) | 68 (100%) |
| Median (IQR) | 965 (146, 3170) | 812 (23, 3430) | 370 (70, 1530) |
| mean ±SD $log_{10}$ | 2.80 ± 0.96 | 2.45 ± 1.24 | 2.46 ± 1.01 |

| Quanterix Antigen | GenScript Positive Tertile 1 30-51 % | GenScript Positive Tertile 2 52-73 % | GenScript Positive Tertile 3 74+ % |
|---|---|---|---|
| < 100 pg/mL | 5 (10%) | 14 (28%) | 33 (66%) |
| 100-999 pg/mL | 17 (35%) | 17 (34%) | 12 (24%) |
| 1000-2999 pg/mL | 12 (25%) | 8 (16%) | 3 (6%) |
| 3000+ pg/mL | 14 (29%) | 11 (22%) | 2 (4%) |
| Total | 48 (100%) | 50 (100%) | 50 (100%) |
| Median (IQR) | 1198 (410, 3573) | 601 (74, 2568) | 37 (12, 200) |
| mean ±SD $log_{10}$ | 2.97 ± 0.93 | 2.57 ± 0.97 | 1.68 ± 0.97 |

Program Name = AgxAbTer Run date = 11APR2021

#### Table A-4. Baseline antibody and antigen data by days since symptom onset

##### Baseline Antibody and Antigen Data by Symptom Days Prior to Enrollment

| | BioRad Ab Sample/Cutoff Ratio | | GenScript Ab Binding Inhibition % | | Quanterix Ag Concentration (pg/mL) | |
|---|---|---|---|---|---|---|
| **Symptom Days** | **Median (IQR)** | **N (%) Positive** | **Median (IQR)** | **N (%) Positive** | **Median (IQR)** | **N (%) > LOQ** |
| < 5 days | 0.18 (0.10, 3.34) | 23 (35%) | 12 (4, 45) | 24 (37%) | 448 (74, 2304) | 61 (94%) |
| 5-7 days | 1.41 (0.14, 3.37) | 47 (53%) | 22 (7, 53) | 36 (41%) | 2515 (822, 5146) | 88 (100%) |
| 8-9 days | 3.22 (0.32, 4.85) | 48 (63%) | 33 (19, 62) | 41 (54%) | 883 (108, 2550) | 72 (92%) |
| 10+ days | 3.34 (2.08, 4.85) | 56 (85%) | 54 (26, 78) | 47 (71%) | 654 (44, 2131) | 63 (95%) |
| Total | 2.61 (0.17, 4.63) | 174 (59%) | 30 (11, 61) | 148 (50%) | 994 (141, 3430) | 284 (96%) |

* > 3.0, the level of quantification

Program Name = AnAbxSymp Run date = 12APR2021

#### Table A-5. Antigen levels at baseline (Day 0) and at Days 1, 3, and 5

**Quanterix Antigen Levels by Visit**
**Both Treatment Groups Combined**

| | Day 0 | | Day 1 | | Day 3 | | Day 5 | |
|---|---|---|---|---|---|---|---|---|
| Result (pg/mL) | No. | Pct. | No. | Pct. | No. | Pct. | No. | Pct. |
| < 3 (< LOQ) | 13 | 4.4 | 15 | 5.5 | 47 | 17.3 | 110 | 43.1 |
| 3 - 99.9 | 50 | 16.8 | 73 | 26.5 | 153 | 56.3 | 130 | 51.0 |
| 100 - 999 | 86 | 29.0 | 73 | 26.5 | 56 | 20.6 | 10 | 3.9 |
| 1,000 - 9,999 | 129 | 43.4 | 91 | 33.1 | 15 | 5.5 | 4 | 1.6 |
| 10,000 - 99,999 | 19 | 6.4 | 23 | 8.4 | 1 | 0.4 | 1 | 0.4 |
| Total | 297 | 100.0 | 275 | 100.0 | 272 | 100.0 | 255 | 100.0 |

**Alternate Categorization A**

| | Day 0 | | Day 1 | | Day 3 | | Day 5 | |
|---|---|---|---|---|---|---|---|---|
| Result (pg/mL) | No. | Pct. | No. | Pct. | No. | Pct. | No. | Pct. |
| < 10 | 23 | 7.7 | 34 | 12.4 | 87 | 32.0 | 162 | 63.5 |
| 10 - 999.9 | 126 | 42.4 | 127 | 46.2 | 169 | 62.1 | 88 | 34.5 |
| 1,000 - 2,999 | 67 | 22.6 | 39 | 14.2 | 11 | 4.0 | 3 | 1.2 |
| ≥ 3,000 | 81 | 27.3 | 75 | 27.3 | 5 | 1.8 | 2 | 0.8 |
| Total | 297 | 100.0 | 275 | 100.0 | 272 | 100.0 | 255 | 100.0 |

**Alternate Categorization B**

| | Day 0 | | Day 1 | | Day 3 | | Day 5 | |
|---|---|---|---|---|---|---|---|---|
| Result (pg/mL) | No. | Pct. | No. | Pct. | No. | Pct. | No. | Pct. |
| < 3 | 13 | 4.4 | 15 | 5.5 | 47 | 17.3 | 110 | 43.1 |
| 3 - 9.9 | 10 | 3.4 | 19 | 6.9 | 40 | 14.7 | 52 | 20.4 |
| 10 - 99.9 | 40 | 13.5 | 54 | 19.6 | 113 | 41.5 | 78 | 30.6 |
| 100 - 999.9 | 86 | 29.0 | 73 | 26.5 | 56 | 20.6 | 10 | 3.9 |
| 1,000-2,999.9 | 67 | 22.6 | 39 | 14.2 | 11 | 4.0 | 3 | 1.2 |
| ≥ 3,000 | 81 | 27.3 | 75 | 27.3 | 5 | 1.8 | 2 | 0.8 |
| Total | 297 | 100.0 | 275 | 100.0 | 272 | 100.0 | 255 | 100.0 |

Program Name = AgxVis Run date = 09APR2021

#### Table A-6. Deviation in Timing of Blood Draws

| Deviation in Timing of Blood Draw by Study Day | | | | |
|---|---|---|---|---|
| | Study Visit CRF Completed | | | |
| Day Specimen Obtained | Day 0 | Day 1 | Day 3 | Day 5 |
| 0 | 297 | 0 | | |
| 1 | | 273 | | |
| 2 | | 2 | 0 | |
| 3 | | | 269 | |
| 4 | | | 3 | 3 |
| 5 | | | | 248 |
| 6 | | | | 4 |
| Total | 297 | 275 | 272 | 255 |

Program Name = AgRealDay Run date = 12APR2021

#### Table A-7. Patterns of Missingness for antigen data over time

**Pattern of Missingness in Availability of Antigen Data**

| | Study Visit* | | | |
|---|---|---|---|---|
| **No. Participants** | **Day 0** | **Day 1** | **Day 3** | **Day 5** |
| 3 | X | - | - | - |
| 3 | X | - | - | X |
| 3 | X | - | X | - |
| 13 | X | - | X | X |
| 5 | X | X | - | - |
| 14 | X | X | - | X |
| 31 | X | X | X | - |
| 225 | X | X | X | X |
| 297 | | | | |

* X denotes that an antigen result is available for this visit.

Program Name = AgMissPattern Run date = 12APR2021
